# Parkinson’s disease causality and heterogeneity: a proteogenomic view

**DOI:** 10.1101/2022.03.09.22272131

**Authors:** Sergio Kaiser, Luqing Zhang, Brit Mollenhauer, Jaison Jacob, Simonne Longerich, Jorge Del-Aguila, Jacob Marcus, Neha Raghavan, David Stone, Olumide Fagboyegun, Douglas Galasko, Mohammed Dakna, Bilada Bilican, Mary Dovlatyan, Anna Kostikova, Jingyao Li, Brant Peterson, Michael Rotte, Vinicius Sanz, Tatiana Foroud, Samantha J. Hutten, Mark Frasier, Hirotaka Iwaki, Andrew Singleton, Ken Marek, Karen Crawford, Fiona Elwood, Mirko Messa, Pablo Serrano-Fernandez

**Affiliations:** Translational Medicine Department. Novartis Institutes for Biomedical Research. Basel, Switzerland; Cardiovascular and Metabolism Department. Novartis Institutes for Biomedical Research. Cambridge, USA; Department of Neurology, University Medical Center Göttingen. Göttingen, Germany; Neuroscience Department. Novartis Institutes for Biomedical Research. Cambridge USA; Genome and Biomarker Sciences. Merck Exploratory Science Center. Cambridge, USA; Department of Genetics, Cerevel Therapeutics. Cambridge, USA; Department of Neurosciences, University of Southern California, San Diego. La Jolla, USA; Department of Medical and Molecular Genetics, Indiana University School of Medicine. Indianapolis, USA; Michael J. Fox Foundation for Parkinson’s Research. New York, USA; Laboratory of Neurogenetics, National Institute on Aging, National Institutes of Health. Bethesda, USA; Institute for Neurodegenerative Disorders. New Haven, USA; Laboratory of Neuroimaging, University of Southern California. Los Angeles, USA

**Keywords:** proteomics, genetics, proteogenomics, CSF, pQTL

## Abstract

The pathogenesis and clinical heterogeneity of Parkinson’s disease have been evaluated from molecular, pathophysiological, and clinical perspectives. High-throughput proteomic analysis of CSF has opened new opportunities for scrutinizing this heterogeneity. To date, this is the most comprehensive CSF-based proteomics profiling study in Parkinson’s disease (1103 patients, 4135 proteins). Combining CSF aptamer-based proteomics with genetics we determined protein quantitative trait loci (pQTLs). Analyses of pQTLs together with summary statistics from the largest Parkinson’s disease genome wide association study (GWAS) identified 68 potential causal proteins by Mendelian randomization. The top causal protein, GPNMB was previously reported to be upregulated in the substantia nigra of Parkinson’s disease patients.

We also compared the CSF proteomes of patients and controls. The Parkinson’s disease cohort comprised not only *LRRK2*+ and *GBA*+ mutation carriers but also idiopathic patients. Proteome differences between *GBA+* patients and unaffected *GBA*+ controls suggest degeneration of dopaminergic neurons, altered dopamine metabolism and increased brain inflammation. The proteins discriminating *LRRK2+* patients from unaffected *LRRK2+* controls, revealed dysregulated lysosomal degradation, as well as altered alpha-synuclein processing, and neurotransmission. Proteome differences between idiopathic patients and controls suggest increased neuroinflammation, mitochondrial dysfunction / oxidative stress, altered iron metabolism and potential neuroprotection mediated by vasoactive substances.

Finally, we used proteomic data to stratify idiopathic patients into “endotypes”. The identified endotypes show differences in cognitive and motor disease progression based on the use of previously reported protein-based risk scores.

In summary, we: i) identified causal proteins for Parkinson’s disease, ii) assessed CSF proteome differences in Parkinson’s disease patients of genetic and idiopathic etiology, and. iii) stratified idiopathic patients into robust clinically relevant subtypes. Our findings not only contribute to the identification of new therapeutic targets but also to shaping personalized medicine in CNS neurodegeneration.

## Introduction

Parkinson’s disease is the second most prevalent neurodegenerative disorder.^1^ Parkinson’s disease patients experience selective degeneration and loss of dopaminergic neurons in the substantia nigra *pars compacta*. In most cases, Parkinson’s disease is classified as idiopathic, but a growing set of genetic variants increase Parkinson’s disease risk or accelerate its onset. Many of the identified genes are involved either in mitochondrial or endo-lysosomal biology.^2^ The two most common Parkinson’s disease risk genes are leucine rich kinase 2 (*LRRK2*) and glucosidase beta acid (*GBA*). Mutations in these genes are linked to ∼10% of sporadic cases and up to 30% in specific ethnic subgroups and familial disease.^3^ Some of the genetic variants in *LRRK2* and *GBA* have been associated with specific clinical phenotypes.^4,5^ For both genes, the pathological mutations are thought to exacerbate the toxicity of alpha-synuclein, which – in an aggregated form – contributes to neuronal death and amplifies the neuroinflammatory response.

Clinical heterogeneity of Parkinson’s disease has motivated many disease stratification efforts. Some of those have focused on clinical variables, mostly hypothesis-driven, while others have focused on molecular data, mostly hypothesis-free.^6^ An association between such strata and the Parkinson’s disease risk mutations remains elusive.^7^

Proteins hold great potential as predictors, causal biomarkers and surrogates of disease progression and/or stratification. However, the biological and pathophysiological complexity of Parkinson’s disease, the difficulties of collecting standardized biological samples (especially CSF) from large cohorts throughout the course of disease, and the technical limitations of high-throughput proteomic analyses hamper the identification of biomarkers at a proteomic level. The multicenter Parkinson Progression Marker Initiative (PPMI) was initiated to overcome some of these limitations, particularly in terms of number of samples and clinical data ^8^. In this collaboration, 1103 baseline (not longitudinal) CSF samples from patients and control participants with known status of *LRRK2* and *GBA* pathogenic variants were analyzed using the SomaScan® aptamer-based proteomics platform.^9^ There is also whole genome sequencing data from 804 patients out of the 1103, after quality control. To our knowledge, this is to date the largest proteomic and genetic data set for interrogating causal proteins for Parkinson’s disease in a neurologically relevant biofluid.

The main goals of this study are summarized in Figure 1: i. Parkinson’s disease causal protein identification using mendelian randomization based on proteomics and genetics, ii. identification of differences between Parkinson’s disease patients and controls within and between subcohorts (*LRRK2+, GBA+* and idiopathic), and iii. hypothesis-free stratification of idiopathic Parkinson’s disease patients into clinically relevant endotypes and then compared by means of protein-based risk scores reflecting cognitive and motor progression ^10^.

**Figure 1.**
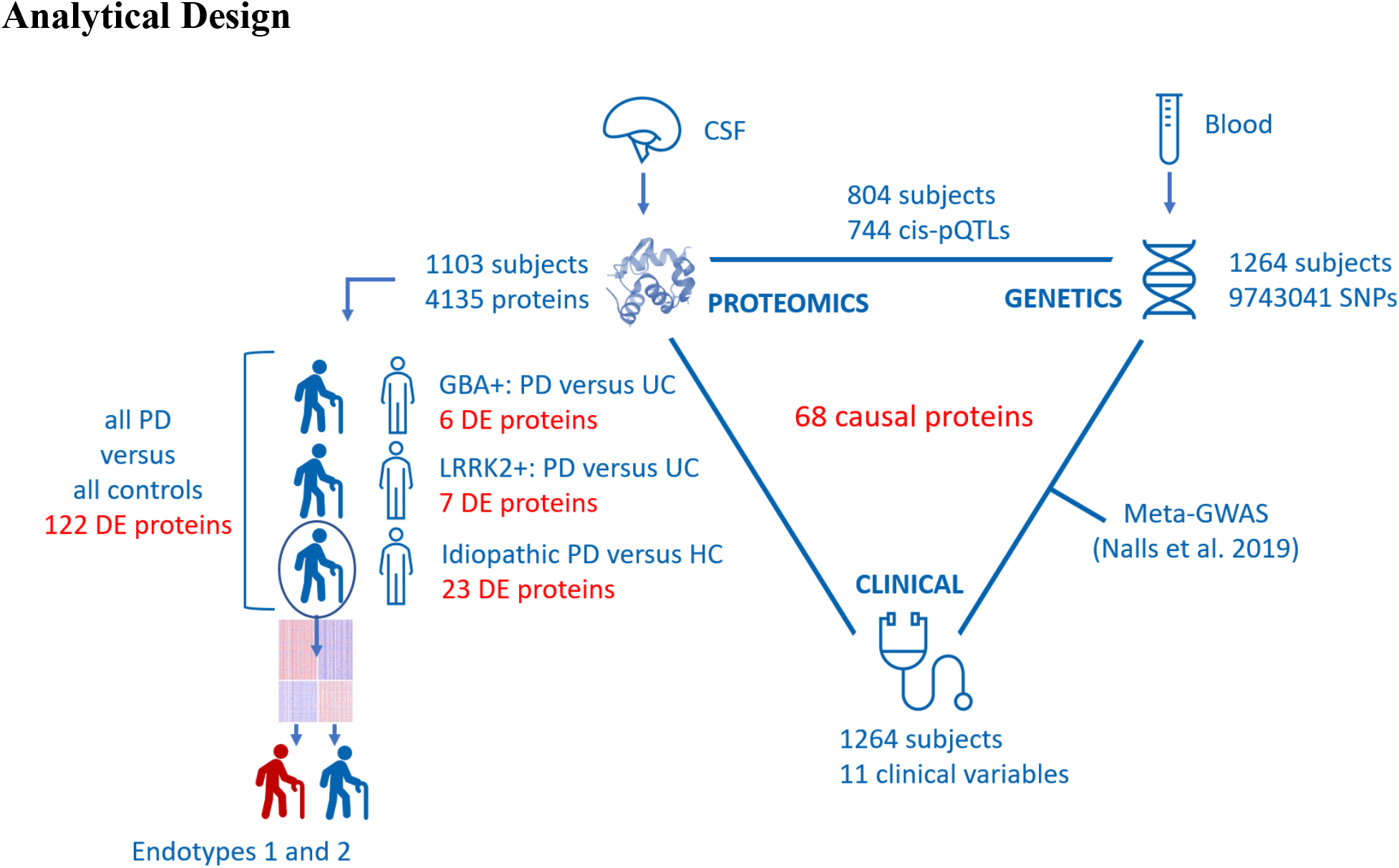
Full analytical design. 1103 subjects were analyzed with SomaScan for 4135 unique proteins in CSF. The comparison between *GBA+* patients and *GBA+* unaffected controls (UC) retrieved six differentially expressed (DE) proteins. The comparison between *LRRK2+* patients and *LRRK2+* UC retrieved seven DE proteins. The comparison between idiopathic patients and healthy controls (HC) non-mutation carriers retrieved 23 DE proteins. Patients and controls were also combined and compared, which retrieved 122 DE proteins. Idiopathic patients were further analyzed, and two endotypes were identified based on CSF proteomics. 1264 subjects were sequenced genome wide to detect a total of 9743041 SNPs. For the 804 patients that had both genomic and proteomic data, a pQTL analysis was performed that identified 744 unique proteins with a significant cis-pQTL. The pQTLs combined with a meta GWAS for Parkinson’s disease performed by Nalls *et al*. (2019), led to the proposal of 68 unique CSF proteins presumed to be causal for Parkinson’s disease.

## Materials and methods

The clinical data and samples used in this study were obtained from the PPMI (http://www.ppmi-info.org/data) on October 1, 2020. PPMI samples were collected under a standardized protocol over 33 centers and includes clinical and imaging data as well as plasma and CSF samples. Study protocol and manuals are available online (http://www.ppmi-info.org/study-design).

Separate subcohorts of patients with Parkinson’s disease and their respective controls were enrolled following inclusion and exclusion criteria.^11^ One subcohort is comprised of recently diagnosed, drug-naïve, idiopathic Parkinson’s disease patients and healthy controls, while the second and third subcohorts are comprised of Parkinson’s disease patients, carriers of a severe *GBA* or *LRRK2* mutation, either Parkinson’s disease patients or unaffected controls. Parkinson’s disease patients from the genetic subcohorts had a higher disease duration, were partially under Parkinson’s disease medication (*n* = 203), were over-represented for individuals of Ashkenazi Jewish descent and differed by sex distribution from the idiopathic Parkinson’s disease patients (higher proportion of men among idiopathic). The study was approved by the Institutional Review Board at each site, and participants provided written informed consent.

Genetic testing was done by the centralized PPMI genetic testing core. Non-manifesting carriers received pre-testing and post-testing genetic counselling by phone from certified genetic counsellors at the University of Indiana or site-qualified personnel. The *LRRK2* genetic testing battery includes G2019S and R1441G mutations. *GBA* genetic testing includes N370S (for all participants), and L483P, L444P, IVS2+1, and 84GG (for a subset of participants) mutations. Dual mutation carriers *(LRRK2* and *GBA)* were considered as *LRRK2* carriers for simplicity (*n* = 1).

Six patients were diagnosed as idiopathic Parkinson’s disease at enrolment but were re-classified during follow-up (two patients were diagnosed as multiple system atrophy and four patients did not have a final diagnose but Parkinson’s disease had been excluded). These patients were removed from the analysis. Four patients were initially diagnosed as genetic Parkinson’s disease, but the diagnose changed to prodromal during follow up. These patients were considered as unaffected controls in their corresponding genetic subcohort (Five *LRRK2*+ unaffected controls and one *GBA*+ unaffected control).

One subject originally classified as healthy, but later shown to have an unclear health status, was removed from the analysis. Subjects recruited into the subcohort of idiopathic Parkinson’s disease patients and healthy controls but identified as carriers of a severe *GBA* or *LRRK2* mutation, were moved to the corresponding genetic subcohort (*GBA n* = 15; *LRRK2 n* = 7).

The genetic screening also detected *GBA* mutations of unknown or moderate risk: A459P, E365K, T408M. Carriers of these mutations were removed from analysis (*n* = 38).

Finally, 10 carriers of a mutation in *SCNA* (eight Parkinson’s disease patients and two unaffected controls), were also removed due to lack of statistical power for analysis.

The original data set was comprised of 1190 samples out of which 32 samples were pools, which were discarded for this study, and as described above, additional six Parkinson’s disease patients and one healthy control were removed due to change in diagnose, 38 subjects were removed due to non-severe *GBA* mutations and 10 patients were removed for being carriers of a mutation in *SCNA*. Hence, the final data set used for analysis was comprised of 1103 proteomic samples divided into three subcohorts: no mutation (idiopathic Parkinson’s disease patients and healthy controls with no severe mutation in *GBA* or *LRRK2*), *GBA*+ (Parkinson’s disease patient carriers of severe *GBA* mutations and unaffected controls carrying the same mutations) and *LRRK2*+ (Parkinson’s disease patients carriers of severe *LRRK2* mutations and unaffected controls carrying the same mutations). The exact composition is summarized in Table I.

**Table I.** Parkinson’s disease subcohorts.

| Subcohort | Patients | Controls |
| --- | --- | --- |
| *GBA*+ | 65 | 162<sup>a</sup> |
| *LRRK2*+ | 154 | 196<sup>a</sup> |
| Idiopathic | 350 | 176<sup>b</sup> |

<sup>a</sup> “unaffected controls” in the main text
<sup>b</sup> “healthy controls” in the main text

We make an explicit distinction between “healthy” and “unaffected” controls, because there is evidence of prodromal pathophysiology in *LRRK2*+ and *GBA*+ controls when compared to healthy controls that are non-mutation carriers.^12^

### Proteomics

Proteomic profiling was performed using SomaScan® in a platform version that is proprietary to Novartis and includes 4785 SOMAmers® (Slow off-rate modified aptamers) targeting 4135 human proteins. SOMAmer levels were determined and standardized at SomaLogic Inc. (Boulder, US) including hybridization normalization (controls for variability in the readout of individual microarrays), plate scaling (accounts for plate-by-plate variation), median signal normalization (controls for total signal differences between individual samples) and calibration (removes the variation between assay runs within and across experiments).

Relative fluorescence units are transformed to log_2_ scale, normalized to the median separately by dilution level across all plates. Finally, the data set is adjusted for batch effects between plates using an empirical Bayes method as implemented in the R package *sva*.^13^

### Genetics

PPMI whole genome sequencing results were lifted over to hg19 coordinates. Biallelic SNPs on autosomes were extracted. Standard GWAS quality control (QC) was applied at both individual and SNP level. 22 patients with outlying heterozygosity and 93 patients with high identity-by-descent were excluded after QC. 306031 SNPs were removed due to missing genotype and 35907596 SNPs removed due to minor allele count less than 20. Finally, 9743041 variants and 1264 subjects passed QC.

### pQTL calculation

Among the 1264 subjects who passed QC for genetics and the 1103 who passed QC for proteomics, 804 subjects overlapped. Protein expression values were ranked and inverse normal transformed. For pQTL calculation, each protein level was regressed with each independent genetic variant (SNP; MAF > 0.05), adjusted for age, sex, subcohort, protein principal components 1-4 and genetic principal components 1-10 using R package *MatrixEQTL*.^14^ Cis-pQTLs were defined as SNPs located inside the +/- 1Mb region flanking the gene that encodes the given protein. A genome-wide threshold of *P* < 5×10^−8^ defined a significant cis-pQTL.

### Causal Analysis

The two-sample mendelian randomization method implemented in the R package *TwoSampleMR*^*15,16*^ was applied to find causal proteins for Parkinson’s disease in CSF. Although PPMI is a well-controlled study with genetic data, to avoid weak instrumental variable bias and take the advantage of a larger Parkinson’s disease GWAS we relied on the meta-analysis from Nalls *et al*.^17^, which includes 17 datasets with Parkinson’s disease cases ranging from 363 to 33674 individuals, and healthy controls ranging from 165 to 449056 individuals. Instrumental variables were selected for each SNP with MAF > 0.05, F-statistics larger than 10 and significant cis-pQTL *P* < 5×10^−8^. Shared SNPs in both cis-pQTL and Parkinson’s disease GWAS were harmonized and then clumped using a linkage disequilibrium (LD) threshold of either *r*^*2*^ < 0.01 or *r*^*2*^ < 0.3. The more stringent LD threshold of r^2^<0.01 resulted in only one instrumental variable for most proteins, therefore the results we present are from the less stringent threshold of r^2^<0.3. The Wald ratio was used when only one instrument survived clumping, while the inverse variance weighted meta-analysis method was used when more than one instrumented SNP was available. Horizontal pleiotropy was tested using the R package *MRPRESSO*.^18^

Colocalization probability was calculated using the R package *coloc*.^19^ Default priors of *p*_*1*_ = 10^−4^, *p*_*2*_ = 10^−4^, and *p*_*12*_ = 10^−5^ were used, where p_1_ is the prior probability of a SNP being associated with Parkinson’s disease, p_2_ is the prior probability of a SNP being associated with CSF pQTL, and *p*_*12*_ is the prior probability of a SNP being associated with both Parkinson’s disease and CSF pQTL. We considered *PPH*_*4*_ > 0.75 as strong evidence for colocalization. *PPH*_*4*_ is the posterior probability of one shared SNP being associated with both Parkinson’s disease and CSF pQTL.

### Clinical variables

The clinical assessment battery is described on the PPMI website (http://www.ppmi-info.org). Parkinson’s disease status was assessed with the Unified Parkinson’s Disease Rating Scale in the revised version published by the Movement Disorder Society (MDS-UPDRS) scores 1, 2, and 3.^20^ Cognitive testing comprised screening with the Montreal Cognitive Assessment (MoCA).^21^ High resolution xy-weighted 3 tesla MRI was available for 545 Parkinson’s disease patients and 177 controls. Caudate, putamen and striatum thicknesses were calculated as the arithmetical mean between the right and left brain hemispheres.

CSF was collected using standardized lumbar puncture procedures. Sample handling, shipment and storage were carried out as described in the PPMI biologics manual (http://ppmi-info.org). Besides the SomaScan analysis described earlier, data from immunoassay kits were also used for measuring CSF total alpha-synuclein, amyloid-beta 1-42, total tau and phospho-tau (p-tau 181) protein as described previously.^22,23^ Phospho-tau was measured with the Elecsys® assay run on the fully automated Roche Cobas® system.

Use of medications for Parkinson’s disease was recorded at each visit after baseline assessment. For simplicity, we used this as a binary variable (medication present / absent).

### Protein risk scores

The Parkinson’s disease protein risk scores used in this study were generated as previously described ^10^. They were defined as “non-genetic Parkinson’s Disease-associated Proteomic Score (ngPD-ProS).

### Statistical analysis

We first compared the protein profiles of Parkinson’s disease patients with controls within each subcohort (idiopathic, *GBA*+ and *LRRK2*+). A linear regression model was applied using the Bioconductor R package *limma*.^24^ The model included the following covariates: age, sex, study center and proteomic principal components 1-4. The genetic subcohorts also included levodopa treatment (yes/no) as a covariate in the model to exclude treatment effects. This was skipped for idiopathic patients as they were drug-naïve.

Additionally, a linear model with an interaction term was tested. The interaction term was between the disease status (case /control) and the mutation status (no mutation / *GBA+* / *LRRK2+*). The covariates were the same as in the models above.

Comparisons of clinical variables between endotypes of idiopathic patients were performed using a chi-squared test for categorical variables or a generalized linear model adjusted for age and sex for quantitative variables. To test differences in age between endotypes a Mann Whitney U-test was used.

Predictive modeling for the idiopathic classes was performed using a partition tree with pruning as implemented in the R package *rpart*.^25^ The model was defined on a training set (70% of the idiopathic patients) and tested on an independent test set (30% of the idiopathic patients). The pruning was based upon a 10-fold cross validation, the default for *rpart*.

All p-values were adjusted for multiple testing using false discovery rate (FDR).

### Cluster analysis

Network analysis of the CSF proteome of idiopathic patients was carried out using the R package *WGCNA*.^26^ Co-expressed proteins (SOMAmers) were grouped into modules.

Consensus clustering as implemented in the R package *ConsensusClusterPlus*^27^ used the SOMAmer modules to identify idiopathic patient subclasses. To avoid confounders being responsible for the differences between patient subclasses, network analysis was performed on the SOMAmer residuals of a linear regression on age, sex and study center.

Heatmaps were generated using the R package *Heatplus*.^28^

### Data availability

The data used for this study is publicly available in the PPMI web page https://www.ppmi-info.org/access-data-specimens/download-data. The free access requires registration. The clinical data snapshot used here is kept under the tab “Archived PPMI data” >> “Publication Associated Archives” >> “2022-0001 Serrano-Fernandez: Parkinson’s Disease Proteogenomics (Version: 2022-05-18)”. The proteomic data is available under “Biospecimen” >> “Proteomic Analysis” >> “Project 151 Identification of proteins & protein networks & pQTL analysis in CSF x of 7 (Batch Corrected)” (7 files in total). The original adat files are also available under “Biospecimen” >> “Proteomic Analysis” >> “Project 151 Identification of proteins & protein networks & pQTL analysis in CSF - ADAT files”.

## Results

### Causal analysis

Our analysis reported significant cis-pQTLs for 856 SOMAmers – corresponding to 744 unique proteins (Supplementary Table 5). From these 856 cis-pQTLs, we identified statistically significant evidence for causation for 68 proteins in CSF (Table 2). Out of those proteins, GPNMB, FCGR2A and FCGR2B also had a strong colocalization signal (see Methods), indicating the same SNP is both associated with protein level and Parkinson’s disease risk (Figure 2). The full tables with nominal *P* ≤ 0.05 are included in Supplementary Tables 1 and 2 for less or more stringent clumping, respectively.

**Table 2.** Parkinson’s disease causal proteins from CSF.

| Protein | Nr of IVs | Method | $\beta$ | FDR | Colocalization PP.H4.abf | Horizontal Pleiotropy Test (MRPRESSO P-value) |
| --- | --- | --- | --- | --- | --- | --- |
| GPNMB<sup>a</sup> | 11 | IVW | 0.14 | $1.17 \times 10^{-17}$ | 0.94 | 0.65 |
| FCGR2B | 17 | IVW | 0.07 | $5.11 \times 10^{-14}$ | 0.96 | 0.49 |
| FCGR2A | 18 | IVW | 0.07 | $2.70 \times 10^{-12}$ | 0.95 | 0.26 |
| CTSB | 12 | IVW | -0.11 | $2.63 \times 10^{-10}$ | 0.22 | 0.47 |
| HLA-DQA2<sup>b</sup> | 33 | IVW | -0.14 | $1.43 \times 10^{-9}$ | 0.03 | 0.00 |
| CD38 | 1 | Wald ratio | -0.53 | $2.45 \times 10^{-9}$ | 0.45 | NA |
| HP | 19 | IVW | 0.06 | $2.01 \times 10^{-6}$ | 0.01 | 0.60 |
| LTF | 23 | IVW | 0.05 | $2.16 \times 10^{-6}$ | 0.04 | 0.65 |
| HAVCR2 | 13 | IVW | -0.10 | $2.87 \times 10^{-5}$ | 0.30 | 0.67 |
| BST1<sup>b</sup> | 10 | IVW | 0.12 | $4.00 \times 10^{-5}$ | 0.24 | 0.00 |
| CLEC3B | 4 | IVW | -0.16 | $1.02 \times 10^{-4}$ | 0.29 | 0.97 |
| HAPLN1 | 15 | IVW | 0.06 | $3.21 \times 10^{-4}$ | 0.05 | 0.79 |
| MANEA | 17 | IVW | 0.05 | $3.21 \times 10^{-4}$ | 0.10 | 0.94 |
| NQO2 | 11 | IVW | 0.05 | $3.21 \times 10^{-4}$ | 0.05 | 0.98 |
| ARSA<sup>a</sup> | 5 | IVW | 0.13 | $6.82 \times 10^{-4}$ | 0.58 | 0.62 |
| LGALS3 | 8 | IVW | 0.07 | $7.11 \times 10^{-4}$ | 0.02 | 0.50 |
| ILIRLI | 26 | IVW | 0.03 | $9.63 \times 10^{-4}$ | 0.01 | 0.90 |
| PAM | 12 | IVW | 0.09 | $9.63 \times 10^{-4}$ | 0.08 | 0.47 |
| TPSAB1 | 8 | IVW | -0.06 | 0.0011 | 0.15 | 0.80 |
| HSP90B1 | 22 | IVW | 0.04 | 0.0011 | 0.03 | 0.19 |
| GLCE | 14 | IVW | 0.05 | 0.0015 | 0.10 | 0.96 |
| MANSC4 | 18 | IVW | 0.05 | 0.0016 | 0.02 | 0.85 |
| RABEPK | 10 | IVW | -0.05 | 0.0021 | 0.10 | 0.86 |
| ICAM1 | 8 | IVW | -0.07 | 0.0021 | 0.32 | 0.74 |
| C4B<sup>b,c</sup> | 101 | IVW | 0.02 | 0.0021 | 0.00 | 0.00 |
| LCT<sup>b</sup> | 25 | IVW | 0.04 | 0.0021 | 0.00 | 0.00 |
| SIRPBI | 25 | IVW | -0.03 | 0.0023 | 0.01 | 0.60 |
| C4A<sup>b,c</sup> | 101 | IVW | 0.02 | 0.0023 | 0.00 | 0.00 |
| PCSK7 | 18 | IVW | -0.04 | 0.0028 | 0.02 | 0.98 |
| IL9 | 12 | IVW | 0.06 | 0.0028 | 0.05 | 0.96 |
| CLN5<sup>a</sup> | 2 | IVW | -0.15 | 0.0028 | 0.38 | NA |
| AGT | 4 | IVW | 0.12 | 0.0039 | 0.06 | 0.97 |
| CD274 | 16 | IVW | 0.08 | 0.0042 | 0.02 | 0.46 |
| RBP7 | 3 | IVW | 0.20 | 0.0047 | 0.09 | NA |
| PLA2G7 | 14 | IVW | 0.04 | 0.0061 | 0.05 | 0.57 |
| EGF | 3 | IVW | -0.13 | 0.010 | 0.11 | NA |
| ASIP | 3 | IVW | -0.13 | 0.010 | 0.17 | NA |
| TPSB2 | 10 | IVW | -0.05 | 0.011 | 0.15 | 0.27 |
| ACPI | 15 | IVW | 0.04 | 0.011 | 0.04 | 0.88 |
| RNASE3 | 6 | IVW | 0.09 | 0.011 | 0.01 | 0.26 |
| A4GALT | 2 | IVW | -0.21 | 0.012 | 0.11 | NA |
| DSCAM | 3 | IVW | -0.30 | 0.015 | 0.19 | NA |
| COLEC11 | 8 | IVW | -0.05 | 0.016 | 0.03 | 0.61 |
| SPOCK2 | 8 | IVW | 0.08 | 0.017 | 0.03 | 0.98 |
| VWA2 | 5 | IVW | −0.11 | 0.018 | 0.04 | 0.76 |
| RPN1 | 10 | IVW | 0.08 | 0.020 | 0.02 | 0.57 |
| ADAMTS4 | 8 | IVW | −0.06 | 0.020 | 0.19 | 0.36 |
| PDCD1LG2 | 12 | IVW | −0.09 | 0.020 | 0.04 | 0.07 |
| PRTN3 | 2 | IVW | 0.17 | 0.020 | 0.06 | NA |
| ADGRE2 | 10 | IVW | −0.05 | 0.020 | 0.01 | 0.87 |
| RNASE2 | 10 | IVW | 0.06 | 0.020 | 0.02 | 0.61 |
| MPIG6B | 18 | IVW | −0.05 | 0.020 | 0.00 | 0.18 |
| SIGLEC9 | 19 | IVW | 0.03 | 0.020 | 0.02 | 0.99 |
| TAPBPL | 10 | IVW | −0.03 | 0.020 | 0.03 | 0.50 |
| LRP12<sup>a</sup> | 1 | Wald ratio | 0.73 | 0.023 | 0.66 | NA |
| DNAJC30 | 3 | IVW | −0.09 | 0.024 | 0.03 | NA |
| CCL15 | 3 | IVW | −0.08 | 0.024 | 0.12 | NA |
| VTN | 22 | IVW | −0.03 | 0.028 | 0.01 | 0.75 |
| NUCB1 | 3 | IVW | −0.12 | 0.029 | 0.05 | NA |
| TRH | 4 | IVW | −0.08 | 0.029 | 0.07 | 0.75 |
| POSTN | 8 | IVW | −0.07 | 0.034 | 0.02 | 0.76 |
| PLXNB2<sup>a</sup> | 36 | IVW | −0.02 | 0.034 | 0.08 | 0.09 |
| IL18RI | 23 | IVW | 0.03 | 0.034 | 0.01 | 0.79 |
| IDUA | 3 | IVW | 0.10 | 0.038 | 0.00 | NA |
| CFD | 10 | IVW | −0.05 | 0.039 | 0.01 | 0.58 |
| GGH<sup>a</sup> | 4 | IVW | 0.08 | 0.040 | 0.02 | 0.49 |
| FGFRL1 | 5 | IVW | 0.13 | 0.046 | 0.00 | 0.07 |
| LMAN2L | 3 | IVW | -0.15 | 0.049 | 0.03 | NA |

<sup>a</sup> PD protein marker in Supplementary Table 3
<sup>b</sup> corrected by removing outliers by MRPRESSO
<sup>c</sup> has homolog detected by the same SOMAmer
744 proteins have a significant cis-pQTL

**Figure 2.**
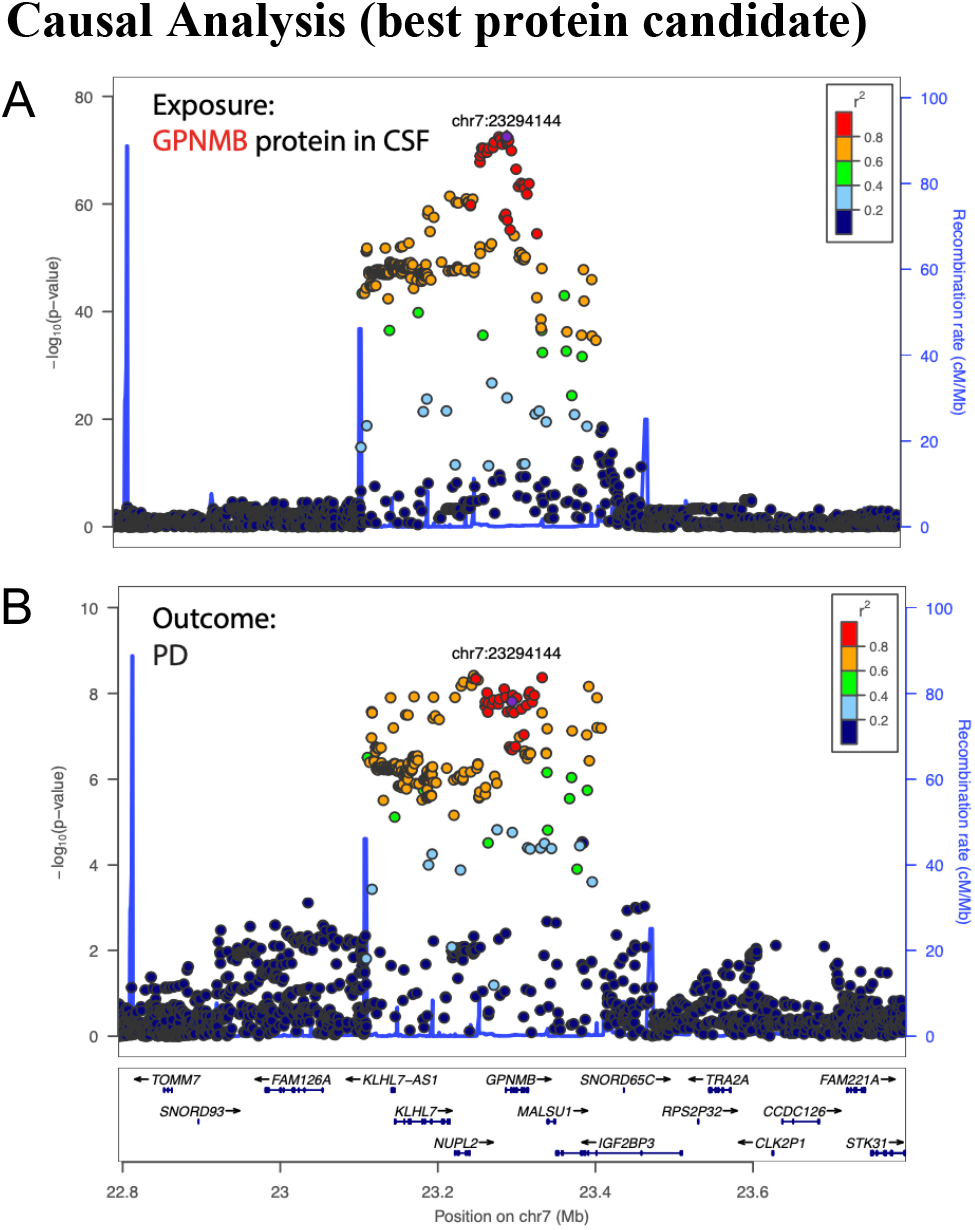
Causal Analysis (best protein candidate). (**A**) Locus visualization of GPNMB pQTL hits suggest a strong association between GPNMB SOMAmer levels and its cis-SNPs. Colors indicate the linkage disequilibrium (LD) correlation of other SNPs with chr7:23294144 (rs858275). (**B**) Locus visualization of GWAS hits in the GPNMB locus for the risk of developing Parkinson’s disease. The y axis is the -log_10_ nominal *P*-value of the GWAS results.

### Differential protein expression in subcohorts of Parkinson’s disease patients

To identify proteins differentially expressed in Parkinson’s disease, we compared SOMAmers in each of the subcohorts (*GBA+, LRRK2+* and idiopathic) to their corresponding controls. Our statistical analyses revealed six differentially expressed SOMAmers for *GBA+* patients, seven SOMAmers for *LRRK2+* patients and 23 SOMAmers for idiopathic patients. Directionality of the change and adjusted *P*-values for each of these markers are reported in Table 3.

**Table 3.** **SOMAmers differentially expressed between Parkinson’s disease patient vs. controls, divided by subcohort**

| Subcohort | Gene Symbol | PD change direction | FDR |
| --- | --- | --- | --- |
| GBA+ | CALCA<sup>48,49, 51</sup> | + | 0.022 |
|  | CD2 | + | 0.025 |
|  | DLKI<sup>46</sup> | – | 0.025 |
|  | GCHI<sup>47, 50</sup> | + | 0.025 |
|  | IL17A<sup>52, 53</sup> | + | 0.025 |
|  | SEMG2 | – | 0.022 |
| LRRK2+ | ARSA<sup>54</sup> | + | 0.023 |
|  | ACP7 | + | 0.043 |
|  | CA10 | + | 0.037 |
|  | CTSB<sup>36, 55, 38-40</sup> | + | 0.037 |
|  |  |  | 0.027<sup>a</sup> |
|  | SIAE | + | 0.037 |
|  | SMPDI<sup>55, 56</sup> | + | 0.037 |
|  | TENM4<sup>58, 72,73</sup> | – | 0.037 |
| Idiopathic | AKI<sup>60</sup> | + | 0.049 |
|  | CCL14 | + | 0.023 |
|  | DLKI<sup>46</sup> | – | 0.032 |
|  | FRZB | + | 0.036 |
|  | GPI<sup>66</sup> | + | 0.006 |
|  | HAMP<sup>64, 65</sup> | + | 0.001 |
| | LPO<sup>63</sup> | – | $1.5 \times 10^{-6}$ |
|  | LRRTM4 | – | 0.023 |
|  | NETO1 | – | 0.049 |
|  | NFH | + | 0.014 |
|  | PTHLH | – | 0.029 |
|  | PTPRR | – | 0.032 |
|  | RAB3I<sup>68</sup> | – | 0.044 |
|  | RELT | – | 0.046 |
| | RIPK2<sup>69</sup> | – | $4.6 \times 10^{-5}$ |
|  | ROBO3 | – | 0.049 |
|  | RSPO4 | – | 0.029 |
|  | SEMG2 | – | 0.029 |
|  | SHANKI<sup>61</sup> | – | 0.023 |
|  | SPINK9 | – | 0.040 |
|  | TXN | – | 0.023 |
|  | VEGFA<sup>70</sup> | – | 0.006 |
|  | VIP<sup>71</sup> | – | 0.023 |

<sup>a</sup> Interaction term.
The change in PD represents (+) increased and (–) decreased in PD vs controls, respectively. The reference numbers close to the gene symbol correspond to literature positions that link those genes with PD.

For each subcohort, several identified markers confirmed previously reported proteins dysregulated in Parkinson’s disease. Interestingly, there was little overlap between proteins dysregulated in *GBA+, LRRK2+* and idiopathic subcohorts (SEMG2 and DLK1 were shared by *GBA+* and the idiopathic subcohort) though in each list there is a high percentage (4/6 in *GBA+*, 4/7 in *LRRK2+* and 10/23 in the idiopathic subcohort) of markers previously reported in relation to Parkinson’s disease (Table 3).

Only one protein, CTSB, passed the FDR significance threshold for the interaction between disease status and mutation status. As shown in Table 3, CTSB was also differentially expressed in the *LRRK2+* subcohort.

Additional analysis comparing all patient subcohorts with all controls, using subcohort membership (no mutation/*GBA*+/*LRRK2*+) and treatment status (yes/no) as additional covariates resulted in 129 SOMAmers, tagging 122 distinct proteins, passing FDR correction (Supplementary Table 3).

### Identification of subtypes of idiopathic Parkinson’s disease patients

#### Identification of endotypes

To determine if distinct endotypes were present in the idiopathic subcohort we performed a network analysis on the CSF proteome of idiopathic patients. Two modules of co-expressed proteins were identified. They comprised 889 and 600 SOMAmers, respectively. Applying consensus clustering on these two protein modules split the idiopathic subcohort (350 patients) in proteome-based patient endotypes 1 (85 patients) and 2 (165 patients) (Figure 3A). As seen in the tracking plot (Figure 3B) these endotypes suffer only negligible changes as the number of modeled subclasses increases. Moreover, a predictive model for the endotypes was built based on clinical parameters, avoiding the re-use of the same proteomic data involved in the definition of the endotypes. Patients with CSF phospho-tau ≥ 11pg/mL (as measured with the Elecsys® assay) were enriched for endotype 2, and patients with CSF phospho-tau < 11pg/mL were enriched for endotype 1 (Figure 3C). The model accuracy in the training (244 patients) and in the independent test (106 patients) sets, was 0.82 and 0.73, respectively. The estimated area under the curve (AUC) for the test set was 0.77.

**Figure 3.**
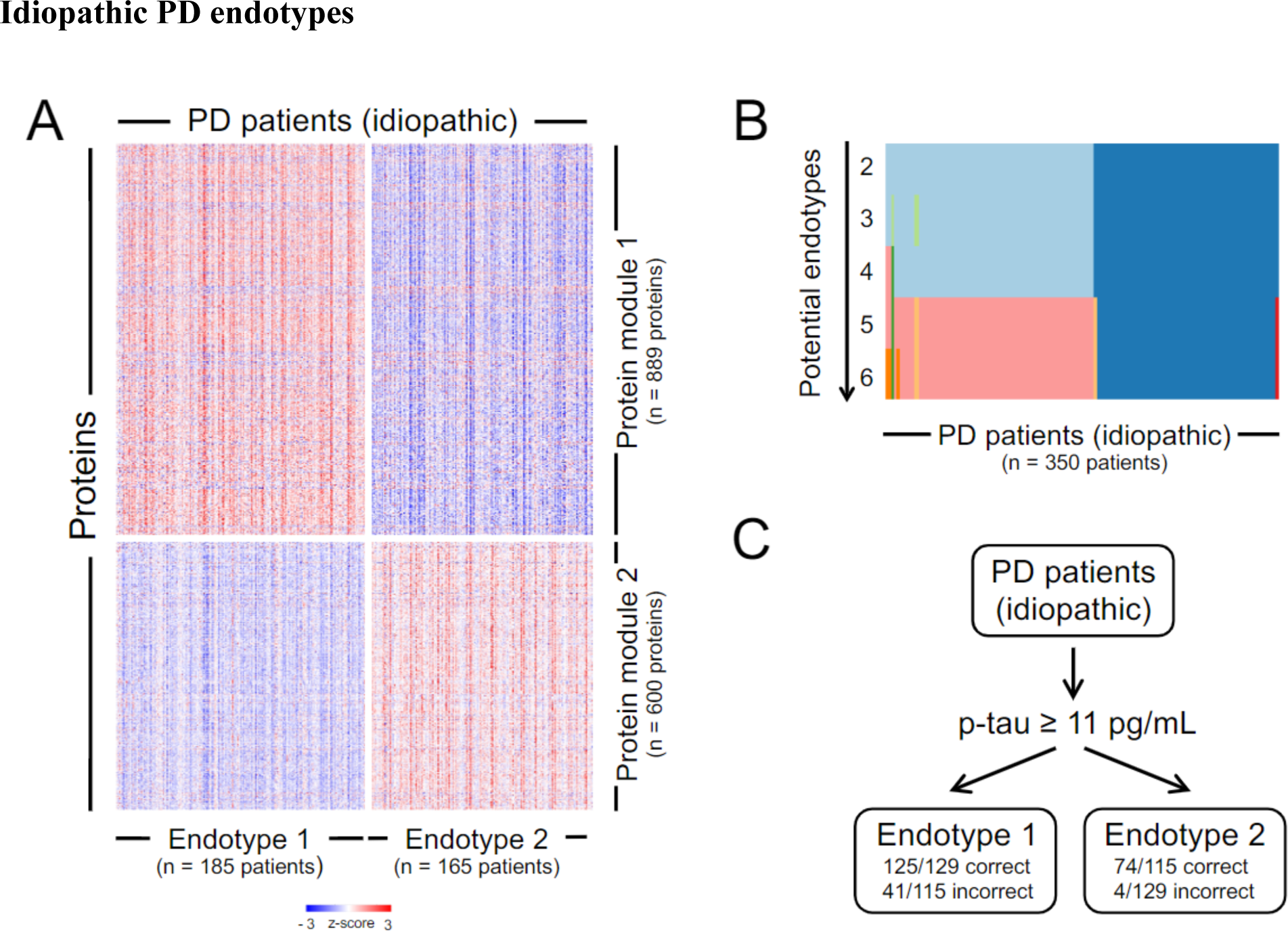
Idiopathic Parkinson’s disease endotypes. (**A**) Heatmap of z-scores of the protein values as measured with SomaScan, corresponding to the two modules identified using Weighted Gene Co-expression Network Analysis (WGCNA). The proteins in these modules are used for cluster analysis using Consensus Clustering, which retrieves two clusters (endotypes) of idiopathic patients. Patients are shown in the x-axis, separated by endotype, while proteins are shown in the y-axis, separated by module. (**B**) Tracking plot depicting how the idiopathic patients are assigned to specific endotypes by Consensus Clustering as the number of potential endotypes increases. (**C**) Partition Tree predicting endotype membership of the idiopathic patients based on clinical variables only. One node suffices to separate patients into endotypes based on phospho-tau levels (p-tau) in CSF as measured with a clinical assay, the cutoff being 11 pg/mL.

Endotypes did not significantly differ in age or sex. There were neither significant differences in caudate, striatum or putamen thickness, nor in UPDRS score parts II and III or MoCA scores. Significant differences were found for UPDRS score part I (higher for endotype 2; *P* = 0.044), as well as CSF levels of amyloid beta (lower for endotype 2; *P* = 1.95×10^−15^.), phospho-tau (lower for endotype 2; *P* =1.25×10^−15^.), total tau (lower for endotype 2; *P* < 2×10^−16^.) and alpha-synuclein (lower for endotype 2; *P* < 2×10^−16^). Endotypes also showed significant differences in ngPD-ProS scores (higher for endotype 2; *P* = 0.034).

### Proteins differentially expressed in endotypes (CSF SomaScan)

To identify the unique proteins significantly dysregulated in each endotype, a linear model was used to identify the differences between each of these two endotypes and the healthy controls to the idiopathic subcohort. For endotype 1, five markers were significantly different compared to the control group (CNTFR, LPO, MMP10, RIPK2 and VEGFA). LPO, RIPK2 and VEGFA were also part of the differences between healthy controls and the whole idiopathic group (see above). Endotype 2, however, showed 200 differentially expressed SOMAmers, 197 unique proteins (see Supplementary Table 4). Among those proteins, AK1, CCL14, FRZB, GPI, HAMP, LPO, NETO1, PTPRR, RAB31, RELT, RIPK2, ROBO3, RSPO4, SHANK1, SPINK9, VEGFA and VIP were dysregulated for the whole idiopathic group as compared to healthy controls. Differentially expressed SOMAmers between endotypes added up to 155, 153 unique proteins (see Supplementary Table 4).

## Discussion

### Causal analysis

Independently of its etiology (genetic or idiopathic), multiple cellular and metabolic alterations (e.g. iron metabolism), inflammation, and oxidative stress underly neurodegeneration in Parkinson’s disease. In this regard, the identification of causal proteins not only enhances its understanding, but also assists the search for druggable targets. This study identified 68 CSF proteins as causal and GPNMB, FCGR2A, FCGR2B and CTSB were among the top ones.

GPNMB - expressed by myeloid cells - is found at high levels in the substantia nigra of Parkinson’s disease patients.^29^ It has been suggested that GPNMB modulates immune response ^30,31^ having primarily a protective role.^32-35^ CTSB cleaves alpha-synuclein fibrils with the potential for decreasing alpha-synuclein aggregation.^36^ FCGR2A and FCGR2B are involved in phagocytosis and modulate inflammatory responses.^37^ Supporting our findings, independent reports suggest that GPNMB, CTSB and FCGR2A are causal to Parkinson’s disease.^38-40^ FCGR2B - aggregated alpha-synuclein interaction inhibits its microglial phagocytosis, promoting neurodegeneration.^41^ However, blockade of FCGR2B in neurons may suppress Lewy body-like inclusion body formation.^42^

### Genetic patients

GBA and LRRK2 are enzymes involved in ceramide metabolism and lysosomal degradation of aggregated alpha-synuclein. Genetic variants of the *GBA* and *LRRK2* genes are known risk factors for the development of Parkinson’s disease and dementia associated with accumulation of Lewy bodies.^43-45^ To better understand the differences between diseased and unaffected mutation carriers we compared their CSF proteomes (see Table 3).

Protein differences between *GBA*+ patients and unaffected *GBA*+ controls are related to brain dopaminergic neurons (DLK1), dopamine metabolism (CALCA, GCH1) and inflammation effector cells (IL17A). All these proteins have been previously linked to Parkinson’s disease, though not specifically to *GBA*+. DLK1 is coupled to both tyrosine hydroxylase expression and neurotrophic signaling.^46^ Significantly lower levels of DLK1 in *GBA*+ patients suggest increased degeneration of dopaminergic neurons. GCH1 and CALCA are involved in nigrostriatal dopamine synthesis,^47^ and dopamine release and metabolism,^48,49^ respectively. We found both elevated in *GBA*+ patients. *GCH1* variants contribute to the risk and earlier age-at-onset of Parkinson’s disease.^50^ Moreover, CALCA was reported as significantly elevated in the CSF of depressed Parkinson’s disease patients relative to major depressive disorder patients.^51^ One of the inflammatory mechanisms proposed for neurodegeneration in Parkinson’s disease involves Th17 cells.^52^ Our study found increased IL17A levels in *GBA+* patients. It has been recently proposed that elevated IL17A plays a key role in neurodegenerative diseases.^53^

Among the CSF proteins differentially expressed between *LRRK2*+ patients and unaffected *LRRK2*+ controls, several stood out for their relevance in Parkinson’s disease: ARSA, SMPD1, CTSB and TENM4.ARSA is a lysosomal chaperone that prevents alpha-synuclein aggregation, secretion and cell-to-cell propagation.^54^ We found ARSA elevated in *LRRK2*+ patients, which could be interpreted as a protection mechanism to prevent the formation of alpha-synuclein aggregates. CTSB (causal protein, see causal analysis) and SMPD1 play important roles in Parkinson’s disease autophagy and lysosomal degradation processes.^55^ *CTSB* and *SMPD1* genetic variants are known to be associated with Parkinson’s disease risk.^55^ In this study, higher CTSB and SMPD1 levels in *LRRK2*+ patients indicate dysregulation of autophagy-endolysosomal pathway and potentially increased macroautophagy.^56,57^ Brain TENM4 is involved in axon guidance and myelination.^58^ In our study, it was significantly reduced in *LRRK2*+ patients. Loss-of-function and missense variants in *TENM4* are associated with early onset Parkinson’s disease and essential tremor, a potential risk factor for developing Parkinson’s disease.^60-59^

### Idiopathic patients

Worldwide, ∼90% of Parkinson’s disease cases are idiopathic. Proteome differences between idiopathic patients and controls comprise not only markers of inflammation, but also of mitochondrial dysfunction / oxidative stress, iron metabolism and other pathological processes (see Table 3). The AK1 kinase is expressed by neurons and astrocytes.^60^ At advanced Parkinson’s disease stages, AK1 is downregulated in the substantia nigra probably due to mitochondrial dysfunction and dopaminergic neuronal death.^60^ The observed elevation of CSF AK1 levels may be associated with Parkinson’s disease progression stages, frontal cortex primary alteration or compensation of altered purine metabolism.^60^ SHANK may be regulated by the mitochondrial kinase PINK1, for which variants are known to be causal for Parkinson’s disease.^61^ It has been reported that knockdown in neurons of *PINK* decreases PSD95 and SHANK1.^61^ SHANK1 was decreased in idiopathic patients suggesting impaired synaptic plasticity. TXN promotes cell proliferation, protection against oxidative stress and anti-apoptotic functions in the brain, which makes it a good candidate for a neurodegeneration marker. Here we find TXN decreased in Parkinson’s disease patients. Iron dysregulation is associated with oxidative stress and lipid peroxidation.^62^ It has been proposed that LPO - heme peroxidase - in the substantia nigra is involved in neurodegeneration.^63^ Lower LPO levels in idiopathic patients found here contrasts with previously reported elevated CSF LPO levels.^63^ The high CSF levels of HAMP could help explaining this discrepancy. HAMP reduces iron accumulation and neuroinflammation by decreasing mitochondrial dysfunction, oxidative stress, and ultimately dopamine neuronal loss.^64^ Moreover, HAMP overexpression – as seen here – promotes alpha-synuclein clearance through autophagy.^65^ It has been also reported that dopamine and levodopa reduce LPO levels.^63^ Given that the idiopathic patients recruited were drug-naïve early Parkinson’s disease patients, dopamine levels in this subpopulation may have helped maintaining low levels of CSF LPO. The glucose metabolism enzyme GPI was elevated in idiopathic patients and may be protective. Its overexpression in dopaminergic neurons protects against alpha-synuclein-induced neurotoxicity.^66^ As seen in *GBA*+ patients, reduced DLK1 levels in idiopathic patients may suggest neurodegeneration. Lower levels of RAB31, a small GTPase involved in exosome biogenesis^67^ and potentially in alpha-synuclein spreading in Parkinson’s disease^68^ were observed, too. RIPK2, a LRRK2 substrate, is lower in idiopathic patients, which matches the fact that LRRK2 deficiency leads to reduced activation of RIPK2.^69^ The neurotrophic factor VEGFA is neuroprotective and has genetic variant associated with Parkinson’s disease risk.^70^ VIP enhances striatal plasticity and prevents dopaminergic cell loss in parkinsonian rats.^71^ VEGFA and VIP at lower levels in idiopathic patients may reflect ongoing neurodegeneration.

### Heterogeneity of idiopathic patients

Disease heterogeneity challenges the development of disease modifying therapies. In this study, we used proteomic data to stratify idiopathic patients into clinically relevant endotypes. The lack of differences in the CSF levels of causal proteins, suggests that endotype molecular differences may be downstream from causal effects and affect the specific characteristics of Parkinson’s disease phenotypes rather than Parkinson’s disease risk. And yet, those molecular differences can affect progression as reflected by differences in ngPD-ProS and might be the basis of personalized therapy approaches.

The endotype robustness was reflected by the identified patient cluster stability (Figure 3B) and the high performance of the endotype predictive model (see Figure 3C). The fact that CSF phospho-tau levels sufficed to predict endotypes, suggests that targeted assessment of CSF proteins may be appropriate for idiopathic patient stratification in a clinical setting.

It is worth noting that, while in Tsukita *et al*. ^10^ the ngPD-ProS perform similarly for both the idiopathic and the combined genetic subcohorts, here we see little overlap in the differentially expressed proteins between the idiopathic and each of the genetic subcohorts separately. This apparent discrepancy could just be reflecting a common CSF proteomic signature rising from the combination of genetic subcohorts even though each genetic subcohort may have different etiology. Supporting this hypothesis, when we compared the proteomes of the combined genetic subcohorts (*GBA+* and *LRRK2+*) with their respective controls (data not shown) we found seven proteins shared with the 24 in the ngPD-ProS (i.e. DLK1, LPO, NEFH, RIPK2, SEMG2, VIP and LRFN2). These seven proteins intersected almost completely with the ten proteins that overlapped between the 24 proteins in the ngPD-ProS and the proteins differentially expressed between idiopathic patients and controls (i.e. DLK1, LPO, NEFH, RIPK2, SEMG2, VIP, CCL14, HAMP, RSPO4, and TXN). This similarity between the idiopathic subcohort and the combination of the genetic subcohorts could help explaining the apparent discrepancy mentioned above.

In summary, we: i) identified causal proteins for Parkinson’s disease, ii) assessed CSF proteome differences in Parkinson’s disease patients of genetic and idiopathic etiology, and. iii) stratified idiopathic patients into robust subtypes. Our findings not only contribute to the identification of new therapeutic targets but also to shaping personalized medicine in CNS neurodegeneration.

## Data Availability

The results of the current study are publicly available on the LONI database and all data produced in the present work are contained in the manuscript

## Abbreviations

AUC: Area Under the Curve
FDR: False Discovery Rate
GWAS: Genome-wide Association Study
LD: Linkage Disequilibrium
MAF: Major Allele Frequency
MDS: Movement Disorder Society
MoCA: Montreal Cognitive Assessment
ngPD-ProS: non-genetic Parkinson’s Disease-associated Proteomic Score
PPMI: Parkinson Progression Marker Initiative
pQTL: Proteomic Quantitative Trait Locus
QC: Quality Control
SOMAmer: Slow Off-rate Modified Aptamer
SNP: Single Nucleotide Polymorphism
UPDRS: Unified Parkinson’s Disease Rating Scale
WGCNA: Weighted Gene Co-expression Network Analysis

## Acknowledgements

The authors would like to thank Rose Case (Indiana University Genetics Biobank, Indiana University School of Medicine, Indianapolis, USA) for her key role in sample management and Myung Shin (Genome and Biomarker Sciences. Merck Exploratory Science Center. Cambridge, USA), Karla Gonzalez (Department of Medical and Molecular Genetics, Indiana University School of Medicine. Indianapolis, USA) and Faraz Faghri (Laboratory of Neurogenetics, National Institute on Aging, National Institutes of Health. Bethesda, USA) for constructive feedback.

## Supplementary material

Supplementary material is available at *Brain* online.

