## Supplementary Material for "Parkinson’s disease causality and heterogeneity: a proteogenomic view"

**Supplementary Table 1 Mendelian randomization using low linkage disequilibrium (LD) threshold**

| Protein | Nr of IVs | Method | β | Nominal *P*-value | Colocalization PP.H4.abf | Horizontal Pleiotropy Test (MRPRESSO *P*-value) |
|---|---|---|---|---|---|---|
| GPNMB[a,b] | 11 | IVW | 0.14 | $1.39 \times 10^{-20}$ | 0.94 | 0.65 |
| FCGR2B[b] | 17 | IVW | 0.07 | $2.43 \times 10^{-16}$ | 0.96 | 0.49 |
| FCGR2A[b] | 18 | IVW | 0.07 | $1.93 \times 10^{-14}$ | 0.95 | 0.26 |
| CTSB[b] | 12 | IVW | −0.11 | $2.19 \times 10^{-12}$ | 0.22 | 0.47 |
| CD38[b] | 1 | Wald ratio | −0.53 | $2.62 \times 10^{-11}$ | 0.45 | NA |
| HP[b] | 19 | IVW | 0.06 | $2.38 \times 10^{-8}$ | 0.01 | 0.60 |
| LTF[b] | 23 | IVW | 0.05 | $2.82 \times 10^{-8}$ | 0.04 | 0.65 |
| HLA-DQA2[b,c] | 33 | IVW | −0.14 | $1.20 \times 10^{-7}$ | 0.03 | 0.00 |
| HAVCR2[b] | 13 | IVW | −0.10 | $4.77 \times 10^{-7}$ | 0.30 | 0.67 |
| CLEC3B[b] | 4 | IVW | −0.16 | $1.94 \times 10^{-6}$ | 0.29 | 0.97 |
| BST1[b,c] | 10 | IVW | 0.12 | $2.63 \times 10^{-6}$ | 0.24 | 0.00 |
| HAPLN1[b] | 15 | IVW | 0.06 | $7.24 \times 10^{-6}$ | 0.05 | 0.79 |
| MANEA[b] | 17 | IVW | 0.05 | $7.26 \times 10^{-6}$ | 0.10 | 0.94 |
| NQO2[b] | 11 | IVW | 0.05 | $7.63 \times 10^{-6}$ | 0.05 | 0.98 |
| ARSA[a,b] | 5 | IVW | 0.13 | $1.70 \times 10^{-5}$ | 0.58 | 0.62 |
| LGALS3[b] | 8 | IVW | 0.07 | $1.86 \times 10^{-5}$ | 0.02 | 0.50 |
| PAM[b] | 12 | IVW | 0.09 | $2.64 \times 10^{-5}$ | 0.08 | 0.47 |
| IL1RL1[b] | 26 | IVW | 0.03 | $2.75 \times 10^{-5}$ | 0.01 | 0.90 |
| TPSAB1[b] | 8 | IVW | −0.06 | $3.22 \times 10^{-5}$ | 0.15 | 0.80 |
| HSP90B1[b] | 22 | IVW | 0.04 | $3.50 \times 10^{-5}$ | 0.03 | 0.19 |
| GLCE[b] | 14 | IVW | 0.05 | $4.84 \times 10^{-5}$ | 0.10 | 0.96 |
| MANSC4[b] | 18 | IVW | 0.05 | $5.20 \times 10^{-5}$ | 0.02 | 0.85 |

| | | | | | | |
|---|---|---|---|---|---|---|
| ICAM1[b] | 8 | IVW | −0.07 | $7.29 \times 10^{-5}$ | 0.32 | 0.74 |
| RABEPK[b] | 10 | IVW | −0.05 | $7.33 \times 10^{-5}$ | 0.10 | 0.86 |
| SIRPB1[b] | 25 | IVW | −0.03 | $9.11 \times 10^{-5}$ | 0.01 | 0.60 |
| CLN5[a,b] | 2 | IVW | −0.15 | $1.19 \times 10^{-4}$ | 0.38 | NA |
| IL9[b] | 12 | IVW | 0.06 | $1.21 \times 10^{-4}$ | 0.05 | 0.96 |
| PCSK7[b] | 18 | IVW | −0.04 | $1.21 \times 10^{-4}$ | 0.02 | 0.98 |
| AGT[b] | 4 | IVW | 0.12 | $1.77 \times 10^{-4}$ | 0.06 | 0.97 |
| CD274[b] | 16 | IVW | 0.08 | $1.96 \times 10^{-4}$ | 0.02 | 0.46 |
| RBP7[b] | 3 | IVW | 0.20 | $2.23 \times 10^{-4}$ | 0.09 | NA |
| C4B[b,c,d] | 101 | IVW | 0.02 | $2.85 \times 10^{-4}$ | 0.00 | 0.00 |
| PLA2G7[b] | 14 | IVW | 0.04 | $2.96 \times 10^{-4}$ | 0.05 | 0.57 |
| C4A[b,c,d] | 101 | IVW | 0.02 | $3.76 \times 10^{-4}$ | 0.00 | 0.00 |
| ASIP[b] | 3 | IVW | −0.13 | $5.13 \times 10^{-4}$ | 0.17 | NA |
| EGF[b] | 3 | IVW | −0.13 | $5.23 \times 10^{-4}$ | 0.11 | NA |
| TPSB2[b] | 10 | IVW | −0.05 | $5.96 \times 10^{-4}$ | 0.15 | 0.27 |
| RNASE3[b] | 6 | IVW | 0.09 | $6.14 \times 10^{-4}$ | 0.01 | 0.26 |
| ACP1[b] | 15 | IVW | 0.04 | $6.25 \times 10^{-4}$ | 0.04 | 0.88 |
| A4GALT[b] | 2 | IVW | −0.21 | $6.62 \times 10^{-4}$ | 0.11 | NA |
| DSCAM[b] | 3 | IVW | −0.30 | $8.78 \times 10^{-4}$ | 0.19 | NA |
| LCT[b,c] | 25 | IVW | 0.04 | $8.91 \times 10^{-4}$ | 0.00 | 0.00 |
| COLEC11[b] | 8 | IVW | −0.05 | $9.58 \times 10^{-4}$ | 0.03 | 0.61 |
| SPOCK2[b] | 8 | IVW | 0.08 | 0.0010 | 0.03 | 0.98 |
| VWA2[b] | 5 | IVW | −0.11 | 0.0011 | 0.04 | 0.76 |
| ADAMTS4[b] | 8 | IVW | −0.06 | 0.0013 | 0.19 | 0.36 |
| RNASE2[b] | 10 | IVW | 0.06 | 0.0013 | 0.02 | 0.61 |
| PRTN3[b] | 2 | IVW | 0.17 | 0.0013 | 0.06 | NA |
| RPN1[b] | 10 | IVW | 0.08 | 0.0013 | 0.02 | 0.57 |

| | | | | | | |
|---|---|---|---|---|---|---|
| PDCD1LG2[b] | 12 | IVW | −0.09 | 0.0013 | 0.04 | 0.07 |
| ADGRE2[b] | 10 | IVW | −0.05 | 0.0013 | 0.01 | 0.87 |
| MPIG6B[b] | 18 | IVW | −0.05 | 0.0014 | 0.00 | 0.18 |
| TAPBPL[b] | 10 | IVW | −0.03 | 0.0014 | 0.03 | 0.50 |
| SIGLEC9[b] | 19 | IVW | 0.03 | 0.0015 | 0.02 | 0.99 |
| LRP12[a,b] | 1 | Wald ratio | 0.73 | 0.0017 | 0.66 | NA |
| DNAJC30[b] | 3 | IVW | −0.09 | 0.0018 | 0.03 | NA |
| CCL15[b] | 3 | IVW | −0.08 | 0.0019 | 0.12 | NA |
| VTN[b] | 22 | IVW | −0.03 | 0.0022 | 0.01 | 0.75 |
| NUCB1[b] | 3 | IVW | −0.12 | 0.0023 | 0.05 | NA |
| TRH[b] | 4 | IVW | −0.08 | 0.0023 | 0.07 | 0.75 |
| POSTN[b] | 8 | IVW | −0.07 | 0.0028 | 0.02 | 0.76 |
| PLXNB2[a,b] | 36 | IVW | −0.02 | 0.0029 | 0.08 | 0.09 |
| IL18R1[b] | 23 | IVW | 0.03 | 0.0029 | 0.01 | 0.79 |
| IDUA[b] | 3 | IVW | 0.10 | 0.0033 | 0.00 | NA |
| CFD[b] | 10 | IVW | −0.05 | 0.0035 | 0.01 | 0.58 |
| GGH[a,b] | 4 | IVW | 0.08 | 0.0035 | 0.02 | 0.49 |
| FGFRL1[b] | 5 | IVW | 0.13 | 0.0041 | 0.00 | 0.07 |
| LMAN2L[b] | 3 | IVW | −0.15 | 0.0045 | 0.03 | NA |
| TXNDC15 | 6 | IVW | −0.05 | 0.0048 | 0.12 | 0.46 |
| KNG1 | 6 | IVW | 0.05 | 0.0050 | 0.03 | 0.80 |
| EPHA1 | 6 | IVW | 0.05 | 0.0050 | 0.08 | 0.98 |
| CCL23 | 2 | IVW | 0.10 | 0.0054 | 0.03 | NA |
| JAML | 13 | IVW | −0.02 | 0.0055 | 0.03 | 0.86 |
| CLEC7A | 5 | IVW | 0.05 | 0.0059 | 0.09 | 0.87 |
| HRG | 2 | IVW | −0.08 | 0.0060 | 0.09 | NA |
| NUDT9 | 4 | IVW | −0.08 | 0.0070 | 0.03 | 0.87 |

| | | | | | | |
|---|---|---|---|---|---|---|
| CHI3L2 | 11 | IVW | −0.04 | 0.0072 | 0.04 | 0.45 |
| OAS1 | 5 | IVW | 0.08 | 0.0080 | 0.02 | 0.41 |
| TNFRSF11A | 12 | IVW | −0.06 | 0.0082 | 0.02 | 0.99 |
| NDNF | 4 | IVW | −0.10 | 0.0083 | 0.01 | 0.32 |
| INPP5B | 6 | IVW | −0.08 | 0.0096 | 0.01 | 0.43 |
| GRN[a] | 1 | Wald ratio | −0.19 | 0.0097 | 0.22 | NA |
| WFIKKN2 | 10 | IVW | 0.03 | 0.010 | 0.00 | 0.70 |
| ROR2 | 4 | IVW | 0.13 | 0.010 | 0.01 | 0.42 |
| H6PD | 6 | IVW | −0.07 | 0.010 | 0.05 | 0.92 |
| GFRAL | 8 | IVW | 0.05 | 0.012 | 0.04 | 0.97 |
| BTN2A1 | 12 | IVW | 0.07 | 0.013 | 0.00 | 0.19 |
| MMP2 | 5 | IVW | −0.08 | 0.014 | 0.03 | 0.96 |
| PGD | 2 | IVW | 0.23 | 0.014 | 0.48 | NA |
| NPW[c] | 7 | IVW | 0.09 | 0.014 | 0.00 | 0.02 |
| AMY2B | 2 | IVW | 0.19 | 0.015 | 0.03 | NA |
| IL6R | 15 | IVW | 0.03 | 0.016 | 0.00 | 0.09 |
| GSN | 6 | IVW | −0.06 | 0.016 | 0.03 | 0.98 |
| TBL2 | 4 | IVW | −0.07 | 0.016 | 0.08 | 0.98 |
| TMEM9 | 18 | IVW | −0.04 | 0.018 | 0.01 | 0.43 |
| LAMC2 | 11 | IVW | −0.03 | 0.018 | 0.04 | 0.58 |
| TDGF1[c] | 19 | IVW | −0.03 | 0.019 | 0.00 | 0.01 |
| PPIE | 7 | IVW | 0.07 | 0.020 | 0.03 | 0.96 |
| CCN3 | 3 | IVW | −0.09 | 0.020 | 0.03 | NA |
| EFEMP1 | 13 | IVW | 0.05 | 0.020 | 0.01 | 0.42 |
| B3GLCT | 3 | IVW | −0.09 | 0.020 | 0.03 | NA |
| ESD | 3 | IVW | −0.04 | 0.021 | 0.09 | NA |
| TNFRSF14 | 9 | IVW | −0.07 | 0.021 | 0.03 | 0.80 |

| | | | | | | |
|---|---|---|---|---|---|---|
| ACP5 | 6 | IVW | 0.05 | 0.021 | 0.04 | 0.88 |
| TEK | 9 | IVW | −0.05 | 0.021 | 0.04 | 0.71 |
| SEMA3E | 9 | IVW | −0.04 | 0.022 | 0.02 | 0.90 |
| WARS1 | 2 | IVW | 0.12 | 0.022 | 0.19 | NA |
| TIE1 | 1 | Wald ratio | −0.18 | 0.022 | 0.09 | NA |
| QPCT | 6 | IVW | 0.06 | 0.023 | 0.03 | 0.51 |
| FABP6[ac] | 9 | IVW | 0.08 | 0.024 | 0.01 | 0.01 |
| CNTN2 | 9 | IVW | 0.05 | 0.025 | 0.00 | 0.26 |
| PPT1 | 6 | IVW | 0.05 | 0.026 | 0.01 | 0.76 |
| MBL2 | 10 | IVW | 0.04 | 0.027 | 0.02 | 0.66 |
| CPA4 | 11 | IVW | −0.03 | 0.027 | 0.01 | 0.18 |
| LY86 | 3 | IVW | −0.08 | 0.027 | 0.03 | NA |
| CCL16 | 2 | IVW | 0.05 | 0.028 | 0.06 | NA |
| MCEE | 5 | IVW | −0.05 | 0.031 | 0.01 | 1.00 |
| IFNAR1 | 6 | IVW | −0.05 | 0.031 | 0.02 | 0.55 |
| NQO1 | 14 | IVW | 0.03 | 0.032 | 0.03 | 0.18 |
| ICAM4 | 1 | Wald ratio | 0.32 | 0.033 | 0.09 | NA |
| TRIL | 2 | IVW | −0.08 | 0.034 | 0.03 | NA |
| ADAMTS5 | 12 | IVW | −0.03 | 0.034 | 0.01 | 0.69 |
| THBS3 | 2 | IVW | 0.08 | 0.034 | 0.00 | NA |
| BTN3A1 | 24 | IVW | −0.02 | 0.035 | 0.00 | 0.83 |
| MXRA7 | 6 | IVW | −0.12 | 0.037 | 0.10 | 0.18 |
| TNFRSF10B | 5 | IVW | 0.09 | 0.038 | 0.02 | 0.24 |
| FKBP7 | 12 | IVW | 0.04 | 0.038 | 0.01 | 0.95 |
| AKR1B1 | 1 | Wald ratio | 0.23 | 0.039 | 0.06 | NA |
| TAPBP | 4 | IVW | −0.07 | 0.039 | 0.00 | 0.62 |
| BCHE | 2 | IVW | 0.12 | 0.040 | 0.02 | NA |

| | | | | | | |
|---|---|---|---|---|---|---|
| RSPO4[a] | 1 | Wald ratio | 0.15 | 0.041 | 0.06 | NA |
| CST5 | 11 | IVW | −0.04 | 0.041 | 0.02 | 0.15 |
| MAX | 1 | Wald ratio | 0.17 | 0.042 | 0.05 | NA |
| IL15RA | 3 | IVW | −0.08 | 0.045 | 0.02 | NA |
| INSR | 2 | IVW | 0.24 | 0.045 | 0.07 | NA |
| IL10RB | 6 | IVW | −0.04 | 0.047 | 0.01 | 0.92 |
| LILRA4 | 6 | IVW | −0.07 | 0.048 | 0.02 | 0.21 |
| FAH | 2 | IVW | 0.11 | 0.048 | 0.03 | NA |
| B3GNT8 | 5 | IVW | −0.03 | 0.049 | 0.02 | 0.99 |
| F11 | 3 | IVW | −0.08 | 0.049 | 0.02 | NA |
| SIGLEC7 | 7 | IVW | 0.03 | 0.050 | 0.03 | 0.99 |
| MAN2B2 | 6 | IVW | −0.02 | 0.050 | 0.01 | 0.65 |

[a] PD protein marker in Supplementary Table 3
[b] passed FDR correction
[c] corrected by removing outlier by MRPRESSO
[d] has homolog detected by the same SOMAmer

MAF>0.05, cis-pQTL p<5e-08, F-statistic>10, clumped by r2<0.3 for SNPs within 1000kb region
744 proteins have a significant cis-pQTL

**Supplementary Table 2 Mendelian randomization using high LD threshold**

| Protein | Nr of IVs | Method | β | Nominal *P*-value | Colocalization PP.H4.abf |
|---|---|---|---|---|---|
| CD38[a] | 1 | Wald ratio | −0.53 | $2.62 \times 10^{-11}$ | 0.45 |
| GPNMB[a,b] | 1 | Wald ratio | 0.15 | $1.47 \times 10^{-8}$ | 0.94 |
| HLA-DQA2[a] | 2 | IVW | −0.21 | $1.93 \times 10^{-7}$ | 0.03 |
| FCGR2A[a] | 1 | Wald ratio | 0.06 | $4.37 \times 10^{-5}$ | 0.95 |
| ASIP | 2 | IVW | −0.14 | $7.99 \times 10^{-4}$ | 0.17 |
| CTSB | 1 | Wald ratio | −0.10 | 0.0011 | 0.20 |
| ARSA[b] | 1 | Wald ratio | 0.15 | 0.0012 | 0.58 |
| TPSB2 | 2 | IVW | −0.06 | 0.0015 | 0.15 |
| NCR1 | 1 | Wald ratio | 0.19 | 0.0015 | 0.39 |
| LRP12[b] | 1 | Wald ratio | 0.73 | 0.0017 | 0.66 |
| HAVCR2 | 2 | IVW | −0.09 | 0.0027 | 0.30 |
| CLN5[b] | 1 | Wald ratio | −0.13 | 0.0033 | 0.38 |
| CLEC3B | 1 | Wald ratio | −0.15 | 0.0035 | 0.29 |
| ICAM1 | 1 | Wald ratio | −0.07 | 0.0037 | 0.32 |
| DSCAM | 1 | Wald ratio | −0.22 | 0.0044 | 0.19 |
| PGD | 1 | Wald ratio | 0.17 | 0.0052 | 0.48 |
| ADAMTS4 | 1 | Wald ratio | −0.07 | 0.0058 | 0.19 |
| WARS1 | 1 | Wald ratio | 0.16 | 0.0060 | 0.19 |
| FGFRL1 | 1 | Wald ratio | 0.14 | 0.0071 | 0.00 |
| GRN[b] | 1 | Wald ratio | −0.19 | 0.0097 | 0.22 |
| EPHA1 | 2 | IVW | 0.06 | 0.010 | 0.08 |
| SIGLEC6 | 1 | Wald ratio | 0.27 | 0.014 | 0.19 |
| AGT | 1 | Wald ratio | 0.14 | 0.015 | 0.06 |
| PAM | 1 | Wald ratio | 0.09 | 0.015 | 0.08 |
| LTF | 1 | Wald ratio | 0.04 | 0.016 | 0.04 |
| PLXNB2[b] | 1 | Wald ratio | −0.03 | 0.019 | 0.08 |
| A4GALT | 1 | Wald ratio | −0.17 | 0.019 | 0.11 |
| TXNDC15 | 1 | Wald ratio | −0.05 | 0.020 | 0.12 |
| EGF | 1 | Wald ratio | −0.11 | 0.021 | 0.11 |
| HRG | 1 | Wald ratio | −0.09 | 0.021 | 0.09 |
| TIE1 | 1 | Wald ratio | −0.18 | 0.022 | 0.09 |

| | | | | | |
|---|---|---|---|---|---|
| BST1 | 2 | IVW | 0.10 | 0.023 | 0.24 |
| RABEPK | 1 | Wald ratio | −0.05 | 0.023 | 0.10 |
| MXRA7 | 1 | Wald ratio | −0.14 | 0.024 | 0.10 |
| GLCE | 1 | Wald ratio | 0.04 | 0.025 | 0.10 |
| S100A7 | 1 | Wald ratio | 0.08 | 0.026 | 0.16 |
| ESD | 1 | Wald ratio | −0.05 | 0.030 | 0.09 |
| CCN3 | 2 | IVW | −0.09 | 0.032 | 0.03 |
| MANEA | 3 | IVW | 0.04 | 0.032 | 0.10 |
| ICAM4 | 1 | Wald ratio | 0.32 | 0.033 | 0.09 |
| FBP2 | 1 | Wald ratio | −0.17 | 0.035 | 0.05 |
| CCL15 | 1 | Wald ratio | −0.07 | 0.037 | 0.12 |
| PLA2G7 | 1 | Wald ratio | 0.05 | 0.039 | 0.05 |
| AKR1B1 | 1 | Wald ratio | 0.23 | 0.039 | 0.06 |
| IDUA | 1 | Wald ratio | 0.05 | 0.039 | 0.00 |
| HP | 2 | IVW | 0.05 | 0.039 | 0.01 |
| C1QL1 | 2 | IVW | 0.10 | 0.040 | 0.00 |
| CCL23 | 2 | IVW | 0.08 | 0.040 | 0.03 |
| RSPO4[b] | 1 | Wald ratio | 0.15 | 0.041 | 0.06 |
| MAX | 1 | Wald ratio | 0.17 | 0.042 | 0.05 |
| ENTPD6 | 2 | IVW | −0.15 | 0.044 | 0.11 |
| CLEC7A | 1 | Wald ratio | 0.04 | 0.045 | 0.09 |
| INSR | 2 | IVW | 0.24 | 0.045 | 0.07 |
| TMPRSS5 | 1 | Wald ratio | −0.07 | 0.047 | 0.06 |
| H6PD | 1 | Wald ratio | −0.08 | 0.049 | 0.05 |

[a] passed FDR correction
[b] PD protein marker in Supplementary Table 3

MAF>0.05, cis-pQTL p<5e-08, F-statistic>10, clumped by r2<0.01 for SNPs within 1000kb region
744 proteins have a significant cis-pQTL

**Supplementary Table 3 Significant differences at proteome level between PD patients (all) and controls (all)**

| Gene symbol | PD change direction | *P*-value (FDR adjusted) |
|---|---|---|
| LPO | - | $2.35 \times 10^{-08}$ |
| SEMG2 | - | $9.67 \times 10^{-08}$ |
| DLK1 | - | $1.76 \times 10^{-07}$ |
| RIPK2 | - | $3.91 \times 10^{-07}$ |
| AMP | + | $4.35 \times 10^{-05}$ |
| NETO1 | - | $1.02 \times 10^{-04}$ |
| PLXNB2 | + | $4.62 \times 10^{-04}$ |
| BRICD5 | - | $1.05 \times 10^{-03}$ |
| ADM | - | $1.05 \times 10^{-03}$ |
| UNC5D | - | $1.05 \times 10^{-03}$ |
| SCUBE1 | + | $1.05 \times 10^{-03}$ |
| CTSO | + | $1.05 \times 10^{-03}$ |
| VEGFA | - | $1.19 \times 10^{-03}$ |
| DDR1 | + | $1.28 \times 10^{-03}$ |
| GPI | + | $1.64 \times 10^{-03}$ |
| VWC2L | - | $1.64 \times 10^{-03}$ |
| CNTFR | + | $1.67 \times 10^{-03}$ |
| TMEM106A | + | $1.68 \times 10^{-03}$ |
| MDK | + | $1.68 \times 10^{-03}$ |
| VIP | - | $1.76 \times 10^{-03}$ |
| ETS2 | + | $2.16 \times 10^{-03}$ |
| SHANK1 | - | $2.16 \times 10^{-03}$ |
| PTPRR | - | $2.20 \times 10^{-03}$ |
| MAN1C1 | + | $2.63 \times 10^{-03}$ |
| SEMA6A | + | $2.63 \times 10^{-03}$ |
| HS6ST1 | + | $3.66 \times 10^{-03}$ |
| TENM4 | - | $5.51 \times 10^{-03}$ |
| FRZB | + | $7.48 \times 10^{-03}$ |
| ARSA | + | $7.58 \times 10^{-03}$ |
| DPP7 | + | $7.71 \times 10^{-03}$ |
| FAM171B | + | $7.80 \times 10^{-03}$ |
| CREG1 | + | $7.80 \times 10^{-03}$ |
| OCRL | - | $8.08 \times 10^{-03}$ |
| CNTN1 | + | $8.08 \times 10^{-03}$ |
| PRDX3 | + | $8.08 \times 10^{-03}$ |
| DPYSL5 | + | $8.70 \times 10^{-03}$ |
| EPHA5 | - | $8.70 \times 10^{-03}$ |
| IL17D | + | $8.70 \times 10^{-03}$ |
| CHST5 | + | $8.70 \times 10^{-03}$ |
| FZD10 | - | $8.83 \times 10^{-03}$ |
| ARTN | - | $9.03 \times 10^{-03}$ |
| EFNB3 | - | $9.05 \times 10^{-03}$ |
| SMIM24 | - | 0.0101 |
| NPTX2 | - | 0.0106 |
| LRP12 | - | 0.0106 |
| CD109 | + | 0.0112 |
| IFNGR1 | + | 0.0116 |
| CDH1 | + | 0.0117 |
| GCH1 | + | 0.0121 |
| CCL14 | + | 0.0121 |
| AK1 | + | 0.0121 |
| SPINK9 | - | 0.0121 |
| GXYLT1 | + | 0.0121 |

| | | |
|---|---|---|
| NEFH | + | 0.0121 |
| HGF | + | 0.0147 |
| ATP1B2 | + | 0.0166 |
| TFPI | + | 0.0169 |
| FTH1 / FTL | + | 0.0169 |
| MAPK1 | + | 0.0180 |
| TREM2 | + | 0.0180 |
| GPNMB | + | 0.0185 |
| MEGF10 | - | 0.0187 |
| CPE | + | 0.0187 |
| NIPAL4 | + | 0.0201 |
| TOPBP1 | - | 0.0201 |
| PLOD3 | + | 0.0220 |
| CLN5 | + | 0.0227 |
| RSPO4 | - | 0.0228 |
| CDCP1 | + | 0.0258 |
| PTN | + | 0.0261 |
| OMD | + | 0.0261 |
| NFASC | + | 0.0261 |
| SMPD1 | + | 0.0274 |
| MMP8 | - | 0.0274 |
| CHST6 | + | 0.0274 |
| CEL | + | 0.0278 |
| GNS | + | 0.0304 |
| C1QTNF1 | + | 0.0311 |
| CA10 | + | 0.0325 |
| IL10 | - | 0.0325 |
| CD209 | - | 0.0325 |
| ACP2 | + | 0.0325 |
| ROBO3 | - | 0.0331 |
| RAF1 | - | 0.0354 |
| SLITRK3 | + | 0.0359 |
| PCSK1 | - | 0.0359 |
| CTSC | + | 0.0359 |
| CECR1 | + | 0.0359 |
| NDUFB4 | - | 0.0363 |
| SERPINB1 | + | 0.0363 |
| TGFBR3 | + | 0.0363 |
| IGF1R | + | 0.0363 |
| GZMB | + | 0.0365 |
| ADAM9 | + | 0.0365 |
| ADAM22 | + | 0.0370 |
| AP2A2 | - | 0.0381 |
| GRN | + | 0.0381 |
| ADCYAP1 | - | 0.0383 |
| CD44 | - | 0.0390 |
| CLMP | + | 0.0390 |
| RELT | - | 0.0393 |
| MATN2 | + | 0.0402 |
| CD2 | + | 0.0402 |
| PCDH9 | + | 0.0402 |
| B3GALT2 | - | 0.0402 |
| PSMB5 | + | 0.0402 |
| DNAJB11 | - | 0.0402 |
| KIAA1549L | + | 0.0426 |
| GGH | + | 0.0437 |

| | | |
|---|---|---|
| BCAN | + | 0.0454 |
| SIAE | + | 0.0454 |
| GRAMD1C | - | 0.0455 |
| LIFR | + | 0.0465 |
| GRB2 | - | 0.0470 |
| TRAPPC4 | + | 0.0470 |
| SMURF1 | - | 0.0473 |
| CTSH | + | 0.0473 |
| EPHB6 | - | 0.0490 |
| LARGE1 | + | 0.0490 |
| LYVE1 | - | 0.0494 |
| BTC | + | 0.0494 |

The change in PD represents (+) increased and (-) decreased in Parkinson's disease patients vs controls, respectively. Note**:** there are 122 proteins tagged by 129 SOMAmers. Duplicated SOMAmers are removed from the table for clarity (the one with the lowest *P*-value is kept). No discrepancies in directionality were present for duplicated SOMAmers.

**Supplementary Table 4 Significant differences at proteome level between endotypes 1 and 2 of the Parkinson's disease idiopathic subcohort and healthy controls**

| Gene symbol | PD change direction | *P*-value (FDR adjusted) |
|---|---|---|
| **Endotype 1 vs HC** | | |
| CNTFR | + | 0.0178 |
| LPO | - | $5 \times 10^{-06}$ |
| MMP10 | - | 0.0178 |
| RIPK2 | - | 0.0178 |
| VEGFA | - | 0.0378 |
| **Endotype 2 vs HC** | | |
| ACVR2B | + | 0.0302 |
| ADAM10 | - | 0.0289 |
| ADGRF5 | - | 0.0113 |
| ADH4 | - | 0.0260 |
| ADI1 | - | 0.0087 |
| ADM | - | 0.0152 |
| AGA | - | 0.0322 |
| AK1 | + | 0.0039 |
| ANGPTL7 | - | 0.0128 |
| ANTXR1 | - | 0.0443 |
| APBB2 | - | 0.0390 |
| ARHGAP30 | - | 0.0184 |
| ASCC1 | - | 0.0336 |
| ATRAID | - | 0.0461 |
| BCAN | + | 0.0266 |
| BHMT2 | - | 0.0185 |
| BMPR1A | + | 0.0152 |
| BMPR1B | + | 0.0282 |
| BRICD5 | - | 0.0268 |
| BTC | + | 0.0328 |
| C1RL | - | 0.0378 |
| C4BPB | - | 0.0398 |
| CACYBP | - | 0.0253 |
| CALB1 | - | 0.0016 |
| CBLN4 | - | 0.0339 |
| CBS | - | 0.0113 |
| CCL14 | + | 0.0184 |
| CCL26 | + | 0.0113 |
| CD109 | + | 0.0103 |
| CDC42BPA | - | 0.0311 |
| CDH1 | + | 0.0113 |
| CELA3B | - | 0.0111 |
| CES1 | - | 0.0103 |
| CHFR | - | 0.0367 |
| CLPSL2 | + | 0.0291 |
| COL15A1 | + | 0.0087 |
| COMP | - | 0.0185 |
| COX4I2 | + | 0.0291 |
| CPE | + | 0.0339 |
| CRIM1 | + | 0.0039 |
| CRTAC1 | - | 0.0122 |
| CYCS | - | 0.0157 |

| | | |
|---|---|---|
| DAG1 | + | 0.0121 |
| DCTN2 | - | 0.0151 |
| DEFA5 | - | 0.0444 |
| DGKB | - | 0.0121 |
| DLK2 | - | 0.0339 |
| DNER | + | 0.0268 |
| DUSP6 | - | 0.0256 |
| EDA | + | 0.0493 |
| EFNB3 | - | 0.0260 |
| EFS | - | 0.0303 |
| EMID1 | + | 0.0183 |
| ENTPD5 | - | 0.0339 |
| EPHA3 | - | 0.0311 |
| EPHA4 | - | 0.0157 |
| EPHA5 | - | 0.0087 |
| EPHB6 | - | 0.0468 |
| FAF2 | + | 0.0363 |
| FAP | - | 0.0113 |
| FCN3 | - | 0.0039 |
| FGF12 | - | 0.0215 |
| FGF7 | + | 0.0495 |
| FGL1 | - | 0.0435 |
| FRZB | + | 0.0103 |
| GABBR1 | - | 0.0089 |
| GDF11 | - | 0.0443 |
| GFRA2 | - | 0.0378 |
| GP1BA | - | 0.0296 |
| GPI | + | 0.0039 |
| GRB2 | - | 0.0269 |
| GSTA1 | + | 0.0138 |
| HAMP | + | 0.0033 |
| HAPLN1 | - | 0.0067 |
| HCAR2 | - | 0.0113 |
| HEPACAM2 | - | 0.0450 |
| HGF | + | 0.0260 |
| HIKESHI | + | 0.0432 |
| HLA-DMA | - | 0.0311 |
| HPGD | - | 0.0303 |
| HS6ST1 | + | 0.0334 |
| ICAM2 | - | 0.0141 |
| IGFBP2 | + | 0.0113 |
| IGFBPL1 | - | 0.0172 |
| IGLL1 | - | 0.0289 |
| IL17D | + | 0.0157 |
| IL19 | - | 0.0387 |
| IL27RA | - | 0.0018 |
| ITSN1 | - | 0.0138 |
| JAG1 | - | 0.0039 |
| KAT6A | + | 0.0157 |
| KERA | + | 0.0103 |
| LIPN | - | 0.0435 |
| LPO | - | 0.0068 |
| LRFN2 | - | 0.0071 |
| LRP12 | - | 0.0138 |
| LYVE1 | - | 0.0432 |
| MAN1B1 | - | 0.0119 |

| | | |
|---|---|---|
| MAN1C1 | + | 0.0131 |
| MATN2 | + | 0.0032 |
| MDK | + | 0.0087 |
| MEGF10 | - | 0.0151 |
| MFNG | - | 0.0458 |
| MINOS1 | - | 0.0496 |
| MLEC | + | 0.0183 |
| MMP8 | - | 0.0243 |
| MPZ | + | 0.0493 |
| MST1R | + | 0.0067 |
| NEFH | + | 0.0222 |
| NELL2 | + | 0.0241 |
| NETO1 | - | 0.0021 |
| NPTN | - | 0.0157 |
| NPTX2 | - | 0.0113 |
| NRXN1 | - | 0.0113 |
| NXPH3 | - | 0.0151 |
| OCIAD1 | - | 0.0303 |
| OCRL | - | 0.0154 |
| OMD | + | 0.0115 |
| OXT | + | 0.0112 |
| PAPPA2 | + | 0.0311 |
| PGD | + | 0.0260 |
| PKN1 | - | 0.0289 |
| PLA2G2C | + | 0.0113 |
| PLOD3 | + | 0.0087 |
| PLXNA1 | - | 0.0087 |
| PPA1 | - | 0.0089 |
| PRDM4 | - | 0.0346 |
| PRG2 | - | 0.0183 |
| PRRG4 | - | 0.0379 |
| PSMD5 | - | 0.0244 |
| PTPRR | - | 0.0036 |
| PZP | - | 0.0328 |
| RAB31 | - | 0.0447 |
| RAB6B | - | 0.0121 |
| RAF1 | - | 0.0103 |
| RAPGEF5 | + | 0.0468 |
| RB1 | + | 0.0121 |
| RBL2 | - | 0.0333 |
| RBP4 | - | 0.0291 |
| RELT | - | 0.0334 |
| RIPK2 | - | 0.0014 |
| RNASE4 | + | 0.0138 |
| ROBO3 | - | 0.0089 |
| ROR1 | - | 0.0336 |
| RSPO4 | - | 0.0445 |
| SCARA5 | - | 0.0499 |
| SCG2 | - | 0.0119 |
| SCUBE1 | + | 0.0005 |
| SERPINA10 | - | 0.0151 |
| SERPINA9 | - | 0.0014 |
| SFTA2 | - | 0.0087 |
| SHANK1 | - | 0.0018 |
| SHANK3 | - | 0.0224 |
| SMIM24 | - | 0.0103 |

| | | |
|---|---|---|
| SMURF1 | - | 0.0291 |
| SOD3 | + | 0.0218 |
| SPINK9 | - | 0.0138 |
| SPOCK3 | + | 0.0131 |
| SPSB1 | + | 0.0497 |
| SRXN1 | - | 0.0113 |
| ST6GALNAC6 | + | 0.0260 |
| STMN3 | - | 0.0157 |
| STX10 | - | 0.0014 |
| STX12 | - | 0.0103 |
| STX1A | - | 0.0039 |
| STX1B | - | 0.0398 |
| STX2 | - | 0.0291 |
| STX3 | - | 0.0322 |
| STX7 | - | 0.0132 |
| SV2A | - | 0.0456 |
| TCN2 | + | 0.0339 |
| TESC | - | 0.0365 |
| TFPI | + | 0.0164 |
| TFRC | + | 0.0157 |
| TGFB3 | - | 0.0390 |
| THBS2 | + | 0.0039 |
| TMEM132B | - | 0.0443 |
| TMEM132D | - | 0.0157 |
| TMEM230 | - | 0.0260 |
| TMEM8B | - | 0.0486 |
| TNFRSF11B | + | 0.0121 |
| TNFRSF13C | - | 0.0339 |
| TNFRSF18 | - | 0.0260 |
| TNFRSF1A | + | 0.0289 |
| TPO | + | 0.0289 |
| TPSG1 | + | 0.0205 |
| TYMP | - | 0.0138 |
| UBL4A | - | 0.0113 |
| UNC5D | - | 0.0121 |
| USE1 | - | 0.0113 |
| VEGFA | - | 0.0301 |
| VIP | - | 0.0103 |
| VWC2 | - | 0.0260 |
| VWC2L | - | 0.0028 |
| WFDC13 | + | 0.0483 |
| YWHAG | - | 0.0117 |
| YWHAZ, YWHAE, YWHAG, YWHAH, SFN, YWHAQ, YWHAB | - | 0.0486 |
| ZNF10 | + | 0.0339 |

**Endotype 1 vs Endotype 2**

| | | |
|---|---|---|
| ADAM10 | + | 0.0141 |
| ADAM23 | + | 0.0106 |
| ADAMTSL2 | + | 0.0210 |
| ADGRF5 | + | 0.0497 |
| ADM | + | 0.0480 |
| AGA | + | 0.0325 |
| AKT2 | - | 0.0323 |

| | | |
|---|---|---|
| ANGPTL7 | + | 0.0088 |
| APBB2 | + | 0.0462 |
| ASIC4 | - | 0.0277 |
| ATP5B | + | 0.0465 |
| AXIN2 | - | 0.0497 |
| B4GALT7 | - | 0.0348 |
| BAG4 | + | 0.0235 |
| BDNF | + | 0.0325 |
| BMPR1B | - | 0.0135 |
| BMPR2 | + | 0.0274 |
| BRD2 | - | 0.0421 |
| C1QBP | + | 0.0385 |
| C4BPB | + | 0.0480 |
| CA4 | + | 0.0323 |
| CA5A | + | 0.0309 |
| CACYBP | + | 0.0403 |
| CALB1 | + | 0.0305 |
| CAPN2 | + | 0.0462 |
| CASS4 | + | 0.0309 |
| CCL26 | - | 0.0262 |
| CD300E | + | 0.0467 |
| CD47 | + | 0.0048 |
| CD96 | + | 0.0130 |
| CDC42BPA | + | 0.0391 |
| CELA3B | + | 0.0106 |
| CES1 | + | 0.0048 |
| CHFR | + | 0.0049 |
| CHP1 | + | 0.0323 |
| CKS1B | + | 0.0355 |
| CLCA1 | + | 0.0411 |
| CLEC2L | + | 0.0309 |
| CNTFR | + | 0.0231 |
| COX4I2 | - | 0.0176 |
| CRB1 | - | 0.0480 |
| CRIM1 | - | 0.0106 |
| CTSH | + | 0.0412 |
| CUL4B | + | 0.0160 |
| CXCL2, CXCL3 | + | 0.0325 |
| CYB5R3 | + | 0.0106 |
| CYP3A4 | + | 0.0480 |
| DCTN2 | + | 0.0323 |
| DGKB | + | 0.0274 |
| DLL4 | + | 0.0409 |
| DNAJB12 | - | 0.0323 |
| DUSP26 | + | 0.0326 |
| DUSP6 | + | 0.0298 |
| EIF4G3 | + | 0.0391 |
| EMID1 | - | 0.0256 |
| FAP | + | 0.0210 |
| FBLN5 | - | 0.0325 |
| FCN3 | + | 0.0336 |
| FJX1 | - | 0.0374 |
| FLRT2 | - | 0.0334 |
| FSTL1 | - | 0.0467 |
| GABBR1 | + | 0.0088 |
| GALNT11 | - | 0.0325 |

| | | |
|---|---|---|
| GP1BA | + | 0.0336 |
| GPD1 | + | 0.0274 |
| GRB14 | + | 0.0323 |
| HCAR2 | + | 0.0394 |
| HLA-DMA | + | 0.0277 |
| HPGD | + | 0.0391 |
| ICAM2 | + | 0.0105 |
| IFNG | + | 0.0351 |
| IFNGR2 | + | 0.0176 |
| IGFBPL1 | + | 0.0274 |
| IGLL1 | + | 0.0296 |
| IL1B | - | 0.0088 |
| IL22 | + | 0.0323 |
| IL27RA | + | 0.0106 |
| ILF3 | + | 0.0482 |
| ITSN1 | + | 0.0041 |
| KAT6A | - | 0.0323 |
| KEAP1 | + | 0.0325 |
| KIRREL3 | + | 0.0412 |
| KLRC4 | + | 0.0391 |
| LECT1 | + | 0.0391 |
| LIPN | + | 0.0325 |
| LRFN2 | + | 0.0374 |
| LTBP4 | - | 0.0355 |
| LYG1 | + | 0.0467 |
| MADCAM1 | + | 0.0465 |
| MAN1B1 | + | 0.0355 |
| MDK | - | 0.0467 |
| MINOS1 | + | 0.0176 |
| MRM3 | + | 0.0306 |
| MST1R | - | 0.0268 |
| MTMR1 | - | 0.0262 |
| MYRF | - | 0.0339 |
| NAT14 | + | 0.0480 |
| NDUFV2 | + | 0.0305 |
| NEGR1 | + | 0.0235 |
| NPTN | + | 0.0048 |
| NRXN1 | + | 0.0325 |
| NTM | + | 0.0274 |
| NXPH3 | + | 0.0202 |
| PAPPA2 | - | 0.0176 |
| PELO | + | 0.0296 |
| PLOD3 | - | 0.0165 |
| PPA1 | + | 0.0088 |
| PPBP | + | 0.0480 |
| PRDM4 | + | 0.0157 |
| PRG2 | + | 0.0348 |
| PYDC1 | + | 0.0410 |
| PZP | + | 0.0456 |
| RB1 | - | 0.0295 |
| RBBP6 | + | 0.0480 |
| RBL2 | + | 0.0497 |
| RNF8 | + | 0.0262 |
| ROBO2 | + | 0.0482 |
| ROR2 | - | 0.0262 |
| RYK | + | 0.0391 |

| | | |
|---|---|---|
| SCARA5 | + | 0.0467 |
| SCARB2 | + | 0.0391 |
| SCG2 | + | 0.0325 |
| SCN2B | + | 0.0165 |
| SCUBE1 | - | 0.0130 |
| SERPINA10 | + | 0.0480 |
| SERPINA9 | + | 0.0106 |
| SGF29 | + | 0.0355 |
| SLC14A2 | + | 0.0119 |
| SOD2 | + | 0.0277 |
| SPOCK3 | - | 0.0325 |
| ST6GALNAC6 | - | 0.0305 |
| STAB2 | - | 0.0467 |
| STMN3 | + | 0.0348 |
| STX10 | + | 0.0007 |
| STX12 | + | 0.0193 |
| STX1A | + | 0.0235 |
| STX7 | + | 0.0059 |
| TESC | + | 0.0259 |
| TFRC | - | 0.0274 |
| THBS2 | - | 0.0325 |
| THYN1 | + | 0.0325 |
| TMEM52B | + | 0.0421 |
| TMEM8B | + | 0.0325 |
| TNFRSF18 | + | 0.0274 |
| TP63 | + | 0.0165 |
| TPO | - | 0.0391 |
| TPST1 | - | 0.0275 |
| TRAPPC5 | - | 0.0176 |
| TYMP | + | 0.0077 |
| UBTD2 | + | 0.0077 |
| VTA1 | - | 0.0339 |
| XG | + | 0.0409 |
| YWHAG | + | 0.0274 |

The change in PD represents (+) increased and (-) decreased in the corresponding endotype vs controls, and endotype 1 vs endotype 2 respectively. Note**:** there are 198 proteins tagged by 200 SOMAmers for the contrast Endotype 2 vs HC and 153 proteins tagged by 155 SOMAmers for the contrast Endotype 1 vs 2. Duplicated SOMAmers are removed from the table for clarity (the one with the lowest *P*-value is kept). No discrepancies in directionality were present for duplicated SOMAmers. Proteins separated by comma in the same row represent the fact that due to lack of specificity the same SOMAmer is tagging more than one protein.

**Supplementary Table 5 Proteins with a significant cis-pQTL and their index SNPs**

| Gene symbol | RSID | CHR | POS | REF | ALT | ß | std.err | *P*-value |
|---|---|---|---|---|---|---|---|---|
| A4GALT | rs9611914 | 22 | 43206587 | A | T | −0.24 | 0.04 | $5.92 \times 10^{-10}$ |
| ABO | rs8176719 | 9 | 136132908 | T | TC | 1.18 | 0.03 | $2.93 \times 10^{-190}$ |
| ACBD7 | rs72774381 | 10 | 15127366 | A | G | −0.43 | 0.06 | $3.56 \times 10^{-12}$ |
| ACE | rs4335 | 17 | 61565025 | G | A | −0.25 | 0.04 | $2.87 \times 10^{-10}$ |
| ACP1 | rs7595075 | 2 | 264019 | C | A | 1.15 | 0.03 | $8.70 \times 10^{-179}$ |
| ACP5 | rs78934187 | 19 | 11695690 | A | G | 0.60 | 0.04 | $5.81 \times 10^{-43}$ |
| ACP6 | rs2153463 | 1 | 147124310 | T | G | 1.08 | 0.03 | $4.95 \times 10^{-185}$ |
| ACP7 | rs1654245 | 19 | 39583160 | G | A | −0.59 | 0.06 | $8.36 \times 10^{-24}$ |
| ACYP2 | rs3930909 | 2 | 54204922 | C | T | −0.33 | 0.06 | $9.55 \times 10^{-9}$ |
| ADA2 | rs2231495 | 22 | 17669306 | T | C | −1.00 | 0.03 | $8.47 \times 10^{-129}$ |
| ADAM12 | rs10794080 | 10 | 128053754 | T | C | 0.37 | 0.04 | $3.61 \times 10^{-22}$ |
| ADAM9 | rs62504410 | 8 | 38822313 | T | C | 0.14 | 0.02 | $6.63 \times 10^{-9}$ |
| ADAMTS1 | rs56251528 | 21 | 28210854 | C | T | −0.61 | 0.09 | $7.28 \times 10^{-11}$ |
| ADAMTS13 | rs28647808 | 9 | 136305530 | C | G | −0.46 | 0.07 | $1.19 \times 10^{-10}$ |
| ADAMTS4 | rs4233367 | 1 | 161163037 | T | C | 0.66 | 0.04 | $1.76 \times 10^{-60}$ |
| ADAMTS5 | rs2830580 | 21 | 28291149 | C | T | −1.00 | 0.04 | $1.50 \times 10^{-104}$ |
| ADAMTSL1 | rs1832638 | 9 | 18494190 | T | C | 0.29 | 0.03 | $3.80 \times 10^{-18}$ |
| ADGRE2 | rs7256892 | 19 | 14878003 | C | T | −0.55 | 0.04 | $4.17 \times 10^{-33}$ |
| ADGRF5 | rs475991 | 6 | 46822349 | A | G | 0.24 | 0.04 | $1.20 \times 10^{-8}$ |
| ADH1B | rs1229984 | 4 | 100239319 | T | C | 0.37 | 0.06 | $1.25 \times 10^{-9}$ |
| ADM | rs10743132 | 11 | 10266998 | A | G | 0.20 | 0.03 | $2.10 \times 10^{-11}$ |
| AGRP | rs4360931 | 16 | 67471926 | C | G | 0.65 | 0.07 | $4.44 \times 10^{-19}$ |
| AGT | rs12028675 | 1 | 230832275 | A | G | −0.36 | 0.04 | $1.97 \times 10^{-23}$ |
| AHSG | rs4917 | 3 | 186337713 | T | C | −1.27 | 0.03 | $2.87 \times 10^{-216}$ |
| AK1 | rs73602391 | 9 | 130661356 | G | A | −0.49 | 0.08 | $9.71 \times 10^{-10}$ |
| AKR1A1 | rs7520156 | 1 | 45912262 | A | T | −1.57 | 0.07 | $4.15 \times 10^{-83}$ |
| AKR1B1 | rs113764574 | 7 | 134207748 | G | A | −0.20 | 0.04 | $2.54 \times 10^{-8}$ |
| AKR1C1 | rs11594322 | 10 | 4956083 | A | C | −0.44 | 0.05 | $1.92 \times 10^{-16}$ |
| AKR7A2 | rs11800204 | 1 | 19576096 | T | C | −0.42 | 0.05 | $1.67 \times 10^{-18}$ |
| AMIGO2 | rs58366817 | 12 | 47934532 | T | C | −0.27 | 0.04 | $3.32 \times 10^{-10}$ |
| AMY2B | rs17014910 | 1 | 104104990 | C | G | 0.28 | 0.04 | $2.12 \times 10^{-11}$ |
| ANG | rs12433905 | 14 | 21158303 | G | A | 0.62 | 0.06 | $1.17 \times 10^{-20}$ |

| | | | | | | | | |
|---|---|---|---|---|---|---|---|---|
| ANGPTL1 | rs61823772 | 1 | 178925881 | C | G | −0.33 | 0.04 | $2.22 \times 10^{-16}$ |
| ANGPTL4 | rs34099346 | 19 | 8503345 | G | C | 0.33 | 0.06 | $3.02 \times 10^{-9}$ |
| ANGPTL7 | rs28729193 | 1 | 11180151 | T | C | −0.20 | 0.04 | $1.59 \times 10^{-8}$ |
| ANXA2 | rs12900365 | 15 | 60644463 | T | A | 0.20 | 0.03 | $2.62 \times 10^{-9}$ |
| ANXA5 | rs71606236 | 4 | 122580725 | T | C | −0.33 | 0.04 | $3.16 \times 10^{-14}$ |
| AOC1 | rs62492368 | 7 | 150537635 | G | A | 0.92 | 0.04 | $3.68 \times 10^{-94}$ |
| APCDD1 | rs3748415 | 18 | 10471732 | G | A | −0.43 | 0.07 | $1.27 \times 10^{-10}$ |
| APOC2 | rs429358 | 19 | 45411941 | T | C | 0.89 | 0.06 | $3.56 \times 10^{-39}$ |
| APOE | rs429358 | 19 | 45411941 | T | C | 0.82 | 0.05 | $2.49 \times 10^{-45}$ |
| APOH | rs8178824 | 17 | 64224775 | C | T | −1.63 | 0.08 | $1.13 \times 10^{-78}$ |
| APOL1 | rs10854688 | 22 | 36653854 | C | T | 0.20 | 0.03 | $4.54 \times 10^{-9}$ |
| ARHGAP1 | rs79920646 | 11 | 46934260 | C | G | −0.32 | 0.05 | $6.83 \times 10^{-11}$ |
| ARHGAP25 | rs10048745 | 2 | 68962137 | G | A | −0.40 | 0.04 | $3.25 \times 10^{-23}$ |
| ARSA | rs6151419 | 22 | 51064915 | G | A | −0.51 | 0.05 | $2.01 \times 10^{-22}$ |
| ARSB | rs13159135 | 5 | 78196689 | G | C | −1.02 | 0.03 | $1.30 \times 10^{-127}$ |
| ARSK | rs9314144 | 5 | 94797432 | T | C | −0.60 | 0.08 | $2.55 \times 10^{-12}$ |
| ART3 | rs12509988 | 4 | 76989108 | A | G | −0.65 | 0.05 | $9.32 \times 10^{-37}$ |
| ASAH2 | rs202183815 | 10 | 51994615 | C | T | −0.79 | 0.04 | $6.27 \times 10^{-62}$ |
| ASIP | rs62209647 | 20 | 32505658 | G | C | 0.62 | 0.08 | $2.87 \times 10^{-13}$ |
| ASL | rs77627477 | 7 | 66166078 | CTG | C | −0.34 | 0.04 | $6.31 \times 10^{-17}$ |
| ASPN | rs10120980 | 9 | 95020914 | A | G | −0.26 | 0.04 | $3.79 \times 10^{-11}$ |
| ATF6B | rs805262 | 6 | 31628733 | C | T | 0.17 | 0.03 | $4.17 \times 10^{-8}$ |
| ATP1B2 | rs1642764 | 17 | 7557834 | C | T | −0.20 | 0.03 | $7.29 \times 10^{-10}$ |
| B3GAT3 | rs10897288 | 11 | 62386472 | A | T | 0.37 | 0.03 | $3.67 \times 10^{-26}$ |
| B3GLCT | rs11147458 | 13 | 31823239 | A | G | −0.41 | 0.05 | $4.93 \times 10^{-16}$ |
| B3GNT8 | rs284663 | 19 | 41932612 | C | T | 0.68 | 0.03 | $2.70 \times 10^{-70}$ |
| B4GALNT1 | rs34919374 | 12 | 58053464 | G | A | 0.35 | 0.04 | $2.46 \times 10^{-14}$ |
| B4GALT1 | rs4879664 | 9 | 33108246 | T | G | −0.19 | 0.03 | $1.77 \times 10^{-9}$ |
| B4GALT2 | rs3216995 | 1 | 44443874 | T | TGAC | −0.56 | 0.06 | $3.06 \times 10^{-22}$ |
| B4GALT6 | rs16962266 | 18 | 29232293 | T | C | −0.56 | 0.04 | $7.35 \times 10^{-33}$ |
| BCAM | rs1135062 | 19 | 45322744 | A | G | 0.26 | 0.03 | $1.02 \times 10^{-13}$ |
| BCAN | rs2365715 | 1 | 156615114 | A | G | −0.51 | 0.04 | $2.46 \times 10^{-38}$ |
| BCHE | rs3908218 | 3 | 165462643 | A | G | −0.27 | 0.04 | $9.99 \times 10^{-12}$ |

| | | | | | | | | |
|---|---|---|---|---|---|---|---|---|
| BOC | rs3856719 | 3 | 112996541 | T | C | 0.40 | 0.05 | $4.30 \times 10^{-16}$ |
| BPI | rs1341024 | 20 | 36932676 | G | C | 0.20 | 0.03 | $3.92 \times 10^{-15}$ |
| BPIFB1 | rs11699009 | 20 | 31688241 | T | C | −0.43 | 0.05 | $4.38 \times 10^{-20}$ |
| BRPF1 | rs2269112 | 3 | 9788168 | C | T | 0.61 | 0.05 | $1.42 \times 10^{-27}$ |
| BST1 | rs4263397 | 4 | 15739390 | T | G | −1.12 | 0.04 | $2.91 \times 10^{-141}$ |
| BTD | rs6784108 | 3 | 15739777 | C | A | −0.38 | 0.04 | $9.03 \times 10^{-20}$ |
| BTN2A1 | rs2237235 | 6 | 26391395 | A | G | 0.50 | 0.03 | $7.10 \times 10^{-42}$ |
| BTN3A1 | rs9379851 | 6 | 26354780 | A | C | −1.14 | 0.05 | $3.43 \times 10^{-84}$ |
| BTNL9 | rs34492237 | 5 | 180295971 | C | A | 0.27 | 0.04 | $1.04 \times 10^{-9}$ |
| C1QC | rs12058824 | 1 | 22963050 | G | A | 0.67 | 0.08 | $3.52 \times 10^{-16}$ |
| C1QL1 | rs4793169 | 17 | 43152877 | T | C | −0.30 | 0.03 | $3.13 \times 10^{-17}$ |
| C1QL4 | rs7972869 | 12 | 49731167 | A | G | 0.36 | 0.06 | $3.88 \times 10^{-10}$ |
| C1QTNF3 | rs7712366 | 5 | 34038537 | A | G | 0.56 | 0.04 | $4.57 \times 10^{-34}$ |
| C1QTNF5 | rs2248863 | 11 | 119207341 | G | A | 0.52 | 0.07 | $3.17 \times 10^{-14}$ |
| C1S | rs7962629 | 12 | 7166770 | A | G | −0.37 | 0.06 | $6.63 \times 10^{-11}$ |
| C2 | rs116198852 | 6 | 31393014 | C | T | −0.52 | 0.07 | $5.43 \times 10^{-13}$ |
| C4A | rs2280774 | 6 | 31928691 | G | A | −0.95 | 0.04 | $9.57 \times 10^{-105}$ |
| C4B | rs2280774 | 6 | 31928691 | G | A | −0.95 | 0.04 | $9.57 \times 10^{-105}$ |
| C7 | rs147512353 | 5 | 40971249 | T | C | −0.32 | 0.05 | $2.40 \times 10^{-10}$ |
| CA4 | rs923038 | 17 | 58220132 | T | G | −0.63 | 0.08 | $2.09 \times 10^{-15}$ |
| CA6 | rs3765963 | 1 | 9034598 | A | G | 0.32 | 0.04 | $1.55 \times 10^{-13}$ |
| CA9 | rs2071676 | 9 | 35674053 | G | A | 0.28 | 0.04 | $2.66 \times 10^{-10}$ |
| CABLES2 | rs2427312 | 20 | 60970591 | C | T | −0.32 | 0.04 | $1.70 \times 10^{-15}$ |
| CALCOCO2 | rs550510 | 17 | 46926615 | G | A | −0.30 | 0.04 | $3.91 \times 10^{-14}$ |
| CAPG | rs4446071 | 2 | 85543064 | T | C | −0.42 | 0.05 | $1.27 \times 10^{-18}$ |
| CAPSL | rs6865274 | 5 | 35923829 | A | G | −0.39 | 0.07 | $1.28 \times 10^{-8}$ |
| CAT | rs3781709 | 11 | 34504711 | T | C | −0.32 | 0.05 | $5.31 \times 10^{-10}$ |
| CBR1 | rs1005696 | 21 | 37443480 | T | G | −0.30 | 0.04 | $8.01 \times 10^{-12}$ |
| CBR3 | rs1056892 | 21 | 37518706 | G | A | −0.60 | 0.03 | $4.04 \times 10^{-60}$ |
| CCDC50 | rs56134151 | 3 | 191045679 | C | T | −0.20 | 0.04 | $9.99 \times 10^{-9}$ |
| CCL14 | rs7222922 | 17 | 34335694 | C | T | −0.92 | 0.05 | $1.58 \times 10^{-63}$ |
| CCL15 | rs41508645 | 17 | 34329044 | T | G | 1.13 | 0.06 | $7.86 \times 10^{-58}$ |

| | | | | | | | | |
|---|---|---|---|---|---|---|---|---|
| CCL16 | rs112689088 | 17 | 34307457 | T | C | −1.30 | 0.05 | $5.62 \times 10^{-95}$ |
| CCL17 | rs223896 | 16 | 57443146 | G | A | −0.37 | 0.04 | $3.37 \times 10^{-16}$ |
| CCL18 | rs2015086 | 17 | 34391617 | A | G | 0.90 | 0.06 | $1.63 \times 10^{-47}$ |
| CCL23 | rs7208990 | 17 | 34329475 | C | G | −0.79 | 0.08 | $4.05 \times 10^{-23}$ |
| CCL25 | rs62124694 | 19 | 8126830 | A | C | 0.27 | 0.05 | $2.99 \times 10^{-8}$ |
| CCL3L1 | rs56683451 | 17 | 34374696 | T | C | 0.45 | 0.07 | $3.29 \times 10^{-11}$ |
| CCL7 | rs3138036 | 17 | 32647544 | A | G | −0.50 | 0.06 | $3.47 \times 10^{-14}$ |
| CCL8 | rs3138036 | 17 | 32647544 | A | G | −1.25 | 0.05 | $2.78 \times 10^{-106}$ |
| CCN3 | rs2469990 | 8 | 120344567 | A | G | −0.39 | 0.05 | $2.74 \times 10^{-14}$ |
| CCN4 | rs2739063 | 8 | 133948522 | A | G | −1.03 | 0.03 | $2.63 \times 10^{-132}$ |
| CCN5 | rs11086936 | 20 | 43331807 | C | G | 0.44 | 0.03 | $6.73 \times 10^{-33}$ |
| CD109 | rs10223501 | 6 | 74419424 | C | T | 0.41 | 0.04 | $5.84 \times 10^{-25}$ |
| CD14 | rs12517200 | 5 | 140019921 | A | G | 0.34 | 0.03 | $2.33 \times 10^{-21}$ |
| CD177 | rs78530667 | 19 | 43900697 | T | C | −0.74 | 0.07 | $2.59 \times 10^{-23}$ |
| CD209 | rs4804774 | 19 | 7772173 | G | C | −0.57 | 0.05 | $1.94 \times 10^{-29}$ |
| CD274 | rs7048841 | 9 | 5460801 | T | C | −0.53 | 0.03 | $2.48 \times 10^{-66}$ |
| CD300A | rs2272111 | 17 | 72469966 | G | A | −0.58 | 0.04 | $3.80 \times 10^{-49}$ |
| CD300C | rs2670826 | 17 | 72462466 | C | T | −0.28 | 0.04 | $4.05 \times 10^{-14}$ |
| CD300E | rs581157 | 17 | 72613589 | T | G | −0.67 | 0.04 | $5.52 \times 10^{-56}$ |
| CD33 | rs3865444 | 19 | 51727962 | C | A | −1.03 | 0.03 | $2.15 \times 10^{-154}$ |
| CD38 | rs6836946 | 4 | 15813607 | T | G | 0.31 | 0.05 | $5.73 \times 10^{-10}$ |
| CD48 | rs12124234 | 1 | 160675269 | G | C | 0.28 | 0.05 | $5.15 \times 10^{-9}$ |
| CD55 | rs116059019 | 1 | 207572442 | G | A | 0.56 | 0.05 | $6.98 \times 10^{-26}$ |
| CD59 | rs3181269 | 11 | 33755956 | C | T | −0.23 | 0.03 | $4.57 \times 10^{-15}$ |
| CD72 | rs7026779 | 9 | 35581458 | A | G | 0.71 | 0.04 | $1.14 \times 10^{-48}$ |
| CD83 | rs11756936 | 6 | 14110109 | G | C | 0.27 | 0.03 | $2.53 \times 10^{-15}$ |
| CD84 | rs796499427 | 1 | 160574660 | AAC | A | −0.50 | 0.06 | $1.34 \times 10^{-15}$ |
| CD8A | rs3020726 | 2 | 87016506 | A | G | 0.67 | 0.06 | $2.42 \times 10^{-26}$ |
| CD93 | rs11697283 | 20 | 23108635 | A | G | 0.25 | 0.04 | $1.50 \times 10^{-8}$ |
| CDCP1 | rs2276862 | 3 | 45187785 | C | G | −0.43 | 0.05 | $1.10 \times 10^{-16}$ |
| CDH11 | rs1124695 | 16 | 65037770 | C | G | −1.26 | 0.04 | $5.97 \times 10^{-154}$ |
| CDNF | rs72772427 | 10 | 14911640 | C | T | 0.65 | 0.04 | $4.93 \times 10^{-57}$ |
| CDON | rs35088114 | 11 | 125842801 | G | GGCA | −0.53 | 0.06 | $1.63 \times 10^{-18}$ |

| | | | | | | | | |
|---|---|---|---|---|---|---|---|---|
| CEACAM19 | rs429358 | 19 | 45411941 | T | C | 0.46 | 0.05 | $4.05 \times 10^{-17}$ |
| CEL | rs668809 | 9 | 135936678 | A | G | 1.28 | 0.10 | $8.17 \times 10^{-37}$ |
| CER1 | rs2131882 | 9 | 14716849 | T | A | 0.89 | 0.06 | $6.77 \times 10^{-44}$ |
| CFB | rs9296004 | 6 | 31933977 | A | C | −0.47 | 0.07 | $2.41 \times 10^{-10}$ |
| CFD | rs71335276 | 19 | 859368 | G | T | 0.93 | 0.05 | $5.16 \times 10^{-74}$ |
| CFH | rs35617250 | 1 | 196679682 | C | T | 0.40 | 0.05 | $1.35 \times 10^{-13}$ |
| CFHR1 | rs71631867 | 1 | 196814850 | T | A | −0.86 | 0.04 | $1.01 \times 10^{-81}$ |
| CFHR4 | rs200880049 | 1 | 196790437 | T | A | −0.17 | 0.03 | $2.38 \times 10^{-11}$ |
| CFHR5 | rs1694459 | 1 | 196815786 | C | T | 0.33 | 0.04 | $1.04 \times 10^{-13}$ |
| CFI | rs7439493 | 4 | 110656730 | G | A | 0.21 | 0.04 | $2.68 \times 10^{-8}$ |
| CGB3 | rs78248023 | 19 | 49515171 | C | A | 0.97 | 0.06 | $2.72 \times 10^{-50}$ |
| CGB7 | rs78248023 | 19 | 49515171 | C | A | 0.97 | 0.06 | $2.72 \times 10^{-50}$ |
| CHGA | rs729940 | 14 | 93399101 | C | T | −0.27 | 0.04 | $4.92 \times 10^{-12}$ |
| CHI3L1 | rs946259 | 1 | 203152177 | C | T | −1.07 | 0.03 | $2.09 \times 10^{-183}$ |
| CHI3L2 | rs11556868 | 1 | 111778325 | C | T | 1.42 | 0.05 | $8.36 \times 10^{-127}$ |
| CHIT1 | rs2486063 | 1 | 203173094 | A | G | −1.04 | 0.05 | $7.77 \times 10^{-82}$ |
| CHRD | rs73053799 | 3 | 184111595 | C | T | 0.36 | 0.05 | $8.46 \times 10^{-13}$ |
| CHRDL2 | rs7104802 | 11 | 74386800 | G | T | 0.35 | 0.03 | $7.22 \times 10^{-27}$ |
| CHST12 | rs3735099 | 7 | 2472429 | C | A | 0.73 | 0.06 | $5.51 \times 10^{-32}$ |
| CHST9 | rs12968277 | 18 | 24637729 | G | C | 0.91 | 0.03 | $4.71 \times 10^{-119}$ |
| CLEC11A | rs11666008 | 19 | 51226590 | C | G | −0.35 | 0.05 | $2.93 \times 10^{-11}$ |
| CLEC12A | rs607567 | 12 | 10131684 | T | G | −1.12 | 0.03 | $3.05 \times 10^{-179}$ |
| CLEC2L | rs12672312 | 7 | 139230464 | C | T | 0.16 | 0.02 | $5.49 \times 10^{-11}$ |
| CLEC3B | rs4683019 | 3 | 45040682 | T | C | −0.33 | 0.03 | $4.61 \times 10^{-23}$ |
| CLEC4C | rs10845821 | 12 | 7900184 | C | T | 0.56 | 0.04 | $4.37 \times 10^{-35}$ |
| CLEC4G | rs115293707 | 19 | 7794609 | G | C | 1.39 | 0.07 | $1.07 \times 10^{-73}$ |
| CLEC7A | rs16910526 | 12 | 10271087 | A | C | −1.48 | 0.07 | $2.48 \times 10^{-82}$ |
| CLIC5 | rs76820035 | 6 | 45919515 | G | A | 0.78 | 0.08 | $8.69 \times 10^{-21}$ |
| CLN5 | rs2794618 | 13 | 77584224 | G | A | −0.71 | 0.07 | $1.32 \times 10^{-21}$ |
| CLPS | rs12193764 | 6 | 35742556 | C | G | 0.48 | 0.05 | $3.24 \times 10^{-21}$ |
| CNDP1 | rs4329999 | 18 | 72228269 | G | A | −0.48 | 0.04 | $1.20 \times 10^{-28}$ |
| CNTN2 | rs41264869 | 1 | 205030862 | C | T | −0.68 | 0.05 | $3.66 \times 10^{-32}$ |
| CNTN4 | rs163352 | 3 | 3098041 | G | C | −0.18 | 0.02 | $8.68 \times 10^{-13}$ |

| | | | | | | | | |
|---|---|---|---|---|---|---|---|---|
| CNTN5 | rs72998295 | 11 | 99093036 | T | C | −0.25 | 0.04 | $4.89 \times 10^{-9}$ |
| COCH | rs66816140 | 14 | 31239287 | AC | A | 0.67 | 0.04 | $2.71 \times 10^{-47}$ |
| COL11A2 | rs17847933 | 6 | 33165780 | G | C | 0.74 | 0.08 | $3.51 \times 10^{-20}$ |
| COL15A1 | rs7025094 | 9 | 101667626 | G | A | 0.31 | 0.04 | $1.26 \times 10^{-12}$ |
| COL18A1 | rs12483377 | 21 | 46931109 | G | A | −0.42 | 0.06 | $1.01 \times 10^{-10}$ |
| COL23A1 | rs4976753 | 5 | 177681651 | C | T | −0.81 | 0.09 | $1.36 \times 10^{-17}$ |
| COLEC11 | rs11123637 | 2 | 3654190 | T | C | −1.06 | 0.03 | $3.63 \times 10^{-140}$ |
| CPA4 | rs34587586 | 7 | 129938598 | G | T | −1.14 | 0.03 | $2.48 \times 10^{-176}$ |
| CPB1 | rs13079446 | 3 | 148509614 | T | A | −0.84 | 0.06 | $6.26 \times 10^{-38}$ |
| CPB2 | rs9567615 | 13 | 46651080 | A | C | 0.30 | 0.04 | $2.91 \times 10^{-12}$ |
| CPE | rs72703629 | 4 | 166346061 | T | C | 0.27 | 0.03 | $2.73 \times 10^{-15}$ |
| CPM | rs12372220 | 12 | 69371471 | C | T | −0.77 | 0.05 | $1.21 \times 10^{-55}$ |
| CPN2 | rs6809081 | 3 | 194060475 | T | C | −0.34 | 0.04 | $8.78 \times 10^{-15}$ |
| CPXM1 | rs11697820 | 20 | 2781441 | C | T | 0.91 | 0.04 | $4.39 \times 10^{-101}$ |
| CPZ | rs13121547 | 4 | 8601592 | G | T | −0.51 | 0.04 | $1.27 \times 10^{-29}$ |
| CREB3L4 | rs140257179 | 1 | 153963995 | A | AAG | 0.38 | 0.04 | $2.57 \times 10^{-19}$ |
| CRELD1 | rs6762702 | 3 | 10055789 | G | A | −0.62 | 0.04 | $6.18 \times 10^{-58}$ |
| CRHBP | rs6871700 | 5 | 76209399 | A | G | −0.28 | 0.04 | $3.10 \times 10^{-10}$ |
| CRIM1 | rs10189344 | 2 | 36778887 | A | C | −0.22 | 0.03 | $9.03 \times 10^{-12}$ |
| CRISP2 | rs10948527 | 6 | 49746832 | C | T | −0.44 | 0.05 | $1.07 \times 10^{-18}$ |
| CRISPLD2 | rs1466716 | 16 | 84870034 | T | C | 0.51 | 0.06 | $8.58 \times 10^{-16}$ |
| CRLF1 | rs3761031 | 19 | 18721451 | G | A | 0.39 | 0.04 | $2.07 \times 10^{-22}$ |
| CROT | rs7776867 | 7 | 87014355 | C | T | −0.55 | 0.08 | $1.09 \times 10^{-11}$ |
| CRTAC1 | rs10883027 | 10 | 99798234 | G | C | 0.16 | 0.02 | $3.21 \times 10^{-14}$ |
| CRYGD | rs55770589 | 2 | 208985427 | T | C | 0.32 | 0.03 | $2.15 \times 10^{-24}$ |
| CRYZ | rs3819946 | 1 | 75175886 | T | C | 1.02 | 0.04 | $2.91 \times 10^{-101}$ |
| CSGALNACT2 | rs2435381 | 10 | 43678796 | C | T | −0.98 | 0.04 | $6.93 \times 10^{-118}$ |
| CST1 | rs113896553 | 20 | 23731589 | G | C | 1.10 | 0.03 | $1.24 \times 10^{-143}$ |
| CST2 | rs113896553 | 20 | 23731589 | G | C | 1.09 | 0.03 | $1.92 \times 10^{-148}$ |
| CST3 | rs2983644 | 20 | 23592966 | C | A | 0.38 | 0.03 | $3.93 \times 10^{-30}$ |
| CST4 | rs7264810 | 20 | 23673625 | C | A | −0.54 | 0.07 | $1.19 \times 10^{-13}$ |
| CST5 | rs4239743 | 20 | 23859017 | A | C | 0.69 | 0.04 | $3.04 \times 10^{-59}$ |

| | | | | | | | | |
|---|---|---|---|---|---|---|---|---|
| CST6 | rs1131544 | 11 | 65779590 | C | T | 1.18 | 0.04 | $1.02 \times 10^{-126}$ |
| CST7 | rs148851012 | 20 | 24973964 | ATTTGATAAAGTTTATTAAGT | A | 0.85 | 0.07 | $2.74 \times 10^{-32}$ |
| CTHRC1 | rs10093100 | 8 | 104368191 | T | C | 0.41 | 0.04 | $3.27 \times 10^{-19}$ |
| CTNNA2 | rs72917277 | 2 | 79549662 | A | G | −0.41 | 0.06 | $1.26 \times 10^{-10}$ |
| CTRB1 | rs113366803 | 16 | 75299391 | A | G | 0.45 | 0.08 | $3.42 \times 10^{-8}$ |
| CTRB2 | rs889515 | 16 | 75251299 | T | C | −1.00 | 0.05 | $1.35 \times 10^{-65}$ |
| CTSB | rs28577034 | 8 | 11708715 | A | T | −0.68 | 0.04 | $1.76 \times 10^{-50}$ |
| CTSC | rs11600158 | 11 | 88070914 | A | G | 0.89 | 0.07 | $3.39 \times 10^{-37}$ |
| CTSF | rs1127894 | 11 | 66335548 | A | G | 0.26 | 0.04 | $7.79 \times 10^{-12}$ |
| CTSH | rs2289702 | 15 | 79237293 | C | T | −1.49 | 0.06 | $9.07 \times 10^{-90}$ |
| CTSK | rs55890313 | 1 | 150640078 | T | A | −0.47 | 0.07 | $5.00 \times 10^{-12}$ |
| CTSO | rs138798877 | 4 | 156936516 | ATATT | A | −0.46 | 0.08 | $3.91 \times 10^{-8}$ |
| CTSS | rs41271951 | 1 | 150737220 | A | G | −1.26 | 0.09 | $1.04 \times 10^{-41}$ |
| CTSZ | rs163787 | 20 | 57574463 | A | G | 0.40 | 0.05 | $3.58 \times 10^{-14}$ |
| CXCL1 | rs7438101 | 4 | 74728327 | T | C | −0.33 | 0.05 | $9.90 \times 10^{-10}$ |
| CXCL6 | rs62312418 | 4 | 74701512 | G | A | 0.35 | 0.04 | $5.70 \times 10^{-16}$ |
| CYB5D2 | rs35103069 | 17 | 4108063 | G | C | −0.21 | 0.03 | $2.44 \times 10^{-9}$ |
| CYTL1 | rs6446312 | 4 | 5019932 | C | G | −0.69 | 0.05 | $9.77 \times 10^{-39}$ |
| DEFB1 | rs2978857 | 8 | 6741305 | T | G | −1.18 | 0.03 | $3.47 \times 10^{-157}$ |
| DKK2 | rs17037069 | 4 | 107838076 | A | G | −0.62 | 0.07 | $4.69 \times 10^{-16}$ |
| DLK1 | rs1058009 | 14 | 101200860 | G | A | −0.62 | 0.09 | $8.54 \times 10^{-12}$ |
| DLL1 | rs9459968 | 6 | 170497591 | C | T | −0.32 | 0.04 | $5.38 \times 10^{-18}$ |
| DNAJC10 | rs746705411 | 2 | 183612210 | CTT | C | 0.18 | 0.03 | $2.00 \times 10^{-9}$ |
| DNAJC30 | rs10479665 | 7 | 73108924 | A | G | −0.88 | 0.10 | $4.14 \times 10^{-18}$ |
| DNER | rs67033065 | 2 | 230532197 | ATCCT | A | −0.19 | 0.02 | $3.40 \times 10^{-17}$ |
| DPP7 | rs6560658 | 9 | 140004229 | C | G | 0.82 | 0.05 | $4.12 \times 10^{-57}$ |
| DPT | rs12048750 | 1 | 168922582 | G | A | 0.44 | 0.04 | $6.08 \times 10^{-22}$ |
| DRAXIN | rs1555079 | 1 | 11760782 | C | T | −0.71 | 0.04 | $6.37 \times 10^{-72}$ |
| DSC2 | rs1789064 | 18 | 28674402 | A | T | −0.49 | 0.04 | $8.70 \times 10^{-35}$ |
| DSC3 | rs1370450 | 18 | 28741821 | G | A | −0.23 | 0.04 | $7.35 \times 10^{-9}$ |

| | | | | | | | | |
|---|---|---|---|---|---|---|---|---|
| DSCAM | rs401936 | 21 | 41449867 | C | T | 0.27 | 0.02 | $1.71 \times 10^{-26}$ |
| DSG2 | rs9304098 | 18 | 29083630 | G | T | −0.45 | 0.04 | $6.00 \times 10^{-24}$ |
| EBI3 | rs4740 | 19 | 4236996 | G | A | −1.18 | 0.03 | $1.95 \times 10^{-179}$ |
| ECH1 | rs2229259 | 19 | 39307103 | C | T | 0.58 | 0.06 | $2.98 \times 10^{-18}$ |
| ECM1 | rs9326002 | 1 | 150495106 | A | G | −0.42 | 0.03 | $2.03 \times 10^{-51}$ |
| EDAR | rs260689 | 2 | 109579110 | G | A | 0.36 | 0.06 | $2.60 \times 10^{-8}$ |
| EDIL3 | rs13160212 | 5 | 83219474 | T | C | 0.42 | 0.05 | $2.60 \times 10^{-19}$ |
| EFEMP1 | rs1430192 | 2 | 56117040 | T | C | 0.39 | 0.03 | $1.14 \times 10^{-32}$ |
| EFNA5 | rs7709385 | 5 | 107047072 | G | T | −0.18 | 0.03 | $1.23 \times 10^{-8}$ |
| EGF | rs4698801 | 4 | 110882476 | C | A | 0.38 | 0.04 | $6.48 \times 10^{-24}$ |
| EGFLAM | rs12516477 | 5 | 38245951 | T | C | 0.84 | 0.08 | $1.81 \times 10^{-26}$ |
| EGFR | rs151057105 | 7 | 54944920 | C | T | −0.66 | 0.04 | $4.80 \times 10^{-43}$ |
| EHF | rs286924 | 11 | 34642728 | A | T | −0.59 | 0.09 | $1.81 \times 10^{-10}$ |
| EMILIN3 | rs6102384 | 20 | 39997745 | T | C | 0.41 | 0.07 | $1.05 \times 10^{-8}$ |
| ENDOU | rs7976708 | 12 | 48116255 | A | G | −0.58 | 0.05 | $1.03 \times 10^{-26}$ |
| ENPP5 | rs62400861 | 6 | 46100229 | C | G | −1.52 | 0.04 | $3.85 \times 10^{-173}$ |
| ENPP7 | rs36069406 | 17 | 77696496 | GC | G | 0.80 | 0.04 | $4.56 \times 10^{-78}$ |
| ENTPD3 | rs2276868 | 3 | 40498845 | C | T | 0.31 | 0.04 | $2.08 \times 10^{-13}$ |
| ENTPD6 | rs3859660 | 20 | 25915180 | A | T | 0.68 | 0.07 | $1.47 \times 10^{-20}$ |
| EPDR1 | rs2044831 | 7 | 37988589 | C | T | −0.35 | 0.05 | $7.07 \times 10^{-13}$ |
| EPHA1 | rs4725617 | 7 | 143097100 | A | G | 1.44 | 0.08 | $4.17 \times 10^{-66}$ |
| EPHA10 | rs12049522 | 1 | 38245517 | T | G | −0.14 | 0.02 | $1.40 \times 10^{-8}$ |
| EPHA2 | rs28452540 | 1 | 16423834 | A | G | −0.23 | 0.04 | $2.11 \times 10^{-9}$ |
| EPHA5 | rs10434242 | 4 | 66518523 | A | G | 0.14 | 0.02 | $1.65 \times 10^{-9}$ |
| EPO | rs114129687 | 7 | 100290926 | T | G | −0.39 | 0.04 | $2.79 \times 10^{-20}$ |
| ERAP1 | rs26653 | 5 | 96139250 | C | G | −0.95 | 0.03 | $7.78 \times 10^{-117}$ |
| ERAP2 | rs2927608 | 5 | 96252432 | G | A | 1.23 | 0.03 | $1.53 \times 10^{-225}$ |
| ERBB4 | rs12987104 | 2 | 213302799 | A | G | −0.27 | 0.04 | $6.13 \times 10^{-11}$ |
| ERLEC1 | rs17039525 | 2 | 53955513 | A | C | −0.14 | 0.02 | $1.04 \times 10^{-9}$ |
| ERLIN1 | rs2862954 | 10 | 101912064 | T | C | 0.36 | 0.04 | $4.48 \times 10^{-15}$ |
| ERO1B | rs2477599 | 1 | 236413230 | T | A | −1.01 | 0.03 | $4.86 \times 10^{-145}$ |
| ESD | rs1216970 | 13 | 47374012 | C | G | −1.29 | 0.06 | $7.03 \times 10^{-76}$ |
| ESM1 | rs78414307 | 5 | 54434781 | TA | T | −0.38 | 0.05 | $5.97 \times 10^{-12}$ |

| | | | | | | | | |
|---|---|---|---|---|---|---|---|---|
| EYS | rs9342464 | 6 | 66005888 | C | T | 0.22 | 0.04 | $4.08 \times 10^{-8}$ |
| F11 | rs6824705 | 4 | 187213889 | C | T | 0.30 | 0.04 | $4.64 \times 10^{-14}$ |
| F13B | rs35258876 | 1 | 197151626 | C | CA | −0.25 | 0.04 | $6.33 \times 10^{-11}$ |
| F5 | rs72708013 | 1 | 169481731 | T | G | −0.87 | 0.09 | $1.13 \times 10^{-20}$ |
| FABP1 | rs2241883 | 2 | 88424066 | T | C | −0.24 | 0.04 | $1.05 \times 10^{-10}$ |
| FABP3 | rs879491795 | 1 | 31676195 | AG | A | 0.24 | 0.04 | $5.61 \times 10^{-9}$ |
| FABP6 | rs10063628 | 5 | 159631958 | C | G | −0.61 | 0.05 | $8.47 \times 10^{-35}$ |
| FAH | rs34239630 | 15 | 80441888 | C | T | −0.35 | 0.05 | $1.47 \times 10^{-11}$ |
| FAIM | rs641320 | 3 | 138347957 | G | A | 1.47 | 0.08 | $2.43 \times 10^{-69}$ |
| FAM177A1 | rs79698726 | 14 | 35522350 | A | C | −0.22 | 0.03 | $6.18 \times 10^{-12}$ |
| FAM20A | rs12602247 | 17 | 66685034 | C | G | 0.30 | 0.04 | $6.23 \times 10^{-11}$ |
| FAM234B | rs200946929 | 12 | 13201461 | TCTG TTAT CC | T | 0.18 | 0.03 | $1.74 \times 10^{-8}$ |
| FAM3B | rs66817580 | 21 | 42689637 | G | T | 1.42 | 0.04 | $1.50 \times 10^{-147}$ |
| FAM3D | rs3749290 | 3 | 58652292 | G | T | −0.67 | 0.06 | $4.38 \times 10^{-28}$ |
| FAS | rs6586163 | 10 | 90752018 | A | C | 0.90 | 0.03 | $6.05 \times 10^{-121}$ |
| FBP1 | rs28369691 | 9 | 97384265 | C | T | 0.68 | 0.08 | $8.56 \times 10^{-17}$ |
| FBP2 | rs10761342 | 9 | 97346079 | C | T | 0.28 | 0.05 | $2.90 \times 10^{-9}$ |
| FCER1A | rs6703348 | 1 | 159291683 | C | G | 0.61 | 0.04 | $1.44 \times 10^{-38}$ |
| FCER2 | rs12980031 | 19 | 7764436 | G | T | −1.08 | 0.03 | $2.97 \times 10^{-141}$ |
| FCGR2A | rs1801274 | 1 | 161479745 | A | G | 1.23 | 0.02 | $5.70 \times 10^{-262}$ |
| FCGR2B | rs1801274 | 1 | 161479745 | A | G | 1.05 | 0.03 | $2.26 \times 10^{-137}$ |
| FCGR3B | rs2487452 | 1 | 161530869 | G | A | 0.47 | 0.05 | $1.38 \times 10^{-21}$ |
| FCN2 | rs7037264 | 9 | 137775212 | G | A | −0.65 | 0.04 | $1.97 \times 10^{-53}$ |
| FCRL6 | rs12088352 | 1 | 159784539 | G | A | −0.37 | 0.04 | $2.95 \times 10^{-17}$ |
| FGF17 | rs77473749 | 8 | 21877382 | C | G | 1.19 | 0.08 | $3.37 \times 10^{-46}$ |
| FGF19 | rs9667380 | 11 | 69611959 | C | T | 0.33 | 0.04 | $2.83 \times 10^{-17}$ |
| FGFBP3 | rs10881987 | 10 | 93641869 | A | G | 0.40 | 0.04 | $1.61 \times 10^{-21}$ |
| FGFRL1 | rs4647930 | 4 | 1018705 | C | A | 0.35 | 0.04 | $1.44 \times 10^{-17}$ |
| FKBP14 | rs28619411 | 7 | 30096002 | A | T | 0.37 | 0.03 | $3.18 \times 10^{-39}$ |
| FKBP7 | rs2886712 | 2 | 179311529 | G | A | 0.64 | 0.03 | $5.55 \times 10^{-72}$ |
| FLRT1 | rs588177 | 11 | 64024056 | C | A | 0.18 | 0.03 | $2.92 \times 10^{-8}$ |
| FLRT2 | rs10498583 | 14 | 85742934 | G | A | −0.17 | 0.03 | $1.85 \times 10^{-8}$ |

| | | | | | | | | |
|---|---|---|---|---|---|---|---|---|
| FLRT3 | rs150296190 | 20 | 14822666 | T | TTTTG | −0.21 | 0.03 | $1.38 \times 10^{-14}$ |
| FMOD | rs28583560 | 1 | 203315331 | G | A | −0.71 | 0.09 | $8.67 \times 10^{-15}$ |
| FOLH1 | rs56287037 | 11 | 49106344 | G | A | 1.49 | 0.10 | $1.33 \times 10^{-44}$ |
| FREM2 | rs34692381 | 13 | 39331723 | C | T | −0.63 | 0.04 | $1.64 \times 10^{-55}$ |
| FRZB | rs288326 | 2 | 183703336 | G | A | 0.79 | 0.06 | $5.16 \times 10^{-40}$ |
| FSTL1 | rs66685284 | 3 | 120172964 | AAC | A | 0.30 | 0.05 | $3.58 \times 10^{-11}$ |
| FSTL3 | rs71333301 | 19 | 676904 | CGA | C | −0.24 | 0.04 | $2.87 \times 10^{-11}$ |
| FSTL5 | rs2314103 | 4 | 162334838 | T | C | −0.58 | 0.06 | $9.24 \times 10^{-20}$ |
| FUT10 | rs16880849 | 8 | 33246538 | A | G | 0.53 | 0.03 | $2.24 \times 10^{-54}$ |
| FUT5 | rs72989070 | 19 | 5895358 | C | T | 0.91 | 0.03 | $5.33 \times 10^{-115}$ |
| FUT8 | rs7140341 | 14 | 66070417 | C | G | 0.29 | 0.04 | $4.46 \times 10^{-13}$ |
| GALNT10 | rs1106324 | 5 | 153580836 | A | G | 0.24 | 0.04 | $1.10 \times 10^{-8}$ |
| GALNT16 | rs7143324 | 14 | 69755470 | A | G | −0.32 | 0.03 | $7.71 \times 10^{-19}$ |
| GCA | rs17783344 | 2 | 163208893 | T | G | −0.40 | 0.05 | $7.10 \times 10^{-18}$ |
| GDF10 | rs11593867 | 10 | 48443093 | G | A | −0.24 | 0.04 | $9.47 \times 10^{-9}$ |
| GDF15 | rs199580670 | 19 | 18475747 | TGGC GCGG GGGG CTCA AAAC GGG | T | 0.47 | 0.04 | $1.55 \times 10^{-28}$ |
| GFRA1 | rs11197613 | 10 | 118026217 | G | T | −0.38 | 0.06 | $2.37 \times 10^{-9}$ |
| GFRAL | rs6929594 | 6 | 55218980 | G | A | 0.61 | 0.04 | $1.91 \times 10^{-50}$ |
| GGH | rs34223853 | 8 | 63953538 | CA | C | 0.46 | 0.04 | $4.12 \times 10^{-28}$ |
| GKN2 | rs13008230 | 2 | 69154583 | T | G | −0.56 | 0.10 | $4.82 \times 10^{-8}$ |
| GLCE | rs3865014 | 15 | 69561518 | G | A | 1.13 | 0.04 | $6.42 \times 10^{-133}$ |
| GLIPR1 | rs2242435 | 12 | 75874913 | G | A | −0.22 | 0.04 | $3.99 \times 10^{-8}$ |
| GLO1 | rs34637217 | 6 | 38657463 | C | T | −0.23 | 0.04 | $1.25 \times 10^{-8}$ |
| GLRX2 | rs74540121 | 1 | 193177917 | T | A | 0.25 | 0.04 | $2.51 \times 10^{-12}$ |
| GNLY | rs370615710 | 2 | 85932002 | GT | G | 0.88 | 0.05 | $6.04 \times 10^{-52}$ |
| GNRH1 | rs13261573 | 8 | 25248615 | G | A | −0.31 | 0.05 | $1.49 \times 10^{-8}$ |
| GNRH2 | rs8184100 | 20 | 3026415 | C | T | −1.22 | 0.05 | $3.95 \times 10^{-99}$ |
| GOLM1 | rs148272757 | 9 | 88714593 | C | A | 0.45 | 0.05 | $5.14 \times 10^{-17}$ |
| GP6 | rs1613662 | 19 | 55536595 | G | A | 0.44 | 0.05 | $4.44 \times 10^{-19}$ |
| GPC1 | rs60515008 | 2 | 241408434 | T | C | 0.75 | 0.06 | $2.20 \times 10^{-30}$ |
| GPC5 | rs9523326 | 13 | 92052108 | G | A | −0.30 | 0.03 | $2.47 \times 10^{-24}$ |

| | | | | | | | | |
|---|---|---|---|---|---|---|---|---|
| GPC6 | rs1535692 | 13 | 95034749 | G | A | −0.45 | 0.05 | $1.26 \times 10^{-17}$ |
| GPNMB | rs858275 | 7 | 23294144 | T | C | −0.80 | 0.03 | $5.35 \times 10^{-95}$ |
| GRN | rs5848 | 17 | 42430244 | C | T | −0.33 | 0.04 | $7.79 \times 10^{-15}$ |
| GRP | rs8091691 | 18 | 56827121 | G | A | −0.32 | 0.05 | $6.10 \times 10^{-12}$ |
| GSN | rs76098787 | 9 | 124047836 | T | C | 0.92 | 0.07 | $3.36 \times 10^{-33}$ |
| GSTM1 | rs599363 | 1 | 110218486 | G | T | −0.86 | 0.07 | $2.42 \times 10^{-32}$ |
| GSTM3 | rs12137743 | 1 | 110253555 | C | T | −0.65 | 0.04 | $4.67 \times 10^{-48}$ |
| GSTO1 | rs1147611 | 10 | 106025258 | G | T | −1.06 | 0.03 | $5.37 \times 10^{-163}$ |
| GSTP1 | rs1695 | 11 | 67352689 | A | G | −0.79 | 0.03 | $1.64 \times 10^{-106}$ |
| GSTT2B | rs5751777 | 22 | 24267047 | C | T | −0.32 | 0.04 | $5.28 \times 10^{-13}$ |
| GXYLT1 | rs11181303 | 12 | 42437801 | A | T | −0.34 | 0.06 | $8.50 \times 10^{-9}$ |
| GZMB | rs8192918 | 14 | 25101878 | C | CT | −0.97 | 0.04 | $5.96 \times 10^{-85}$ |
| GZMM | rs3760874 | 19 | 544315 | A | G | 0.39 | 0.05 | $4.21 \times 10^{-14}$ |
| H6PD | rs7555568 | 1 | 9292606 | G | A | −0.60 | 0.04 | $1.35 \times 10^{-39}$ |
| HAPLN1 | rs13160212 | 5 | 83219474 | T | C | 1.07 | 0.03 | $8.74 \times 10^{-145}$ |
| HAPLN4 | rs2965187 | 19 | 19522970 | T | A | 0.24 | 0.03 | $1.12 \times 10^{-18}$ |
| HAVCR2 | rs6873507 | 5 | 156529747 | A | G | −0.74 | 0.04 | $5.99 \times 10^{-67}$ |
| HDGF | rs12145743 | 1 | 156700651 | T | G | −0.29 | 0.03 | $3.18 \times 10^{-18}$ |
| HGF | rs10248271 | 7 | 81417306 | T | G | −0.25 | 0.04 | $1.64 \times 10^{-8}$ |
| HGFAC | rs2498323 | 4 | 3451109 | G | A | −1.28 | 0.06 | $7.43 \times 10^{-82}$ |
| HIBCH | rs291466 | 2 | 191184475 | A | G | 0.62 | 0.03 | $7.34 \times 10^{-72}$ |
| HK2 | rs640944 | 2 | 75046839 | T | G | −0.22 | 0.04 | $1.73 \times 10^{-8}$ |
| HLA-DQA2 | rs557011 | 6 | 32587013 | C | T | 0.53 | 0.04 | $1.72 \times 10^{-42}$ |
| HMOX2 | rs79848476 | 16 | 4829602 | T | C | 0.37 | 0.07 | $2.74 \times 10^{-8}$ |
| HP | rs217184 | 16 | 72105965 | T | C | 0.82 | 0.05 | $8.95 \times 10^{-46}$ |
| HRG | rs9898 | 3 | 186390627 | C | T | 0.47 | 0.04 | $6.26 \times 10^{-27}$ |
| HSD17B14 | rs35299026 | 19 | 49318380 | G | A | −0.97 | 0.10 | $4.23 \times 10^{-22}$ |
| HSP90B1 | rs1177457 | 12 | 104336127 | C | T | 1.02 | 0.02 | $6.31 \times 10^{-199}$ |
| HYAL1 | rs78138837 | 3 | 50344178 | A | G | 0.72 | 0.07 | $7.47 \times 10^{-22}$ |
| IBSP | rs2627692 | 4 | 88649369 | C | T | −0.27 | 0.05 | $4.58 \times 10^{-8}$ |
| ICAM1 | rs5498 | 19 | 10395683 | A | G | −0.70 | 0.03 | $1.32 \times 10^{-106}$ |
| ICAM4 | rs901886 | 19 | 10402131 | T | C | 0.16 | 0.03 | $4.22 \times 10^{-9}$ |
| ICOSLG | rs112451903 | 21 | 45683369 | A | ACTG | −0.30 | 0.04 | $4.51 \times 10^{-14}$ |

| | | | | | | | | |
|---|---|---|---|---|---|---|---|---|
| IDUA | rs56079856 | 4 | 994011 | G | T | 0.90 | 0.06 | $1.62 \times 10^{-49}$ |
| IFNAR1 | rs2257167 | 21 | 34715699 | G | C | −1.16 | 0.03 | $3.14 \times 10^{-174}$ |
| IFNL1 | rs30461 | 19 | 39789115 | A | G | 0.43 | 0.06 | $3.12 \times 10^{-11}$ |
| IFNL3 | rs28416813 | 19 | 39735644 | C | G | −0.32 | 0.04 | $8.34 \times 10^{-15}$ |
| IFNLR1 | rs10903035 | 1 | 24481940 | G | A | 0.25 | 0.04 | $1.75 \times 10^{-9}$ |
| IGDCC3 | rs525514 | 15 | 65615556 | T | C | 0.39 | 0.03 | $9.08 \times 10^{-34}$ |
| IGDCC4 | rs8034057 | 15 | 65789430 | G | A | −0.55 | 0.09 | $3.70 \times 10^{-10}$ |
| IGF2R | rs629849 | 6 | 160494409 | A | G | 0.99 | 0.05 | $2.63 \times 10^{-75}$ |
| IGFBPL1 | rs148002733 | 9 | 38852172 | G | A | 0.58 | 0.05 | $1.75 \times 10^{-30}$ |
| IGFLR1 | rs12459634 | 19 | 36230174 | T | C | −0.64 | 0.04 | $5.28 \times 10^{-47}$ |
| IGHG1 | rs57446317 | 14 | 106206565 | C | G | 0.81 | 0.07 | $1.93 \times 10^{-29}$ |
| IGHG2 | rs57446317 | 14 | 106206565 | C | G | 0.81 | 0.07 | $1.93 \times 10^{-29}$ |
| IGHG3 | rs57446317 | 14 | 106206565 | C | G | 0.81 | 0.07 | $1.93 \times 10^{-29}$ |
| IGHG4 | rs57446317 | 14 | 106206565 | C | G | 0.81 | 0.07 | $1.93 \times 10^{-29}$ |
| IGSF11 | rs2903250 | 3 | 118649060 | G | T | 0.81 | 0.03 | $9.25 \times 10^{-138}$ |
| IL10RA | rs3135932 | 11 | 117864063 | A | G | −0.57 | 0.05 | $3.21 \times 10^{-24}$ |
| IL10RB | rs2515717 | 21 | 34662282 | G | A | 0.52 | 0.04 | $5.01 \times 10^{-31}$ |
| IL12B | rs2546890 | 5 | 158759900 | A | G | 0.51 | 0.05 | $2.20 \times 10^{-27}$ |
| IL15RA | rs8177641 | 10 | 6016892 | A | G | 0.59 | 0.04 | $6.70 \times 10^{-39}$ |
| IL16 | rs4778639 | 15 | 81600451 | T | G | −0.69 | 0.09 | $2.88 \times 10^{-15}$ |
| IL17D | rs2314714 | 13 | 21270132 | C | A | −0.53 | 0.09 | $2.32 \times 10^{-9}$ |
| IL17RA | rs3827279 | 22 | 17595929 | G | C | 0.72 | 0.05 | $6.31 \times 10^{-44}$ |
| IL17RB | rs62252978 | 3 | 53949262 | T | C | 0.37 | 0.06 | $2.99 \times 10^{-9}$ |
| IL17RC | rs6765503 | 3 | 9964826 | A | G | 0.39 | 0.04 | $1.55 \times 10^{-25}$ |
| IL17RD | rs35934471 | 3 | 57145830 | C | CA | −1.02 | 0.03 | $2.61 \times 10^{-140}$ |
| IL18R1 | rs12996505 | 2 | 102931802 | A | G | −1.01 | 0.02 | $1.64 \times 10^{-194}$ |
| IL1R1 | rs11689480 | 2 | 102674501 | G | A | 0.32 | 0.04 | $1.40 \times 10^{-18}$ |
| IL1RAP | rs6444442 | 3 | 190346060 | A | G | −1.45 | 0.05 | $1.61 \times 10^{-128}$ |
| IL1RL1 | rs7568913 | 2 | 102920037 | T | C | −1.05 | 0.03 | $2.86 \times 10^{-148}$ |
| IL1RL2 | rs1922292 | 2 | 102829013 | C | A | −0.39 | 0.04 | $3.23 \times 10^{-26}$ |
| IL1RN | rs56341434 | 2 | 113868990 | T | C | −0.25 | 0.04 | $4.45 \times 10^{-9}$ |
| IL20 | rs1150258 | 1 | 207074905 | T | C | −0.21 | 0.03 | $4.97 \times 10^{-10}$ |
| IL22 | rs4144961 | 12 | 68656647 | T | A | 0.69 | 0.04 | $3.51 \times 10^{-69}$ |

| | | | | | | | | |
|---|---|---|---|---|---|---|---|---|
| IL27RA | rs35026308 | 19 | 14153293 | T | C | −1.05 | 0.04 | $4.14 \times 10^{-118}$ |
| IL2RB | rs228953 | 22 | 37531436 | G | A | 0.30 | 0.04 | $1.43 \times 10^{-12}$ |
| IL34 | rs35794088 | 16 | 70666796 | C | T | −0.52 | 0.04 | $1.35 \times 10^{-30}$ |
| IL5RA | rs7619345 | 3 | 3138841 | G | A | 0.51 | 0.05 | $6.27 \times 10^{-27}$ |
| IL6 | rs10274260 | 7 | 22844846 | A | G | 0.44 | 0.05 | $1.22 \times 10^{-17}$ |
| IL6R | rs2228145 | 1 | 154426970 | A | C | 1.06 | 0.03 | $1.89 \times 10^{-194}$ |
| IL6ST | rs35257381 | 5 | 55318612 | A | T | 0.20 | 0.03 | $1.52 \times 10^{-10}$ |
| IL9 | rs31551 | 5 | 135275330 | G | A | 0.56 | 0.04 | $1.08 \times 10^{-43}$ |
| IMPAD1 | rs62511997 | 8 | 57859634 | C | T | −0.42 | 0.04 | $1.85 \times 10^{-21}$ |
| INPP5B | rs579689 | 1 | 38256166 | G | A | −0.52 | 0.04 | $1.91 \times 10^{-36}$ |
| INSR | rs4804774 | 19 | 7772173 | G | C | 0.19 | 0.03 | $1.91 \times 10^{-9}$ |
| IRF2 | rs793785 | 4 | 185368274 | A | C | 0.33 | 0.05 | $5.73 \times 10^{-12}$ |
| ISG15 | rs1921 | 1 | 949608 | G | A | 0.41 | 0.03 | $5.92 \times 10^{-36}$ |
| ITGB5 | rs34410053 | 3 | 124442738 | G | T | 0.30 | 0.05 | $1.80 \times 10^{-9}$ |
| ITIH4 | rs34092621 | 3 | 52887861 | A | AT | 0.22 | 0.04 | $8.61 \times 10^{-9}$ |
| ITIH5 | rs41298373 | 10 | 7622009 | G | A | −0.96 | 0.06 | $1.02 \times 10^{-49}$ |
| JAM2 | rs11909849 | 21 | 27168099 | C | G | −0.19 | 0.03 | $9.08 \times 10^{-11}$ |
| JAM3 | rs6590735 | 11 | 134022087 | G | T | 0.19 | 0.03 | $2.03 \times 10^{-8}$ |
| JAML | rs1540191 | 11 | 118090432 | G | A | 1.19 | 0.02 | $2.41 \times 10^{-240}$ |
| KDR | rs34495369 | 4 | 56122001 | G | A | −0.31 | 0.04 | $3.99 \times 10^{-13}$ |
| KIRREL3 | rs11220650 | 11 | 126760652 | A | G | −0.20 | 0.02 | $5.67 \times 10^{-26}$ |
| KLK10 | rs62115757 | 19 | 51521821 | T | G | −1.02 | 0.04 | $7.27 \times 10^{-103}$ |
| KLK11 | rs1048328 | 19 | 51527364 | G | A | −1.39 | 0.06 | $8.81 \times 10^{-87}$ |
| KLK12 | rs3745540 | 19 | 51535130 | A | G | −0.24 | 0.04 | $2.48 \times 10^{-8}$ |
| KLK13 | rs2569476 | 19 | 51569548 | C | T | −0.51 | 0.08 | $3.03 \times 10^{-11}$ |
| KLK14 | rs3810091 | 19 | 51568690 | C | T | 0.37 | 0.06 | $4.55 \times 10^{-10}$ |
| KLK15 | rs5519 | 19 | 51322312 | T | C | −0.65 | 0.04 | $4.41 \times 10^{-45}$ |
| KLK7 | rs1654526 | 19 | 51482459 | G | A | 0.93 | 0.05 | $1.25 \times 10^{-56}$ |
| KLK8 | rs2659074 | 19 | 51495579 | T | G | −0.34 | 0.04 | $2.90 \times 10^{-14}$ |
| KLKB1 | rs2304595 | 4 | 187172280 | G | A | 0.35 | 0.04 | $2.78 \times 10^{-19}$ |
| KLRB1 | rs2241006 | 12 | 9748209 | T | A | −0.71 | 0.04 | $3.06 \times 10^{-60}$ |
| KNG1 | rs5030049 | 3 | 186450863 | T | C | −0.91 | 0.06 | $7.80 \times 10^{-51}$ |
| KYNU | rs12477146 | 2 | 143793814 | G | A | −0.32 | 0.05 | $9.25 \times 10^{-12}$ |

| | | | | | | | | |
|---|---|---|---|---|---|---|---|---|
| LAMC2 | rs2276543 | 1 | 183155305 | G | A | 1.24 | 0.03 | $7.80 \times 10^{-208}$ |
| LAYN | rs663763 | 11 | 111422217 | C | T | 0.61 | 0.03 | $6.42 \times 10^{-89}$ |
| LBP | rs2232613 | 20 | 36997655 | C | T | −1.04 | 0.07 | $2.21 \times 10^{-40}$ |
| LCT | rs4988235 | 2 | 136608646 | G | A | 0.98 | 0.04 | $1.88 \times 10^{-114}$ |
| LCTL | rs7403574 | 15 | 66857290 | C | T | −0.40 | 0.06 | $1.45 \times 10^{-11}$ |
| LEAP2 | rs59414721 | 5 | 132322505 | T | TG | 0.41 | 0.05 | $3.64 \times 10^{-16}$ |
| LECT2 | rs31530 | 5 | 135282630 | C | T | −0.33 | 0.03 | $3.86 \times 10^{-22}$ |
| LEPR | rs12077336 | 1 | 66069986 | G | T | −1.38 | 0.03 | $4.25 \times 10^{-188}$ |
| LGALS2 | rs6000806 | 22 | 37971179 | T | C | −0.25 | 0.04 | $2.09 \times 10^{-10}$ |
| LGALS3 | rs76426991 | 14 | 55600939 | G | A | −1.22 | 0.07 | $2.43 \times 10^{-64}$ |
| LGALS8 | rs34299988 | 1 | 236701748 | C | T | 0.67 | 0.04 | $6.48 \times 10^{-66}$ |
| LGALS9 | rs4239242 | 17 | 25974258 | T | C | −0.33 | 0.04 | $8.74 \times 10^{-16}$ |
| LGMN | rs9791 | 14 | 93170993 | C | T | 0.27 | 0.04 | $8.19 \times 10^{-11}$ |
| LHB | rs78537284 | 19 | 49515363 | G | A | −0.96 | 0.07 | $2.60 \times 10^{-36}$ |
| LIFR | rs327287 | 5 | 38642552 | G | T | 0.18 | 0.03 | $2.41 \times 10^{-10}$ |
| LILRA4 | rs79828899 | 19 | 54853749 | C | T | 1.18 | 0.08 | $8.28 \times 10^{-42}$ |
| LILRA6 | rs11668526 | 19 | 54749060 | C | T | 0.64 | 0.04 | $3.27 \times 10^{-42}$ |
| LILRB1 | rs10427127 | 19 | 55143982 | T | C | −1.55 | 0.07 | $1.12 \times 10^{-86}$ |
| LILRB2 | rs7247451 | 19 | 54782704 | G | C | 0.62 | 0.04 | $3.33 \times 10^{-50}$ |
| LILRB5 | rs12975366 | 19 | 54759361 | T | C | −1.07 | 0.03 | $5.53 \times 10^{-180}$ |
| LMAN2L | rs72809820 | 2 | 97360079 | C | T | 0.29 | 0.02 | $1.05 \times 10^{-29}$ |
| LMOD1 | rs2644112 | 1 | 201806106 | T | C | −0.23 | 0.04 | $1.83 \times 10^{-8}$ |
| LPO | rs7219860 | 17 | 56321271 | G | A | −0.48 | 0.05 | $8.30 \times 10^{-23}$ |
| LRIT2 | rs4562751 | 10 | 85978723 | T | A | 0.41 | 0.05 | $1.47 \times 10^{-13}$ |
| LRP11 | rs7763718 | 6 | 150186534 | T | G | 0.39 | 0.02 | $1.11 \times 10^{-77}$ |
| LRP12 | rs28627996 | 8 | 105585028 | C | T | −0.22 | 0.04 | $7.06 \times 10^{-9}$ |
| LRP2 | rs10201691 | 2 | 170069028 | G | A | 0.43 | 0.07 | $1.27 \times 10^{-10}$ |
| LRP8 | rs79395289 | 1 | 53734037 | TG | T | 0.36 | 0.02 | $2.43 \times 10^{-68}$ |
| LRRC15 | rs6799819 | 3 | 194089549 | T | C | 0.70 | 0.07 | $1.37 \times 10^{-23}$ |
| LRRC32 | rs1320644 | 11 | 76370187 | G | A | −0.25 | 0.04 | $4.58 \times 10^{-9}$ |
| LRRN1 | rs35362954 | 3 | 3887508 | C | G | −0.54 | 0.05 | $5.60 \times 10^{-24}$ |
| LTF | rs55950019 | 3 | 46511644 | T | A | 1.11 | 0.03 | $3.78 \times 10^{-173}$ |
| LY86 | rs7757934 | 6 | 6578927 | G | A | 0.47 | 0.05 | $2.46 \times 10^{-20}$ |

| | | | | | | | | |
|---|---|---|---|---|---|---|---|---|
| LY9 | rs1333064 | 1 | 160763600 | G | A | 0.33 | 0.05 | $1.32 \times 10^{-12}$ |
| LY96 | rs2929505 | 8 | 74879827 | C | T | −0.29 | 0.05 | $1.17 \times 10^{-8}$ |
| LYPLAL1 | rs147082275 | 1 | 219454644 | TAAAG | T | 0.41 | 0.07 | $7.92 \times 10^{-10}$ |
| LYZ | rs4761234 | 12 | 69732105 | T | C | −0.74 | 0.03 | $4.58 \times 10^{-90}$ |
| MAGI2 | rs7779312 | 7 | 78116661 | G | A | 0.10 | 0.02 | $2.98 \times 10^{-10}$ |
| MAN2B2 | rs2301790 | 4 | 6600012 | A | G | 1.14 | 0.03 | $2.68 \times 10^{-167}$ |
| MANBA | rs223492 | 4 | 103675108 | G | C | 0.38 | 0.04 | $6.00 \times 10^{-24}$ |
| MANEA | rs35772543 | 6 | 96053922 | T | A | −1.59 | 0.05 | $3.36 \times 10^{-142}$ |
| MANF | rs6778196 | 3 | 51770429 | C | T | −0.38 | 0.06 | $3.96 \times 10^{-10}$ |
| MANSC4 | rs9668702 | 12 | 27919491 | T | G | 0.89 | 0.04 | $4.31 \times 10^{-87}$ |
| MAPK13 | rs12210904 | 6 | 36098191 | C | A | 0.35 | 0.04 | $2.78 \times 10^{-15}$ |
| MATN4 | rs11086956 | 20 | 43924106 | T | C | −0.59 | 0.04 | $1.85 \times 10^{-43}$ |
| MAX | rs10143198 | 14 | 65555471 | C | T | −0.23 | 0.03 | $4.07 \times 10^{-11}$ |
| MBL2 | rs7096206 | 10 | 54531685 | G | C | 0.79 | 0.05 | $5.04 \times 10^{-57}$ |
| MCEE | rs4852762 | 2 | 71373036 | C | T | −0.43 | 0.04 | $1.92 \times 10^{-24}$ |
| MESD | rs11855057 | 15 | 81282144 | C | G | −0.39 | 0.05 | $2.84 \times 10^{-15}$ |
| METTL24 | rs13218597 | 6 | 110679736 | A | G | 0.44 | 0.03 | $9.92 \times 10^{-36}$ |
| MFAP2 | rs2284746 | 1 | 17306675 | C | G | 0.64 | 0.04 | $4.24 \times 10^{-48}$ |
| MGP | rs2430737 | 12 | 15035563 | T | C | 0.35 | 0.04 | $1.40 \times 10^{-16}$ |
| MIA | rs2607412 | 19 | 41268243 | A | G | 0.73 | 0.08 | $5.47 \times 10^{-19}$ |
| MICA | rs9281428 | 6 | 31378227 | C | CT | −1.08 | 0.03 | $8.03 \times 10^{-154}$ |
| MICB | rs9266244 | 6 | 31325692 | G | A | 0.86 | 0.05 | $8.03 \times 10^{-49}$ |
| MIF | rs2070766 | 22 | 24237221 | C | G | −0.34 | 0.05 | $3.70 \times 10^{-12}$ |
| MLN | rs55775340 | 6 | 33751784 | C | G | −0.34 | 0.05 | $2.53 \times 10^{-10}$ |
| MMEL1 | rs4648652 | 1 | 2535758 | A | G | 1.16 | 0.03 | $5.28 \times 10^{-167}$ |
| MMP10 | rs486055 | 11 | 102650424 | C | T | −0.67 | 0.06 | $7.72 \times 10^{-28}$ |
| MMP12 | rs114176245 | 11 | 102719534 | C | T | −0.54 | 0.06 | $2.79 \times 10^{-16}$ |
| MMP2 | rs1561220 | 16 | 55504568 | A | G | 0.56 | 0.06 | $3.20 \times 10^{-21}$ |
| MMP7 | rs79643393 | 11 | 102410668 | C | T | 1.25 | 0.08 | $9.51 \times 10^{-47}$ |
| MMP8 | rs2155052 | 11 | 102595666 | C | G | −1.32 | 0.06 | $8.31 \times 10^{-84}$ |
| MPIG6B | rs11575845 | 6 | 31692386 | C | G | −1.14 | 0.08 | $7.17 \times 10^{-40}$ |
| MRC1 | rs201259350 | 10 | 17852064 | TAGAA | T | −0.38 | 0.05 | $5.62 \times 10^{-14}$ |

| | | | | | | | | |
|---|---|---|---|---|---|---|---|---|
| MRC2 | rs138105166 | 17 | 60704973 | TCCT CCCT CCGC | T | −0.33 | 0.04 | 9.58 × $10^{-15}$ |
| MSMB | rs10993994 | 10 | 51549496 | T | C | 0.85 | 0.03 | 1.37 × $10^{-117}$ |
| MSMP | rs10758322 | 9 | 35776190 | C | T | −0.59 | 0.04 | 9.41 × $10^{-52}$ |
| MST1 | rs11130213 | 3 | 49712297 | C | T | −1.12 | 0.03 | 4.99 × $10^{-187}$ |
| MTHFS | rs149183525 | 15 | 80168321 | AAAA T | A | −0.66 | 0.04 | 3.07 × $10^{-48}$ |
| MXRA7 | rs720782 | 17 | 74682602 | C | T | −0.35 | 0.03 | 1.05 × $10^{-36}$ |
| MXRA8 | rs307346 | 1 | 1260733 | A | C | 0.39 | 0.07 | 2.22 × $10^{-8}$ |
| NAAA | rs7686066 | 4 | 76838858 | A | T | −0.87 | 0.04 | 2.73 × $10^{-88}$ |
| NAGK | rs2287327 | 2 | 71297982 | C | T | −0.29 | 0.03 | 1.41 × $10^{-17}$ |
| NAGPA | rs12599777 | 16 | 5079466 | A | G | −0.36 | 0.04 | 2.32 × $10^{-20}$ |
| NBL1 | rs2854108 | 1 | 19973920 | A | G | −0.32 | 0.02 | 1.32 × $10^{-45}$ |
| NCAM2 | rs2009029 | 21 | 22855892 | A | G | 0.32 | 0.03 | 4.61 × $10^{-21}$ |
| NCR1 | rs77273876 | 19 | 55400971 | G | A | 1.23 | 0.08 | 5.52 × $10^{-44}$ |
| NCR3 | rs986475 | 6 | 31556709 | A | G | −0.56 | 0.06 | 6.56 × $10^{-22}$ |
| NDNF | rs6840113 | 4 | 121936507 | A | G | 0.49 | 0.04 | 5.48 × $10^{-31}$ |
| NDST1 | rs2545341 | 5 | 149914401 | T | C | 0.22 | 0.03 | 1.01 × $10^{-14}$ |
| NELL1 | rs79474191 | 11 | 20953407 | C | T | 1.16 | 0.08 | 2.57 × $10^{-45}$ |
| NID1 | rs9662380 | 1 | 236178606 | G | A | −0.34 | 0.05 | 4.20 × $10^{-13}$ |
| NID2 | rs140170580 | 14 | 52494307 | T | TA | −0.70 | 0.05 | 3.36 × $10^{-40}$ |
| NMB | rs34452033 | 15 | 85221993 | A | G | −0.33 | 0.04 | 5.02 × $10^{-17}$ |
| NPNT | rs34712979 | 4 | 106819053 | G | A | −0.35 | 0.05 | 8.20 × $10^{-12}$ |
| NPPB | rs12402728 | 1 | 11922148 | A | G | 0.53 | 0.05 | 8.81 × $10^{-28}$ |
| NPW | rs3785284 | 16 | 2074036 | A | G | −0.78 | 0.04 | 1.19 × $10^{-65}$ |
| NQO1 | rs112668868 | 16 | 69713957 | C | G | −1.00 | 0.04 | 2.79 × $10^{-93}$ |
| NQO2 | rs2756078 | 6 | 3010103 | G | A | 0.98 | 0.04 | 9.61 × $10^{-102}$ |
| NRG1 | rs10089448 | 8 | 32597355 | C | A | −0.50 | 0.06 | 1.25 × $10^{-14}$ |
| NRG4 | rs35468194 | 15 | 76349565 | T | TA | 1.26 | 0.03 | 2.96 × $10^{-169}$ |
| NRP1 | rs2506149 | 10 | 33480713 | C | T | −0.33 | 0.03 | 7.98 × $10^{-23}$ |
| NRP2 | rs11678440 | 2 | 206548443 | T | C | −0.17 | 0.03 | 4.32 × $10^{-8}$ |
| NT5C | rs111346516 | 17 | 73603700 | G | A | −0.40 | 0.06 | 2.70 × $10^{-11}$ |
| NTM | rs12790269 | 11 | 131148713 | G | A | 0.13 | 0.02 | 6.33 × $10^{-15}$ |
| NTN1 | rs940854 | 17 | 8934763 | C | T | −0.28 | 0.04 | 2.32 × $10^{-13}$ |

| | | | | | | | | |
|---|---|---|---|---|---|---|---|---|
| NTN4 | rs7959545 | 12 | 96059957 | G | C | 0.80 | 0.05 | $5.31 \times 10^{-46}$ |
| NTRK1 | rs2365715 | 1 | 156615114 | A | G | −0.43 | 0.04 | $1.01 \times 10^{-22}$ |
| NUCB1 | rs2017135 | 19 | 49383269 | C | T | −0.44 | 0.06 | $3.37 \times 10^{-12}$ |
| NUDCD3 | rs306997 | 7 | 44538882 | A | G | 0.45 | 0.03 | $2.74 \times 10^{-37}$ |
| NUDT9 | rs58601962 | 4 | 88384477 | A | AC | −0.51 | 0.06 | $3.06 \times 10^{-18}$ |
| OAF | rs2508490 | 11 | 120099679 | G | A | −0.87 | 0.05 | $1.72 \times 10^{-58}$ |
| OAS1 | rs10850097 | 12 | 113361117 | C | T | −0.48 | 0.04 | $1.07 \times 10^{-34}$ |
| OLFM1 | rs546682 | 9 | 137970302 | C | T | −0.22 | 0.03 | $4.97 \times 10^{-12}$ |
| OLFM2 | rs1862474 | 19 | 10096650 | T | C | 0.42 | 0.04 | $6.33 \times 10^{-21}$ |
| OLFML3 | rs3841011 | 1 | 114521683 | A | AAAC | 0.34 | 0.05 | $2.24 \times 10^{-12}$ |
| OLR1 | rs17808009 | 12 | 10311929 | C | T | 0.18 | 0.03 | $7.39 \times 10^{-11}$ |
| OMD | rs117800660 | 9 | 95174368 | A | T | −0.47 | 0.08 | $9.14 \times 10^{-9}$ |
| OSCAR | rs254252 | 19 | 54594864 | G | A | 0.52 | 0.08 | $9.23 \times 10^{-11}$ |
| OSMR | rs72732754 | 5 | 38999995 | A | T | −0.31 | 0.04 | $1.19 \times 10^{-11}$ |
| PAM | rs34274728 | 5 | 102306489 | CT | C | −0.47 | 0.03 | $1.56 \times 10^{-40}$ |
| PCBD1 | rs2630336 | 10 | 72648422 | G | T | −0.26 | 0.04 | $5.40 \times 10^{-13}$ |
| PCDH10 | rs1112105 | 4 | 133894412 | T | G | 0.21 | 0.03 | $2.30 \times 10^{-14}$ |
| PCOLCE2 | rs2707975 | 3 | 142611657 | A | G | 0.78 | 0.05 | $5.27 \times 10^{-50}$ |
| PCSK1 | rs6234 | 5 | 95728974 | G | C | −0.83 | 0.03 | $9.39 \times 10^{-122}$ |
| PCSK7 | rs236910 | 11 | 117081974 | A | G | −1.13 | 0.03 | $2.24 \times 10^{-151}$ |
| PCSK9 | rs41294819 | 1 | 55504586 | A | G | −0.85 | 0.05 | $1.39 \times 10^{-49}$ |
| PDCD1LG2 | rs62556118 | 9 | 5515990 | G | C | 0.53 | 0.04 | $2.60 \times 10^{-41}$ |
| PDCD5 | rs4499344 | 19 | 33073431 | G | A | 0.25 | 0.03 | $1.17 \times 10^{-13}$ |
| PDGFD | rs10444324 | 11 | 104095391 | A | C | 0.42 | 0.04 | $8.55 \times 10^{-21}$ |
| PDGFRB | rs3816018 | 5 | 149508475 | C | T | 1.14 | 0.02 | $4.59 \times 10^{-308}$ |
| PDGFRL | rs2720579 | 8 | 17432466 | G | A | −0.83 | 0.05 | $1.06 \times 10^{-51}$ |
| PDLIM4 | rs4877 | 5 | 131607588 | G | T | −0.38 | 0.05 | $2.34 \times 10^{-14}$ |
| PENK | rs1877571 | 8 | 57426778 | A | C | 0.46 | 0.04 | $3.19 \times 10^{-26}$ |
| PGD | rs72867415 | 1 | 10416602 | G | A | −0.68 | 0.08 | $1.52 \times 10^{-17}$ |
| PIGR | rs533494 | 1 | 207117575 | C | T | 0.42 | 0.05 | $1.57 \times 10^{-16}$ |
| PILRA | rs1859788 | 7 | 99971834 | A | G | −1.29 | 0.03 | $1.02 \times 10^{-204}$ |
| PKDCC | rs10495892 | 2 | 42156206 | G | C | 0.24 | 0.04 | $4.13 \times 10^{-8}$ |
| PLA2G2A | rs11573156 | 1 | 20306146 | G | C | 0.29 | 0.05 | $7.72 \times 10^{-9}$ |

| | | | | | | | | |
|---|---|---|---|---|---|---|---|---|
| PLA2G7 | rs1421378 | 6 | 46703513 | A | G | 0.79 | 0.03 | $1.19 \times 10^{-94}$ |
| PLA2R1 | rs3749117 | 2 | 160885442 | T | C | −0.97 | 0.03 | $3.91 \times 10^{-122}$ |
| PLAUR | rs4760 | 19 | 44153100 | A | G | −0.26 | 0.03 | $2.00 \times 10^{-14}$ |
| PLD5 | rs72761220 | 1 | 242267176 | A | T | 0.32 | 0.05 | $2.30 \times 10^{-12}$ |
| PLEK | rs3816281 | 2 | 68607947 | G | T | 0.36 | 0.04 | $4.56 \times 10^{-17}$ |
| PLOD2 | rs1707466 | 3 | 145850024 | G | A | −0.36 | 0.04 | $3.70 \times 10^{-20}$ |
| PLXDC2 | rs7094178 | 10 | 20105223 | G | A | 0.27 | 0.03 | $3.81 \times 10^{-18}$ |
| PLXNB2 | rs28573806 | 22 | 50727792 | T | C | −1.22 | 0.02 | $2.23 \times 10^{-308}$ |
| PLXNC1 | rs12313790 | 12 | 94632933 | C | A | −0.39 | 0.06 | $5.99 \times 10^{-10}$ |
| PNLIPRP2 | rs7910135 | 10 | 118398046 | C | A | 0.82 | 0.03 | $4.86 \times 10^{-93}$ |
| POFUT1 | rs76143353 | 20 | 30815755 | C | T | −1.11 | 0.09 | $7.73 \times 10^{-33}$ |
| POGLUT1 | rs6794833 | 3 | 119191398 | T | C | −0.65 | 0.04 | $2.74 \times 10^{-62}$ |
| POGLUT3 | rs111559044 | 11 | 108254454 | G | C | −0.69 | 0.09 | $2.89 \times 10^{-13}$ |
| POMC | rs3754861 | 2 | 25393722 | A | C | 0.37 | 0.06 | $7.04 \times 10^{-11}$ |
| POMGNT2 | rs2002182 | 3 | 43147538 | T | G | −0.26 | 0.04 | $8.36 \times 10^{-12}$ |
| POSTN | rs9547910 | 13 | 38069999 | G | A | −0.58 | 0.04 | $3.79 \times 10^{-43}$ |
| PPID | rs6856561 | 4 | 159621185 | A | T | 0.43 | 0.03 | $1.29 \times 10^{-31}$ |
| PPIE | rs1046988 | 1 | 40219065 | C | T | −0.45 | 0.03 | $4.74 \times 10^{-42}$ |
| PPIF | rs11002931 | 10 | 81094251 | A | C | −0.47 | 0.04 | $3.83 \times 10^{-24}$ |
| PPT1 | rs112524162 | 1 | 40576715 | G | T | −0.93 | 0.10 | $1.26 \times 10^{-20}$ |
| PRKCSH | rs160839 | 19 | 11557697 | T | C | −0.25 | 0.04 | $2.07 \times 10^{-11}$ |
| PROS1 | rs5013930 | 3 | 93679707 | T | C | 0.35 | 0.05 | $1.28 \times 10^{-10}$ |
| PRTN3 | rs6510982 | 19 | 845535 | C | G | 0.33 | 0.04 | $2.47 \times 10^{-14}$ |
| PSG3 | rs4030933 | 19 | 43237764 | T | G | −0.42 | 0.04 | $1.16 \times 10^{-24}$ |
| PSG9 | rs140022160 | 19 | 43784407 | C | CCCTT | 0.40 | 0.05 | $2.52 \times 10^{-13}$ |
| PSMB1 | rs3734763 | 6 | 170885912 | T | C | −0.36 | 0.03 | $6.32 \times 10^{-37}$ |
| PSPN | rs6510895 | 19 | 6391847 | C | T | 0.43 | 0.04 | $1.23 \times 10^{-20}$ |
| PTGFRN | rs4233450 | 1 | 117490261 | G | T | 0.87 | 0.04 | $1.08 \times 10^{-78}$ |
| PTGR1 | rs56912703 | 9 | 114357659 | T | A | −0.42 | 0.04 | $8.62 \times 10^{-21}$ |
| PTH | rs78157059 | 11 | 13691962 | C | G | −0.45 | 0.07 | $9.70 \times 10^{-12}$ |
| PTHLH | rs10843110 | 12 | 28289141 | T | G | −0.29 | 0.05 | $3.76 \times 10^{-10}$ |
| PTPN4 | rs11896956 | 2 | 120135332 | A | G | −0.49 | 0.06 | $4.95 \times 10^{-17}$ |
| PTPRU | rs212297 | 1 | 29723516 | C | T | −0.42 | 0.03 | $8.55 \times 10^{-37}$ |

| | | | | | | | | |
|---|---|---|---|---|---|---|---|---|
| PXYLP1 | rs6786821 | 3 | 140999276 | C | G | 0.47 | 0.04 | $1.14 \times 10^{-25}$ |
| QDPR | rs4698602 | 4 | 17519712 | G | A | −0.36 | 0.04 | $2.29 \times 10^{-15}$ |
| QPCT | rs12467820 | 2 | 37572806 | G | T | −0.46 | 0.03 | $2.63 \times 10^{-51}$ |
| QPCTL | rs2110574 | 19 | 46197421 | C | T | 0.16 | 0.03 | $1.27 \times 10^{-9}$ |
| QSOX1 | rs12371 | 1 | 180163390 | A | G | −0.52 | 0.05 | $3.82 \times 10^{-28}$ |
| QSOX2 | rs3758199 | 9 | 139097645 | C | T | −0.25 | 0.03 | $2.63 \times 10^{-14}$ |
| RABEPK | rs147332108 | 9 | 127911299 | TA | T | 1.05 | 0.04 | $1.72 \times 10^{-107}$ |
| RAET1L | rs6557222 | 6 | 150300478 | A | G | −0.46 | 0.05 | $3.67 \times 10^{-18}$ |
| RARRES1 | rs6441224 | 3 | 158450417 | T | C | 0.42 | 0.05 | $8.46 \times 10^{-19}$ |
| RARRES2 | rs28432021 | 7 | 150046235 | G | C | 0.72 | 0.05 | $1.41 \times 10^{-45}$ |
| RBP7 | rs3811458 | 1 | 10057203 | G | T | −0.61 | 0.06 | $1.90 \times 10^{-22}$ |
| REG1A | rs76841471 | 2 | 79330169 | T | G | 0.50 | 0.08 | $2.91 \times 10^{-10}$ |
| REG3G | rs436758 | 2 | 79247510 | T | C | 0.67 | 0.04 | $2.84 \times 10^{-45}$ |
| REG4 | rs58163904 | 1 | 120365410 | GCA | G | −0.45 | 0.05 | $5.07 \times 10^{-19}$ |
| RGMA | rs11858429 | 15 | 93844067 | C | T | 0.21 | 0.03 | $2.87 \times 10^{-11}$ |
| RIDA | rs7846130 | 8 | 99109401 | A | G | −0.23 | 0.02 | $1.61 \times 10^{-24}$ |
| RMDN1 | rs10107571 | 8 | 87518985 | G | T | −1.04 | 0.02 | $6.71 \times 10^{-229}$ |
| RNASE2 | rs2233859 | 14 | 21359808 | C | A | 0.63 | 0.04 | $4.16 \times 10^{-59}$ |
| RNASE3 | rs11299601 | 14 | 21349400 | AG | A | −0.67 | 0.05 | $3.39 \times 10^{-38}$ |
| RNASE4 | rs944438 | 14 | 21164627 | A | G | −0.32 | 0.04 | $4.98 \times 10^{-15}$ |
| RNASE6 | rs1045922 | 14 | 21250124 | G | A | 1.23 | 0.03 | $8.93 \times 10^{-240}$ |
| ROBO2 | rs74506149 | 3 | 77580700 | A | C | −0.28 | 0.04 | $3.28 \times 10^{-12}$ |
| ROBO3 | rs55706177 | 11 | 124746180 | C | T | 0.38 | 0.05 | $1.44 \times 10^{-14}$ |
| ROR1 | rs61765448 | 1 | 64611828 | C | T | −0.45 | 0.04 | $2.18 \times 10^{-24}$ |
| ROR2 | rs4744097 | 9 | 94543365 | G | A | 0.25 | 0.03 | $7.41 \times 10^{-15}$ |
| RPE | rs2887872 | 2 | 210878117 | C | T | −0.25 | 0.03 | $6.63 \times 10^{-19}$ |
| RPN1 | rs2712419 | 3 | 128340896 | T | C | −0.51 | 0.02 | $1.79 \times 10^{-79}$ |
| RRM2B | rs2305832 | 8 | 103266805 | C | A | 0.52 | 0.06 | $2.43 \times 10^{-15}$ |
| RSPO2 | rs593872 | 8 | 108941705 | G | A | 0.54 | 0.04 | $2.56 \times 10^{-42}$ |
| RSPO4 | rs4813028 | 20 | 1096630 | T | C | 0.36 | 0.06 | $2.49 \times 10^{-8}$ |
| S100A4 | rs1005436 | 1 | 153521932 | G | A | 0.55 | 0.07 | $3.96 \times 10^{-15}$ |
| S100A7 | rs3006490 | 1 | 153369009 | A | C | −1.07 | 0.09 | $1.86 \times 10^{-28}$ |

| | | | | | | | | |
|---|---|---|---|---|---|---|---|---|
| SAA1 | rs10690148 | 11 | 18293504 | C | CAACT T | −0.94 | 0.04 | $4.53 \times 10^{-87}$ |
| SAT2 | rs11078700 | 17 | 7511936 | A | G | −0.55 | 0.07 | $1.50 \times 10^{-14}$ |
| SCARA3 | rs2640734 | 8 | 27532654 | C | T | −0.31 | 0.05 | $3.08 \times 10^{-9}$ |
| SCARB2 | rs78466149 | 4 | 76393394 | G | A | −0.31 | 0.06 | $4.95 \times 10^{-8}$ |
| SCARF1 | rs145220082 | 17 | 1545567 | T | C | 1.44 | 0.05 | $9.67 \times 10^{-116}$ |
| SCARF2 | rs10606665 | 22 | 20776973 | AACC | A | 0.32 | 0.05 | $1.55 \times 10^{-9}$ |
| SCG3 | rs1456297 | 15 | 51964210 | C | A | 0.34 | 0.04 | $4.77 \times 10^{-14}$ |
| SCGN | rs4419666 | 6 | 25693274 | C | T | 0.40 | 0.05 | $5.65 \times 10^{-18}$ |
| SCT | rs79177795 | 11 | 644748 | C | G | 0.70 | 0.06 | $8.66 \times 10^{-26}$ |
| SCUBE1 | rs138993 | 22 | 43610207 | A | G | 0.25 | 0.04 | $1.95 \times 10^{-8}$ |
| SELL | rs2223286 | 1 | 169665632 | T | C | −0.66 | 0.04 | $3.99 \times 10^{-51}$ |
| SELP | rs6128 | 1 | 169562904 | C | T | −0.66 | 0.05 | $3.16 \times 10^{-33}$ |
| SEMA3E | rs3801487 | 7 | 83033915 | T | C | −1.19 | 0.05 | $1.99 \times 10^{-109}$ |
| SEMA3G | rs34135146 | 3 | 52313432 | G | C | −0.29 | 0.04 | $1.73 \times 10^{-11}$ |
| SEMA4D | rs45464494 | 9 | 91994433 | C | T | −0.79 | 0.05 | $2.89 \times 10^{-47}$ |
| SERPINA1 | rs1243167 | 14 | 94841641 | G | A | 0.28 | 0.05 | $2.40 \times 10^{-8}$ |
| SERPINA4 | rs10140765 | 14 | 95023389 | C | T | 0.36 | 0.04 | $1.57 \times 10^{-16}$ |
| SERPINA5 | rs885786 | 14 | 95050496 | C | G | 0.40 | 0.04 | $2.43 \times 10^{-20}$ |
| SERPINA9 | rs12896629 | 14 | 94940361 | G | T | 0.16 | 0.03 | $3.39 \times 10^{-9}$ |
| SERPINB1 | rs386713 | 6 | 2842139 | C | T | −0.40 | 0.04 | $2.39 \times 10^{-27}$ |
| SERPIND1 | rs6004023 | 22 | 21065285 | A | G | 0.38 | 0.04 | $1.80 \times 10^{-19}$ |
| SERPINE1 | rs2227674 | 7 | 100776208 | A | G | 0.32 | 0.05 | $3.88 \times 10^{-9}$ |
| SERPINF1 | rs12450371 | 17 | 1667674 | C | T | −0.51 | 0.04 | $1.93 \times 10^{-29}$ |
| SERPING1 | rs11229075 | 11 | 57391023 | A | C | 0.84 | 0.04 | $1.20 \times 10^{-82}$ |
| SFRP4 | rs17236800 | 7 | 37944375 | A | G | −0.33 | 0.05 | $8.07 \times 10^{-10}$ |
| SHBG | rs12452603 | 17 | 7504977 | T | C | 0.30 | 0.05 | $1.19 \times 10^{-8}$ |
| SIGLEC12 | rs4801871 | 19 | 52002317 | G | C | −1.35 | 0.04 | $1.01 \times 10^{-141}$ |
| SIGLEC14 | rs147859411 | 19 | 52130148 | T | TTTTT G | −1.12 | 0.06 | $2.60 \times 10^{-62}$ |
| SIGLEC5 | rs147859411 | 19 | 52130148 | T | TTTTT G | −1.16 | 0.06 | $5.75 \times 10^{-78}$ |
| SIGLEC6 | rs2305771 | 19 | 52033572 | T | C | 0.31 | 0.05 | $6.18 \times 10^{-11}$ |
| SIGLEC7 | rs12983058 | 19 | 51642784 | G | T | 1.20 | 0.03 | $1.44 \times 10^{-170}$ |
| SIGLEC9 | rs2075803 | 19 | 51628529 | A | G | −1.18 | 0.02 | $1.09 \times 10^{-240}$ |

| | | | | | | | | |
|---|---|---|---|---|---|---|---|---|
| SIRPA | rs6075340 | 20 | 1894335 | G | A | −1.17 | 0.03 | $5.81 \times 10^{-196}$ |
| SIRPB1 | rs2318043 | 20 | 1596473 | A | G | 1.27 | 0.04 | $4.68 \times 10^{-145}$ |
| SIRPG | rs17855609 | 20 | 1895813 | A | T | −0.60 | 0.05 | $1.87 \times 10^{-34}$ |
| SLAMF7 | rs11581248 | 1 | 160720074 | C | T | −1.21 | 0.06 | $2.92 \times 10^{-74}$ |
| SMOC1 | rs12101270 | 14 | 70463688 | T | C | −0.41 | 0.04 | $2.46 \times 10^{-19}$ |
| SMPDL3A | rs9385271 | 6 | 123125411 | T | C | −0.82 | 0.04 | $3.29 \times 10^{-72}$ |
| SOD3 | rs11938550 | 4 | 24726263 | G | C | −0.68 | 0.04 | $9.29 \times 10^{-54}$ |
| SORBS3 | rs1047030 | 8 | 22428708 | A | G | −0.36 | 0.05 | $8.98 \times 10^{-12}$ |
| SORCS2 | rs3892041 | 4 | 7314962 | A | C | 0.36 | 0.05 | $1.02 \times 10^{-12}$ |
| SOST | rs9899889 | 17 | 41823225 | T | G | −0.55 | 0.04 | $2.32 \times 10^{-39}$ |
| SPARCL1 | rs17012853 | 4 | 88454184 | C | T | −0.35 | 0.05 | $2.82 \times 10^{-13}$ |
| SPAST | rs79204217 | 2 | 31902450 | T | C | 0.23 | 0.04 | $2.04 \times 10^{-8}$ |
| SPATA20 | rs8076632 | 17 | 48625928 | C | G | −0.50 | 0.04 | $2.73 \times 10^{-33}$ |
| SPINK2 | rs781538 | 4 | 57685669 | C | T | 0.52 | 0.04 | $4.30 \times 10^{-41}$ |
| SPINK6 | rs765776722 | 5 | 147592991 | T | TG | −1.39 | 0.07 | $2.38 \times 10^{-75}$ |
| SPINT1 | rs17658212 | 15 | 41145919 | C | T | −0.68 | 0.08 | $1.24 \times 10^{-16}$ |
| SPINT2 | rs71354995 | 19 | 38791841 | A | G | −1.20 | 0.03 | $1.99 \times 10^{-207}$ |
| SPINT3 | rs6017593 | 20 | 44147814 | A | G | 0.25 | 0.03 | $1.07 \times 10^{-13}$ |
| SPOCK2 | rs1245550 | 10 | 73844081 | A | G | −0.56 | 0.04 | $3.06 \times 10^{-39}$ |
| SPON1 | rs1528661 | 11 | 13971798 | T | C | −0.43 | 0.03 | $1.78 \times 10^{-31}$ |
| SPON2 | rs878323 | 4 | 1169813 | G | T | −0.59 | 0.04 | $1.73 \times 10^{-39}$ |
| ST3GAL1 | rs2142306 | 8 | 134470631 | T | C | 0.29 | 0.04 | $1.72 \times 10^{-15}$ |
| ST3GAL6 | rs113325534 | 3 | 98786038 | C | T | −1.45 | 0.08 | $9.24 \times 10^{-64}$ |
| ST6GALNAC1 | rs2286595 | 17 | 74621635 | C | T | 0.20 | 0.03 | $1.78 \times 10^{-8}$ |
| ST6GALNAC2 | rs10852772 | 17 | 74580421 | G | A | −0.48 | 0.05 | $4.92 \times 10^{-20}$ |
| ST8SIA2 | rs34728194 | 15 | 93064556 | A | C | −0.23 | 0.04 | $6.50 \times 10^{-9}$ |
| STC1 | rs138694283 | 8 | 23818092 | C | CT | 0.54 | 0.09 | $2.57 \times 10^{-9}$ |
| STX8 | rs11078801 | 17 | 9337245 | G | C | 0.12 | 0.02 | $1.00 \times 10^{-11}$ |
| SVEP1 | rs74308641 | 9 | 113313412 | G | A | −0.58 | 0.08 | $1.08 \times 10^{-13}$ |
| TAC1 | rs10272589 | 7 | 96896639 | G | A | 0.65 | 0.06 | $3.60 \times 10^{-29}$ |
| TAPBP | rs469064 | 6 | 33250476 | C | A | −0.32 | 0.04 | $3.17 \times 10^{-16}$ |
| TAPBPL | rs2041387 | 12 | 6562823 | C | T | 1.32 | 0.03 | $5.04 \times 10^{-239}$ |

| | | | | | | | | |
|---|---|---|---|---|---|---|---|---|
| TAX1BP3 | rs2873624 | 17 | 3563963 | C | G | −0.25 | 0.04 | $1.86 \times 10^{-10}$ |
| TBCE | rs868815 | 1 | 235584543 | C | G | −0.18 | 0.03 | $3.87 \times 10^{-11}$ |
| TBL2 | rs76029572 | 7 | 72992858 | C | G | 1.28 | 0.08 | $2.03 \times 10^{-51}$ |
| TCN2 | rs12169610 | 22 | 31022590 | C | T | −0.72 | 0.06 | $6.36 \times 10^{-28}$ |
| TDGF1 | rs80045772 | 3 | 46619238 | T | A | 1.22 | 0.03 | $9.62 \times 10^{-237}$ |
| TEK | rs16911092 | 9 | 27142444 | A | G | 0.98 | 0.07 | $2.82 \times 10^{-36}$ |
| TFF1 | rs3761376 | 21 | 43787038 | G | A | −0.32 | 0.05 | $4.15 \times 10^{-9}$ |
| TGFB1 | rs57948035 | 19 | 41882044 | CAAA<br>A | C | 0.23 | 0.03 | $3.81 \times 10^{-14}$ |
| TGFB2 | rs7533619 | 1 | 219346409 | C | T | −0.21 | 0.04 | $5.04 \times 10^{-9}$ |
| TGFB3 | rs11621464 | 14 | 76379798 | C | G | 0.43 | 0.06 | $8.35 \times 10^{-14}$ |
| TGM3 | rs214830 | 20 | 2321105 | G | C | 0.42 | 0.05 | $5.78 \times 10^{-18}$ |
| THBS2 | rs7341189 | 6 | 169622734 | G | A | 0.26 | 0.05 | $1.68 \times 10^{-8}$ |
| THBS3 | rs4072037 | 1 | 155162067 | C | T | −0.34 | 0.04 | $2.53 \times 10^{-14}$ |
| THBS4 | rs256438 | 5 | 79366249 | T | G | 0.53 | 0.04 | $4.95 \times 10^{-33}$ |
| THSD1 | rs191768 | 13 | 53152704 | T | C | −0.46 | 0.08 | $2.72 \times 10^{-8}$ |
| TIE1 | rs1199039 | 1 | 43784956 | A | G | −0.24 | 0.03 | $1.68 \times 10^{-12}$ |
| TIGAR | rs143427116 | 12 | 4430340 | AGCC<br>GGCC<br>GGCT | A | 0.60 | 0.07 | $1.37 \times 10^{-15}$ |
| TLR4 | rs4986791 | 9 | 120475602 | C | T | −1.05 | 0.06 | $5.43 \times 10^{-65}$ |
| TMED2 | rs786430 | 12 | 124087988 | G | C | −0.34 | 0.03 | $4.04 \times 10^{-33}$ |
| TMEM106B | rs4721060 | 7 | 12267552 | G | A | −0.37 | 0.03 | $2.15 \times 10^{-42}$ |
| TMEM132A | rs55920775 | 11 | 60703777 | G | A | 0.45 | 0.03 | $2.41 \times 10^{-57}$ |
| TMEM132C | rs7313554 | 12 | 128791042 | T | C | 0.16 | 0.02 | $6.27 \times 10^{-11}$ |
| TMEM132D | rs12372807 | 12 | 129561332 | G | A | 0.22 | 0.04 | $1.25 \times 10^{-8}$ |
| TMEM190 | rs4806666 | 19 | 55888095 | C | T | −1.14 | 0.03 | $2.15 \times 10^{-172}$ |
| TMEM25 | rs200268771 | 11 | 118446875 | G | GT | −0.51 | 0.09 | $2.54 \times 10^{-9}$ |
| TMEM9 | rs2365296 | 1 | 201114196 | C | T | −0.77 | 0.02 | $3.62 \times 10^{-217}$ |
| TMPRSS5 | rs11315692 | 11 | 113616733 | TG | T | −0.62 | 0.04 | $2.06 \times 10^{-51}$ |
| TNC | rs1138545 | 9 | 117835899 | C | T | 1.38 | 0.06 | $2.85 \times 10^{-100}$ |
| TNFAIP6 | rs34026490 | 2 | 152205792 | T | C | 0.33 | 0.04 | $5.93 \times 10^{-15}$ |
| TNFRSF10B | rs4871844 | 8 | 22879734 | T | C | 0.45 | 0.03 | $1.36 \times 10^{-41}$ |
| TNFRSF11A | rs884205 | 18 | 60054857 | A | C | −0.43 | 0.03 | $1.93 \times 10^{-41}$ |
| TNFRSF14 | rs2281852 | 1 | 2490942 | C | A | 0.36 | 0.02 | $1.24 \times 10^{-56}$ |

| | | | | | | | | |
|---|---|---|---|---|---|---|---|---|
| TNFRSF18 | rs12066716 | 1 | 1123434 | T | A | −0.43 | 0.04 | $4.00 \times 10^{-21}$ |
| TNFRSF19 | rs3751364 | 13 | 24234517 | A | G | 0.27 | 0.04 | $4.70 \times 10^{-11}$ |
| TNFRSF1B | rs616645 | 1 | 12240824 | T | G | −0.25 | 0.04 | $7.23 \times 10^{-9}$ |
| TNFRSF4 | rs190796582 | 1 | 1167796 | C | T | 0.60 | 0.07 | $1.00 \times 10^{-16}$ |
| TNFRSF8 | rs3830887 | 1 | 12066218 | T | TA | 0.50 | 0.04 | $2.72 \times 10^{-34}$ |
| TNFSF13B | rs1224148 | 13 | 108956794 | A | G | 0.40 | 0.05 | $5.00 \times 10^{-14}$ |
| TNFSF15 | rs7848647 | 9 | 117569046 | T | C | −0.53 | 0.04 | $4.25 \times 10^{-41}$ |
| TNFSF8 | rs4979474 | 9 | 117695984 | G | A | −0.26 | 0.04 | $8.36 \times 10^{-9}$ |
| TNXB | rs201459441 | 6 | 31962708 | AC | A | −0.62 | 0.09 | $1.35 \times 10^{-11}$ |
| TPSAB1 | rs112332886 | 16 | 1288563 | G | A | 1.20 | 0.07 | $5.25 \times 10^{-50}$ |
| TPSB2 | rs112332886 | 16 | 1288563 | G | A | 1.37 | 0.07 | $2.28 \times 10^{-73}$ |
| TREM1 | rs9462692 | 6 | 41262236 | A | C | −0.72 | 0.05 | $1.67 \times 10^{-47}$ |
| TRH | rs2670897 | 3 | 129699722 | C | T | 0.64 | 0.05 | $4.18 \times 10^{-33}$ |
| TRIL | rs3735562 | 7 | 28996557 | C | T | −0.47 | 0.05 | $1.06 \times 10^{-20}$ |
| TXNDC15 | rs3733897 | 5 | 134223593 | A | G | 1.10 | 0.06 | $3.45 \times 10^{-72}$ |
| TXNRD1 | rs11111979 | 12 | 104680782 | C | G | −0.13 | 0.02 | $4.15 \times 10^{-9}$ |
| TYMP | rs140522 | 22 | 50971266 | T | C | 0.29 | 0.03 | $8.16 \times 10^{-21}$ |
| UCMA | rs2399954 | 10 | 13276368 | G | A | −0.85 | 0.05 | $3.08 \times 10^{-53}$ |
| ULBP2 | rs6557222 | 6 | 150300478 | A | G | −0.35 | 0.05 | $7.33 \times 10^{-11}$ |
| ULBP3 | rs12661513 | 6 | 150373839 | C | A | −1.01 | 0.05 | $1.26 \times 10^{-75}$ |
| UNC5C | rs10516960 | 4 | 96159677 | T | G | 0.15 | 0.02 | $2.78 \times 10^{-9}$ |
| UST | rs1384668 | 6 | 149223723 | C | G | −0.38 | 0.03 | $6.60 \times 10^{-26}$ |
| VASN | rs886858 | 16 | 4422122 | A | G | 0.40 | 0.05 | $2.92 \times 10^{-16}$ |
| VIT | rs11124542 | 2 | 36994439 | A | C | 0.50 | 0.03 | $1.17 \times 10^{-50}$ |
| VSIR | rs9415993 | 10 | 73514911 | G | C | 0.57 | 0.04 | $3.79 \times 10^{-42}$ |
| VTN | rs704 | 17 | 26694861 | G | A | 1.11 | 0.02 | $2.76 \times 10^{-243}$ |
| VWA2 | rs11196687 | 10 | 116059728 | G | A | −0.75 | 0.05 | $6.72 \times 10^{-41}$ |
| WARS1 | rs4905953 | 14 | 100810426 | G | A | 0.36 | 0.04 | $6.37 \times 10^{-20}$ |
| WFDC1 | rs12448765 | 16 | 84425214 | A | G | −0.30 | 0.05 | $4.00 \times 10^{-11}$ |
| WFIKKN1 | rs8062289 | 16 | 681284 | C | T | 0.80 | 0.05 | $5.91 \times 10^{-59}$ |
| WFIKKN2 | rs7225019 | 17 | 48915879 | G | A | −1.06 | 0.04 | $7.23 \times 10^{-121}$ |
| XCL1 | rs1323532 | 1 | 168496272 | G | T | 0.27 | 0.04 | $1.41 \times 10^{-9}$ |
| ZG16B | rs4785908 | 16 | 2879279 | T | C | 0.56 | 0.05 | $6.04 \times 10^{-32}$ |

RSID stands for Reference SNP Identifier, CHR stands for Chromosome, POS stands for Position in base pairs, REF is the Reference Allele, ALT is the Alternative Allele.
